# “Invisible Killer”: Seasonal Allergies and Accidents

**DOI:** 10.1101/2023.09.08.23294325

**Authors:** Mika Akesaka, Hitoshi Shigeoka

**Author notes:** Corresponding author: Simon Fraser University, Department of Economics, 8888 University Drive, Burnaby, BC V5A1S6, CANADA, University of Tokyo, IZA, and NBER. Jack Madison provided superb research assistance. The authors thank Panle Jia Barwick, Robert Kaestner, Hiroyuki Kasahara, Munechika Katayama, Yoshifumi Konishi, Yuta Kuroda, Young Lee, Michelle Marcus, David Molitor, Claudia Persico, Julian Reif, Yutaro Sakai, Edson Severnini, Shinsuke Tanaka, Rusty Tchernis, Erdal Tekin, Shinsuke Uchida, Hendrik Wolff, and the seminar participants at Advances in Utilizing Mobile Location Data (Waseda University), AASLE 2022, AMES China 2022, AMES Tokyo 2022, AsianWEHE 2022, Japan Economic Association 2022, Hanyang-Kobe-Nanyang Conference, J-TREE, Kobe University, LACEA/LAMES 2022, NBER Health Economics Fall Meeting 2022, Osaka Metropolitan University, and University of Bologna, for their comments and suggestions. We thank the Fire and Disaster Management Agency of the Ministry of Internal Affairs and Communications and the National Policy Agency for providing us with ambulance records and police records. Akesaka acknowledges financial support from JSPS KAKENHI (23K12492, 22K01534, 21H04397, 20K13511) and Inamori Foundation. Shigeoka acknowledges financial support from JSPS KAKENHI (23H00828, 22H00057, 22H00847, 22H05009) and Tokyo Center for Economic Research. Any errors are our own. Kobe University, Research Institute for Economics and Business Administration, 2-1, Rokkodai, Nada, Kobe, Hyogo, 657-8501, JAPAN (Mika Akesaka).

## Abstract

Although at least 400 million people suffer from seasonal allergies worldwide, the adverse effects of pollen on “non-health” outcomes, such as cognition and productivity, are relatively understudied. Using ambulance archives from Japan, we demonstrate that high pollen days are associated with increased accidents and injuries— one of the most extreme consequences of cognitive impairment. We find some evidence of avoidance behavior in buying allergy products but limited evidence in curtailing outdoor activity, implying that the cognitive risk of pollen exposure is discounted. Our results call for governmental efforts to raise public awareness of the risks and promote widespread behavioral change.

## 1. Introduction

“Hay fever,” or seasonal allergic rhinitis (SAR) as it is medically known, is a common chronic condition induced by exposure to airborne allergens such as pollen and dust. SAR affects up to 30% of the population in developed countries, with an estimated 400 million sufferers worldwide (Greiner et al. 2011). Despite its widespread global prevalence, the adverse impact of pollen has, to date, received surprisingly little attention from economists. First, the symptoms of SAR tend to be chronic and relatively mild; however, the non-acute physiological effects of pollen exposure can be potentially problematic as people may not engage in avoidance behaviors to limit their exposure.

Second, in sharp contrast with *man-made* air pollution, the scope of governmental policy intervention for damage caused by *natural* sources, like pollen, is seemingly limited.^1^ However, a recent study projects that global warming will accelerate pollen production and thus increase pollen concentrations and prolong the duration of pollen seasons (Zhang and Steiner 2022). Figure 1 demonstrates the strong positive association between pollen counts and maximum temperature (Panel A) and the number of hot days above 30℃ (Panel B) in the previous summer in Japan—our study setting. The figure content highlights that climate change caused by human activities can exacerbate the potential damage arising from naturally occurring pollen production.

**Figure 1.**
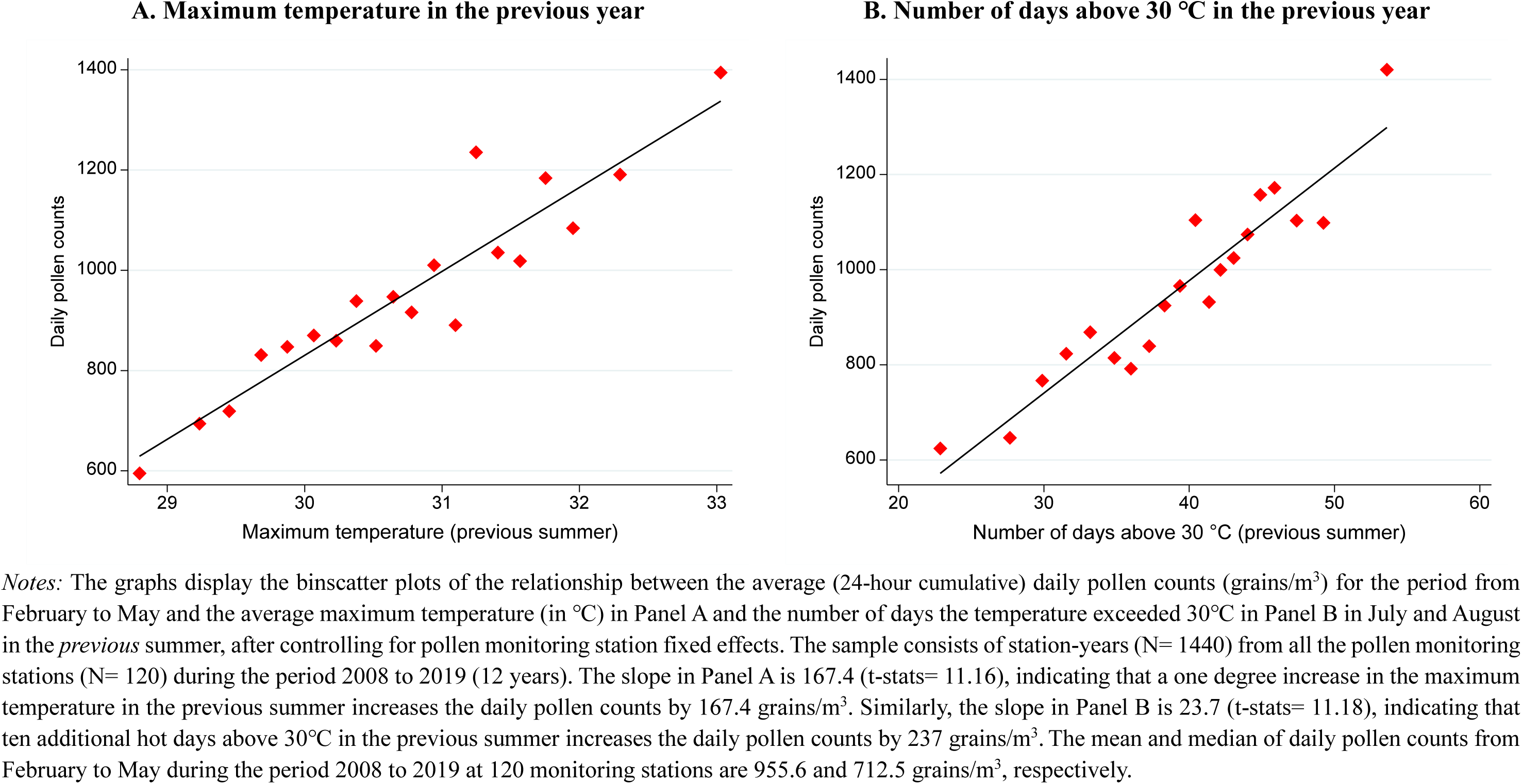
Pollen and temperature in the previous summer.

In addition to the apparent negative health consequences, clinical studies have identified the detrimental effects pollen exposure can have on cognitive performance, in the form of decreased attention span and increased reaction time (Wilken et al. 2002). However, existing studies examining the effects of seasonal allergies on “non-health” outcomes, such as cognition and productivity, in the *field* setting are limited to its influence on the test performance of children (Bensnes 2016; Marcotte 2015, 2017). We hypothesize that exposure to seasonal pollen has much broader consequences—beyond children’s test scores. If pollen exposure severely inhibits cognitive functioning, any daily activity that requires continuous attention and performance, such as driving, can be affected.

We conduct the first investigation of the effects of acute and short-term pollen exposure on the incidence of accidents. These include traffic accidents and work-related injuries, which are arguably some of the most extreme consequences of cognitive impairment. Traffic accidents are the leading cause of accidental deaths globally, and hence, any factor that influences the risk of traffic accidents is of great relevance to social welfare.^2^ Work-related injuries are also worth investigating as they translate to substantial productivity losses in the labor market.^3^ Furthermore, these accidents can cause negative externalities for individuals who do not suffer from seasonal allergies but are involved in the accidents.

We combine the data on pollen counts with the newly available administrative ambulance records covering all accidents that occurred from 2008 to 2019 in Japan. These accidents were especially severe as they required ambulance transportation to hospitals. Japan is the ideal setting to conduct such research as the pollen monitoring stations are densely located across the country at a rate 42 times higher than that of the United States. The intensity of pollen concentration varies widely across space and time, allowing us to accurately measure pollen exposure and examine the potential non-linear dose responses. Furthermore, we are able to provide nationally representative estimates, unlike the samples in previous studies that are limited to only a handful of schools or districts near pollen stations. This assists in mitigating concerns about the generalizability of our findings. We leverage daily levels of spatial and temporal variation in pollen counts of differing magnitudes to identify the effect of pollen exposure on accidents.

There are four primary findings. First, we find that high daily pollen counts are associated with increased accidents of all types, including traffic accidents, work-related injuries, sports injuries, and fire accidents. The relationship between pollen counts and the number of accidents is clearly concave, suggesting that even low concentrations of pollen—which are more frequent than higher concentrations—can have a significant adverse impact on cognitive performance and hence the incidence of accidents. While the effects are more pronounced for less severe accidents, elevated pollen exposure also increases accidents that lead to death.

These findings suggest that pollen exposure not only affects the school performance of children but also has a wider impact on cognition, productivity, and daily activities. For example, an increase in workplace injuries implies a detrimental effect on labor productivity. Therefore, the current estimated cost of exposure to airborne allergens, which is based mainly on straightforward health outcomes, school days missed, and work absenteeism, may severely underestimate the true cost to society.

Second, we find mixed evidence of short-term avoidance behaviors to reduce the risk of pollen-induced accidents. On the one hand, using in-home scanner data, we show that people increase spending on products that protect against seasonal allergies, such as medications, eye drops, and masks. On the other hand, we find limited evidence of individuals taking more serious measures to alleviate the risk of accidents, such as reducing outdoor activities; using geolocation data, we discover a very small reduction in the number of people outdoors in bustling areas on high pollen days.

These results indicate that people may lack awareness of or underestimate the cognitive risk of pollen exposure. Thus, the status quo that relies on individuals’ self-protection to mitigate pollen exposure may only lead to limited benefits. A government intervention that raises awareness regarding the risk that pollen exposure poses to health and cognition might be necessary, in addition to the provision of real-time pollen information. For example, public information campaigns, such as a “pollen alert”—recommending that allergy sufferers curtail outdoor activities and refrain from driving or operating machines on high pollen days—could be a cost-effective tool to promote widespread behavioral change and reduce pollen-induced accidents.

Third, we fail to find strong evidence of medium-term adaptation to the adverse impact of pollen exposure. The fundamental question relating to environmental hazards is whether such hazards are mostly unavoidable or whether individuals can adapt by utilizing current technologies.^4^ In the context of this study, newer medications for seasonal allergies have fewer side effects, including lower chances of drowsiness, which might reduce the risk of accidents caused by medication-induced drowsiness. In addition, one may have more time to invest in defensive strategies such as the purchase of an air purifier. We find that the detrimental effects of pollen exposure have not declined significantly in recent years, at least not within the past 12 years of our sample period. Furthermore, individuals who live in the high-average pollen regions have similar pollen-induced accident rates as those who live in the low-average pollen regions, corroborating previous findings.

Finally, we combine our estimates of the effect of pollen on accidents with projections of the future climate, and also the relationship between the temperature and pollen counts shown in Figure 1 to illustrate the magnitude of the social cost of anthropogenic climate change. The “business as usual” scenario from the Intergovernmental Panel on Climate Change (IPCC)— which predicts an increase of 4.1℃ in the summer temperature in the years 2076–2095 in Japan—would lead to 1,823 additional pollen-induced accidents annually. Multiplying the resulting accident counts with the average accident costs, the expected annual social cost of pollen-induced accidents is approximately $236 million. This estimated social cost is very likely to be a lower bound since we fail to consider the accidents at both sides of the severity distribution (i.e., minor cases as well as immediate deaths) that do not require ambulance transportation to hospital.

While much of the literature on climate change has focused on the impact of rising temperature on direct outcomes such as aggregate incomes, mobility, mortality, and agricultural outcomes (Carleton and Hsiang 2016; Dell et al. 2014), the increase in the number of seasonal allergy sufferers and the associated performance impairment could be the indirect and undiscovered cost of anthropogenic climate change. Consequently, any measure to mitigate the risk of a warming climate could have substantial social benefits by preventing temperature-driven increases in airborne pollen.

The rest of the paper is organized as follows. Section 2 briefly describes the background, Section 3 describes the data, Section 4 presents the econometric model, and Section 5 reports the main findings of this study. Section 6 examines the short-term avoidance behaviors, and Section 7 conducts additional analysis. Section 8 offers conclusions.

## 2. Background

### 2.1. Pollen and seasonal allergies

SAR is a common chronic condition that occurs when an individual’s immune system reacts to allergens in the air, such as pollen and dust, causing the immune system to produce antibodies (such as histamines and cytokines) to fight the perceived threat from the pollen grains. The antibodies, in turn, cause inflammation in the airways, inducing various allergic symptoms, such as a runny nose, nasal congestion, sneezing, and itchy eyes (Greiner et al. 2011).

SAR is a global health problem as otherwise seemingly healthy individuals can be affected. The prevalence rates vary by country but are generally between 10% and 30% in developed countries (Greiner et al. 2011; Schmidt 2016). This number is likely to underestimate the true prevalence rate as some individuals do not seek medical assistance for the condition. Furthermore, numerous studies document increasing prevalence due to various factors such as urbanization, westernization of lifestyles, and climate change (Schmidt 2016).

Japan is no exception. An extensive epidemiological survey guided by the Japan Society of Immunology and Allergology in Otolaryngology for otolaryngologists and their families has been conducted every ten years (1998, 2008, and 2019). According to this study, the prevalence rate of SAR has steadily increased by roughly 10 percentage points every ten years: from 19.6% in 1998 to 29.8% in 2008 and further to 42.5% in 2019. The prevalence rate has been found to peak around middle age, but a substantial number of young and older individuals also suffer from SAR (Matsubara et al. 2020).^5^

Because SAR has relatively mild and chronic symptoms, its economic cost has been overlooked. In addition to direct medical costs such as medications and emergency department visits (Xing et al. 2023), past studies suggest that pollen allergies are a major source of absenteeism from work and school (Hellgren et al. 2010; Lamb et al. 2006).

Most relevant to this study, clinical studies have identified the negative effects of SAR on cognitive performance. This often occurs indirectly as a result of deteriorated sleep quality (Craig et al. 2004; Santos et al. 2006) and directly by antibodies themselves via brain function (McAfoose and Baune 2009). For example, Wilken et al. (2002) find that allergic adults who are randomly exposed to pollen perform worse on a broad range of cognitive measures than those who are not exposed; these measures include longer response times, reduced working memory, divided attention, and slower computation. Unfortunately, medical studies have shown that allergy medications can induce similar or even worse effects on cognitive functioning due to various side effects, such as drowsiness, dry mouth, and lethargy (Jáuregui et al. 2009; Kay 2000).

To date, existing studies of the effect of seasonal allergies on cognition and productivity in the *field* setting have been limited to their effect on the test performance of children (Bensnes 2016; Marcotte 2015, 2017). However, as pollen exposure impairs cognitive performance, almost any daily activity can be severely affected. In this paper, we focus on accidents, including traffic collisions and work-related injuries, as they involve the most extreme forms of performance impairment. For example, it is well established that cognitive function is negatively correlated with the likelihood of motor vehicle accidents (Anstey et al. 2005, 2012).^6^ Similarly, workplace injuries are most commonly caused by distractions (European Commission 2009).

In summary, while scientific knowledge concerning the impact of other environmental stressors (such as pollution and temperature) on cognitive performance and productivity has been gained^7^, the impact of airborne pollen and associated seasonal allergies on these outcomes has yet to be sufficiently examined.

### 2.2. Warming climate and pollen

Because higher temperatures and carbon dioxide (CO2) concentrations have been found to increase pollen production, climate change is expected to impact pollen concentrations and the duration of pollen seasons significantly. Anderegg et al. (2021) track pollen trends across 60 pollen stations in the United States from 1990 to 2018 and find increases of 20.9% and 21.5% in the annual and spring (February–May) pollen concentrations, respectively; the pollen season start date occurred 20 days sooner, and the pollen season duration increased by eight days. They further conduct a model selection analysis to identify the main drivers of pollen proliferation and find that mean annual temperature is the strongest predictor of the above-mentioned pollen metrics among eight climate variables (including temperature, precipitation, frost days, and CO2 concentrations). Increased pollen quantities, an earlier pollen season start date, and prolonged duration of the pollen season have similarly been observed in Europe (D’Amato et al. 2007; Ziello et al. 2012; Hamaoui-Laguel et al. 2015).

We confirm that such a relationship is also observed in the data we collected and analyzed in Japan. Figure A1 displays the time series of the average daily pollen counts for the period from February to May and the average maximum temperature (Panel A), as well as the number of days where the temperature exceeded 30℃ (Panel B) in July and August in the *previous* summer, using station-year panel data from all the pollen monitoring stations (N= 120) during the period from 2008 to 2019. Pollen counts are expressed in (24-hour cumulative) particles per cubic meter of air (grains/m^3^). Both graphs show a positive relationship between the temperature in summer and the pollen counts the following spring. For example, a cold summer was experienced in 2009, and the pollen concentration in the spring of 2010 was relatively low. By contrast, a hot summer was experienced in 2010, and high pollen counts were observed in the spring of 2011.

Figure 1, mentioned in the introduction, presents the associations between pollen counts and temperatures in the previous summer using the same data. The binscatter plot exploits the variation in pollen counts *within* the same pollen monitoring station over time by controlling for station fixed effects (FEs). The linear slope of 167.4 (t-stats= 11.2) in Panel A indicates that an increase of 1℃ in the maximum temperature in the previous summer is associated with an average additional 167 grains/m^3^ daily pollen count the following spring. As the mean and median daily pollen counts of 120 stations in the same period are 955 and 712 grains/m^3^, respectively, such an increase may be sizable. Similarly, the slope in Panel B is 23.7 (t-stats= 11.2), indicating that ten more hot days above 30℃ in the previous summer could increase the daily pollen count by 237 grains/m^3^ the following spring.

To summarize, the evidence thus far indicates that climate change caused by human activities has increased the intensity of pollen seasons in different parts of the world. The expected temperature rise due to global warming is likely to intensify and accelerate this trend in the coming decades (Ziska et al. 2019). For example, Zhang and Steiner (2022), project that climate change will *further* hasten the arrival of the pollen season (by up to 40 days), prolong the duration of the pollen season (by approximately 19 days), and, as a result, increase the annual total pollen emission (from 24% to 40%) in the United States.

## 3. Data

This study has compiled a comprehensive dataset to study the impact of pollen exposure on accidents. For our primary analysis, we combine the daily airborne pollen counts with newly available ambulance records that report accidents from 2008 to 2019 in Japan. To our knowledge, this is the first study to use this dataset for economic research. To avoid confusion, supplementary data used to examine symptoms (Google Trends and Twitter data, Section 5.1) and avoidance behaviors (in-home scanner data and geolocation data, Sections 6.2 and 6.3) are described later. The details of the data sources are provided in Appendix H.

### 3.1. Airborne pollen

We obtain airborne pollen data from the Japanese Ministry of the Environment’s pollen monitoring system (designated “Hanako-san”), which provides hourly readings of pollen counts (grains/m^3^) for Japanese cedar and hinoki cypress (Yamada et al. 2014). Figure A2 displays the pollen season calendar for typical plants, showing that these two types of plants are the primary source of pollen production in Japan, mainly for February to May. Comprehensive data on pollen counts are available beginning from 2008. This information is automatically published on the Ministry of the Environment’s website. In addition, during the pollen season, the pollen level is commonly reported in the weather forecast, along with the usual temperature and precipitation. See Figure 2 for real-time and forecasted pollen levels reported on television on a typical day during the pollen season in Japan. Thus, the cost of acquiring such information is nearly zero.

**Figure 2.**
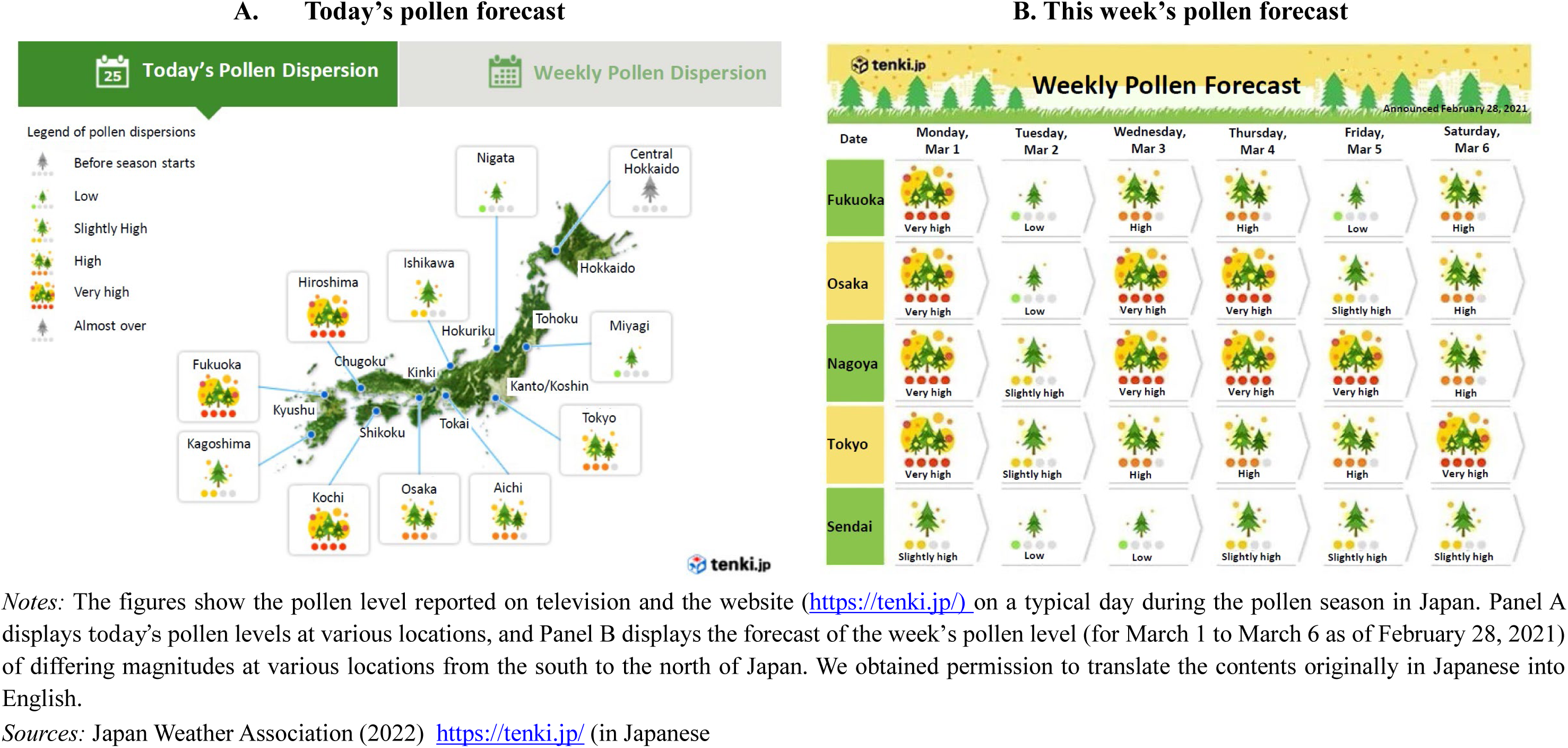
Today’s and weekly forecasted pollen levels.

A total of 120 pollen monitoring stations are located across Japan. Panel A of Figure 3 displays the locations of all monitoring stations as of 2019.^8^ Each of the 46 prefectures has, on average, two to three monitoring stations, both in urban areas with high population densities and in mountainous regions, which are the major sources of pollen production (Wakamiya et al. 2019).^9^ The number of pollen monitoring stations is extremely high, given the size of the country. For example, the United States—which is 26 times larger than Japan—has only 74 pollen monitoring stations nationwide. This suggests that the density of pollen stations is *42 times* higher in Japan than it is in the United States.

**Figure 3.**
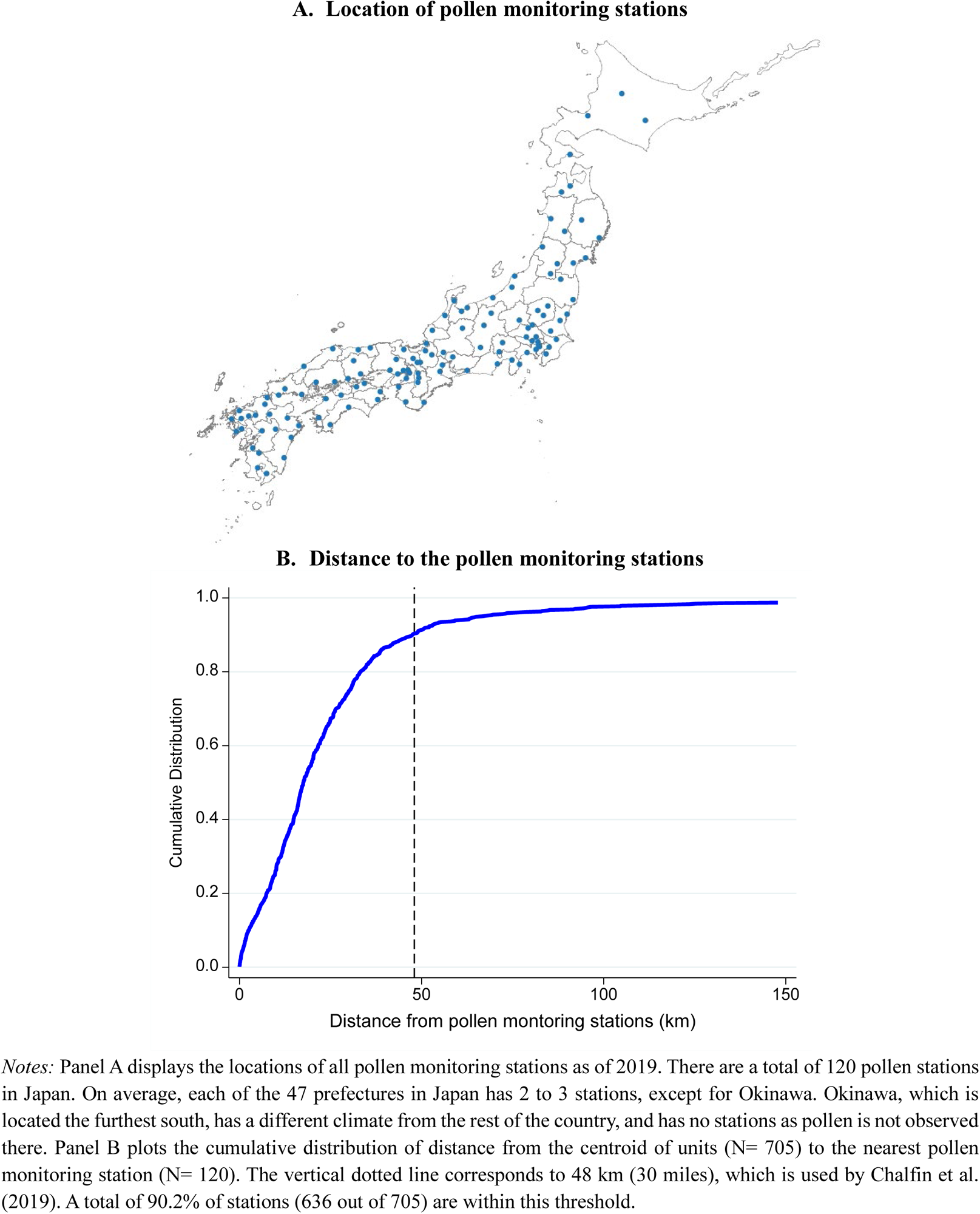
Pollen monitoring stations.

Since typical pollen in Japan can be distributed more than 100 km (160 miles) away and remain in the air for more than 12 hours, almost entire areas of Japan, even in sparsely forested cities, can be contaminated by airborne pollen (Yamada et al. 2014). Panel B of Figure 3 plots the cumulative distribution of the distance from the nearest pollen station to the centroid of each emergency response unit (our regional unit of analysis, as explained in Section 3.2). The mean and median distances from pollen stations are 25.4 and 17.5 km, respectively. Even with a conservative threshold of 48 km (30 miles) for pollen measurements to be valid (Chalfin et al. 2019), 90.2% of all units are within this threshold.^10^

The high density of stations across the country enables us to (i) accurately measure the exposure to pollen, (ii) provide nationally representative estimates of pollen exposure (unlike previous studies that are limited to a handful of schools or districts near pollen stations), and (iii) include the continuous pollen exposure variable with differing magnitudes as the regressor to investigate the potential non-linearity in dose-response (unlike past studies that only included a dichotomous variable that defined high pollen days).

As the pollen emission from the Japanese cedar (the main pollen-producing plant) starts in February and peaks from March to April (as shown in Figure A2), the pollen count is monitored from February to May every year.^11^ We aggregate the monitor readings to obtain the daily level by adding the hourly observations to calculate the accumulated number of pollen grains counted in 24 hours. In addition, weather covariates from nearby weather stations are included in the same data. In particular, hourly temperature, precipitation, and wind speed are recorded. Similarly, we aggregate these variables to obtain the daily level.

### 3.2. Ambulance records

Our data relating to accidents and injuries are derived from the Fire and Disaster Management Agency (FDMA) of the Ministry of Internal Affairs and Communications, Japan. The data cover all ambulance calls, except for those in metropolitan Tokyo,^12^ for the period 2008 to 2019 that required ambulance transportation to hospitals. The registration of all ambulance records in the online system of FDMA became mandatory in 2008. As ambulance service is provided to the public free of charge in Japan, there is no differential selection of the sample by socioeconomic status, unlike in countries like the United States, where ambulance use varies by health insurance status (Meisel et al. 2011).

A total of 14.7 million accidents are recorded for the period 2008 to 2019, with a yearly average of 1.2 million accidents.^13^ The data provide detailed information for individual accidents, including the location of the accidents, date and time of the ambulance calls, type of accidents, severity of injuries, and age and gender of those involved in the accidents.

Two features of this dataset are particularly beneficial for our study. The first feature is that we have knowledge of the type of accidents that occurred, including traffic accidents, work-related injuries, sports injuries, and fire accidents. The other distinctive feature is that it includes information on the severity of injuries sustained. This measure is expected to be accurate as it is based on the initial clinical assessment by the doctors who first attended to the injured person at hospital admission. The severity measure ranges from death/fatal, serious, moderate, to light, where “serious” accidents require more than three weeks of hospitalization and treatment; “moderate” accidents require hospitalization, but for less than three weeks; and “light” accidents do not require hospital admission. Ambulance records have two key advantages over vital statistics: They can capture non-fatal but severe injuries that would be missed in vital statistics, and they can capture the accident data on the day of occurrence without measurement errors, unlike vital statistics, which may record some accidental deaths with delays.

The geographical unit in ambulance records is an emergency response unit (hereinafter referred to as “unit”), which is the level of ambulance service in Japan. While many units are municipalities themselves, some small municipalities form a unit to increase the efficiency of the ambulance service. As of 2019, 1,700 municipalities (equivalent to counties in the United States) across 46 prefectures (equivalent to states in the United States) form 705 units. We aggregate the accident records to the unit-day level by adding the hourly observations within the units.^14^

### 3.3. Sample construction and summary statistics

To construct our primary sample, we merge unit-day level ambulance records with the same-day daily level pollen counts from the nearby monitoring stations. Thus, the primary sample comprises records from February to May (i.e., high pollen seasons only) for all prefectures except for Hokkaido, which records from March to June, for 2008 to 2019.

Table 1 presents summary statistics for our primary estimation sample, which consists of 970,309 unit-day observations. On average, there are 33 daily accidents per million people. Traffic accidents are the leading type of accident and account for 37.6% of all accidents. This is followed by work-related injuries (3.5%), sports injuries (2.6%), and fire accidents (0.5%). The other accidents that do not belong to any of these categories accounted for over half of all accidents (55.8%).^15^

**Table 1.** Summary statistics.

| Variables | Share within category | Obs | Mean | Std. dev. | Min | Max |
| --- | --- | --- | --- | --- | --- | --- |
| **A. Outcomes (per 1,000,000 per day)** |  |  |  |  |  |  |
| All accidents |  | 970,309 | 33.03 | 28.23 | 0 | 5,091 |
| Type: Traffic accidents | 37.6% | 970,309 | 12.41 | 16.09 | 0 | 5,091 |
| Type: Work-related injuries | 3.5% | 970,309 | 1.15 | 3.87 | 0 | 1,367 |
| Type: Sports injuries | 2.6% | 970,309 | 0.86 | 3.24 | 0 | 1,195 |
| Type: Fire accidents | 0.5% | 970,309 | 0.17 | 2.10 | 0 | 2,612 |
| Type: Other accidents | 55.8% | 970,309 | 18.44 | 17.77 | 0 | 2,447 |
| Severity: Death/Fatal | 0.9% | 970,309 | 0.30 | 2.11 | 0 | 943 |
| Severity: Serious | 6.2% | 970,309 | 2.05 | 6.07 | 0 | 1,572 |
| Severity: Moderate | 27.4% | 970,309 | 9.05 | 11.90 | 0 | 1,958 |
| Severity: Light | 65.4% | 970,309 | 21.58 | 20.65 | 0 | 4,570 |
| Ages: 0-24 years | 20.5% | 970,309 | 6.34 | 9.44 | 0 | 2,112 |
| Ages: 25-44 years | 15.0% | 970,309 | 4.61 | 7.64 | 0 | 2,186 |
| Ages: 45-64 years | 18.8% | 970,309 | 5.81 | 8.90 | 0 | 2,637 |
| Ages: 65 years and older | 45.7% | 970,309 | 14.09 | 15.98 | 0 | 2,695 |
| Gender: Male | 53.3% | 970,309 | 15.88 | 17.55 | 0 | 4,769 |
| Gender: Female | 46.7% | 970,309 | 13.91 | 15.82 | 0 | 2,366 |
| Location: Roads | 44.9% | 970,309 | 8.73 | 12.66 | 0 | 198 |
| Location: Home | 34.0% | 970,309 | 6.61 | 9.90 | 0 | 82 |
| Location: Public space | 18.1% | 970,309 | 3.51 | 5.73 | 0 | 51 |
| Location: Workplace | 3.0% | 970,309 | 0.58 | 1.14 | 0 | 58 |
| **B. Regressors (per day)** |  |  |  |  |  |  |
| Pollen counts (grains/m<sup>3</sup>) |  | 970,309 | 984.34 | 2135.26 | 0 | 55,104 |
| Logged (Pollen counts) |  | 970,309 | 0.16 | 0.43 | 0 | 10 |
| Precipitation (mm) |  | 970,309 | 11.90 | 6.14 | 0 | 28 |
| Average temperature (°C) |  | 970,309 | 2.93 | 1.39 | 0 | 16 |
| Average wind speed (m/s) |  | 970,309 | 10.48 | 1.28 | 7 | 13 |
| Darkness (hours) |  | 852,948 | 2.20 | 2.16 | 0 | 521 |
| SO<sub>2</sub> (ppb) |  | 846,719 | 6.14 | 10.07 | 0 | 307 |
| NO<sub>2</sub> (ppb) |  | 846,659 | 15.11 | 9.60 | 0 | 88 |
| CO (0.1ppm) |  | 848,798 | 4.02 | 1.84 | 0 | 61 |
| OX (ppb) |  | 850,847 | 36.76 | 11.51 | 0 | 120 |
| PM10 (µg/m<sup>3</sup>) |  | 850,577 | 20.04 | 11.14 | 0 | 299 |

*Notes:* The sample is derived from the ambulance records for the period 2008 to 2019, and the level of observation is a unit per day. There are 705 units in total. The population in each unit is used as weights. The share within the *category* should be summed to 100%. The pollution data are available from 2009 onwards.

The mean daily concentration of airborne pollen is 984 grains/m^3^ (with a standard deviation of 2,135).^16^ Figure 4 plots the daily pollen counts for five selected locations from the north to the south of Japan, demonstrating substantial variations in pollen concentration across space and time. As pollen counts are rightly skewed, we take the natural log of pollen counts as the main regressor throughout the study unless otherwise mentioned. Panel A of Figure A3 displays the histogram of the logged daily pollen counts during the period 2008 to 2019, while Panel B displays the same counts after residualization by the unit, month-by-year, month-by-prefecture, and day-of-the-week FEs. This figure demonstrates substantial residual variation in pollen concentrations, indicating ample variation for obtaining precise estimates.

**Figure 4.**
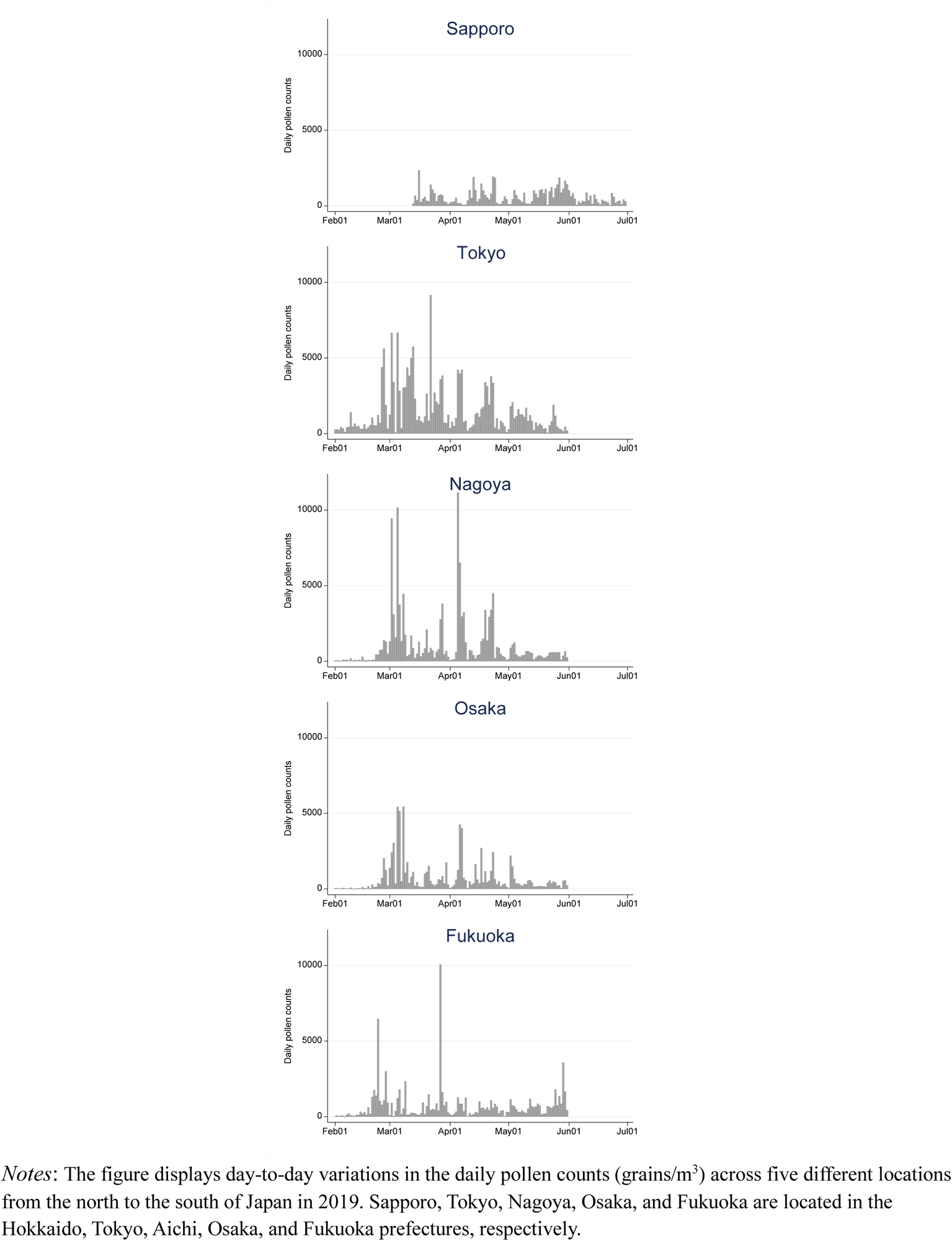
Daily pollen counts for selected locations.

## 4. Econometric model

We estimate the effect of short-run exposure to pollen on the number of accidents, net of any potentially confounding factors:

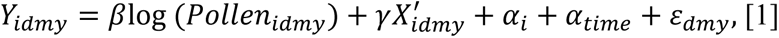

where the dependent variable *Y_idmy_* is the number of accidents per million people in unit *i* on day *d* in month *m* and year *y*. The logged form of the main regressor (pollen counts) is consistent with the non-linearity in dose-response in clinical studies (Erbas et al. 2007).^17^ We later show the results from the alternative specifications (e.g., level-level or “dose-response”) or Poisson model to account for the count nature of the accidents and assess the sensitivity of our results to zero observations. The parameter of interest is β, which measures the change in the outcome associated with a 100% increase in pollen counts. Unit FE (α_*i*_) controls for geographic differences in health and pollen concentrations.

The high granularity of our data allows us to include multiple sets of high-dimensional time FEs (α*_time_*). The baseline specification includes prefecture-by-month (α*_pm_*), month-by-year (α*_my_*), and day-of-the-week FEs (Deryugina et al. 2019). We later replace these with more or less stringent ones as a robustness check. Prefecture-by-month FE controls any seasonal correlation between pollen counts and accidents, allowing this correlation to vary by prefecture. The month-by-year FE controls flexibly for nationwide time-varying shocks during our sample period. Finally, day-of-the-week FE accounts for within-week variation in accidents. In this way, we compare days in the same month in the same unit that happen to differ in pollen concentration across years, alleviating concerns that other seasonal trends in accidents could affect the results.

The 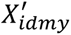 flexibly controls for daily variation in weather covariates. We include seven indicators for 5℃ intervals of daily average temperatures, ranging from 0℃ or less to 25℃ or more. For daily precipitation, we include four indicators (no rain, less than 1 mm of rain, 1 mm to 2 mm of rain, and more than 2 mm of rain). We also control for the average wind speed and duration of darkness—the time between dusk and dawn, which is an important unobserved factor for traffic accidents (Bünnings and Schiele 2021). Finally, we control the logged population, which is related to population density and congestion (once with unit FE included), potentially affecting the risk of accidents (Abouk and Adams 2013).

The underlying assumption for β to reflect the causal impact of pollen is that once we control for the series of location and time FE as well as weather covariates, the temporal, seasonal, and geographic variations in daily pollen counts would be considered as good as random. This assumption is arguably plausible with the very granular sets of FE and daily variation in pollen counts from naturally occurring processes.

We cluster all standard errors at the pollen monitoring station (N= 120)—the level of underlying variation in our treatment variable (Abadie et al. 2017)—to account for possible serial correlation, and weight all estimates by the relevant population in cases where the dependent variable is in per capita terms.

## 5. Main results

### 5.1. “First stage”—Symptoms of seasonal allergies

Before revealing our main findings, we present evidence of a “first stage”—whether people are more likely to suffer from seasonal allergy symptoms on high pollen days than low pollen days—using data on both internet search activities (Google Trends) and social media posts (Twitter data).

We use publicly available Google Trends data on two broad categories: (1) pollen-related keywords, and (2) symptoms-related keywords during the period 2016 to 2019 at the prefecture-day level (N= 21,551). See Table B1 for the complete list of search keywords used. The Google search index reflects search term popularity and takes a value of 0 to 100 in a given prefecture and on a given day in proportion to total searches within the search period.^18^

Figure 5 displays the results of symptom-related keywords (“runny nose,” “nasal congestion,” “sneeze,” and “itchy eyes” from Category 2 in Table B1). Panel A shows the time series of daily pollen counts (grains/m^3^) and the Google search index for these keywords in 2018 as an example. The variables move closely over time. Panel B displays the binscatter plots of the relationship between the logged daily pollen counts and search index after controlling for the month-by-prefecture, month-by-year, and day-of-the-week FEs. The linear lines fit the data well. A 100% increase in daily pollen counts leads to increases in the search index by 3.6 on a 0–100 scale, with a mean of 30.4 (*p*-values<0.01). Similar patterns are observed for pollen-related keywords in Figure B1.

**Figure 5.**
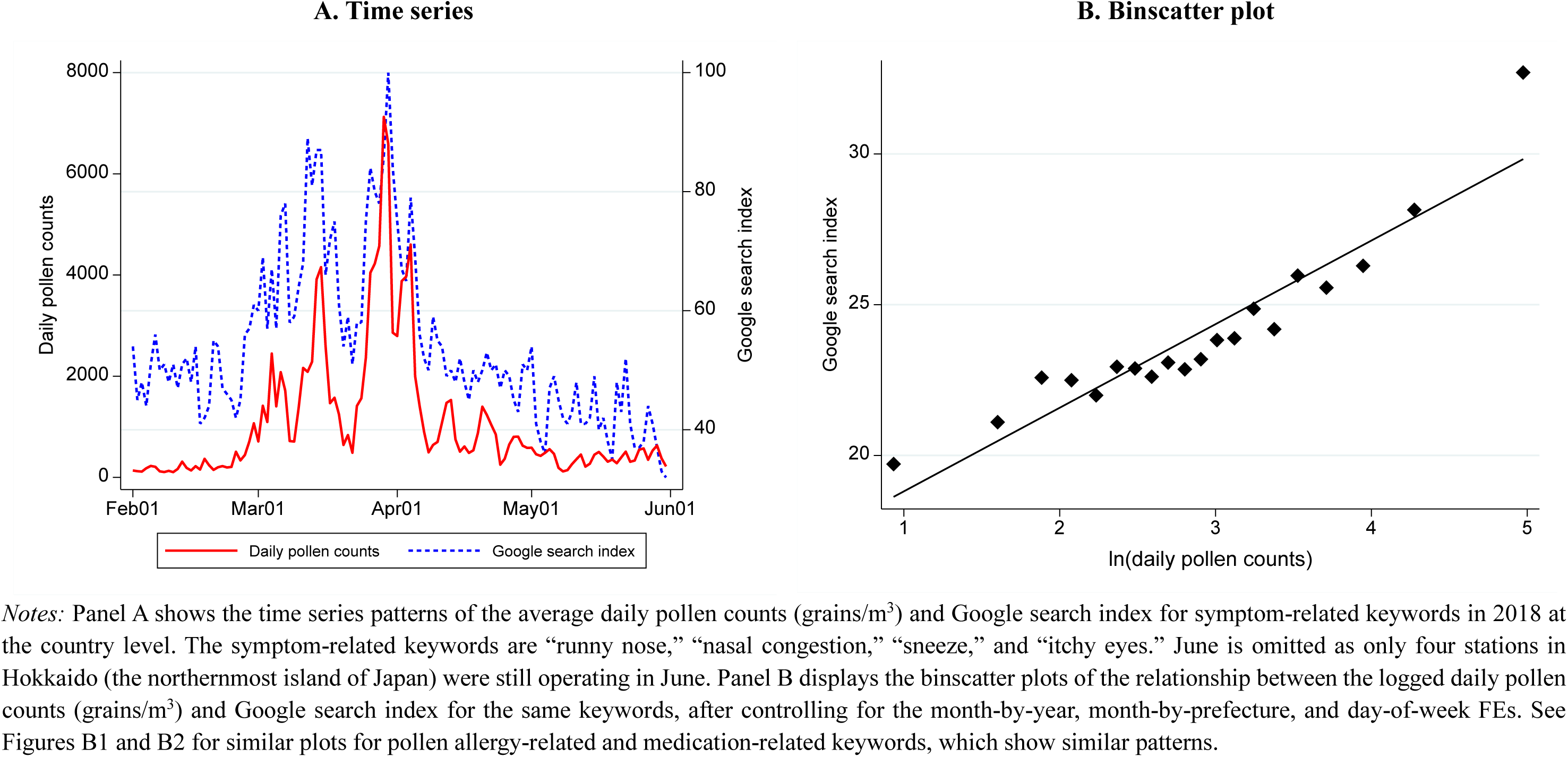
Pollen and symptom-related Google search index.

We replicate the same relationship between pollen counts and keywords using public Twitter data for the period 2016 to 2019 at the prefecture-day level.^19^ The only difference from the above analysis is that the dependent variable is now the number of tweets that contain the same two sets of keywords. Figure B2 demonstrates that people similarly tend to tweet these keywords more on high pollen days.

One advantage of Twitter data over Google Trends data is that while the sample is more skewed to younger cohorts, people tend to express the status of their “feelings” in tweets (Baylis 2020; Burk et al. 2022). Thus, in addition, we collect data pertaining to the number of tweets related to sleep, namely, “having a hard time falling asleep” and “feeling sleepy,” as clinical studies suggest that decreased sleep quality is one way in which pollen exposure worsens cognitive functioning (Craig et al. 2004; Santos et al. 2006). Figure B3 shows that individuals seem to have more sleep-related issues as the pollen level increases. This “first stage” evidence shows that individuals do suffer from typical seasonal allergy symptoms and that some of them are aware of being exposed.

### 5.2. Basic results

Figure 6 displays the binscatter plots of the relationship between the logged daily pollen counts (grains/m^3^) and the number of accidents per million people: all accidents (Panel A), followed by each type of accident by size (except for “other” accidents), namely, traffic accidents (Panel B), work-related injuries (Panel C), sports injuries (Panel D), fire accidents (Panel E), and other accidents (Panel F), after controlling for the unit, month-by-year, month-by-prefecture, and day-of-the-week FEs. All figures display a relatively linear relationship with the logged pollen counts, slightly flattening at a very high pollen concentration level. This simple plot of raw data (net of location and time FEs) reveals a strong relationship between the pollen concentration and the number of accidents. We formally test this relationship below.

**Figure 6.**
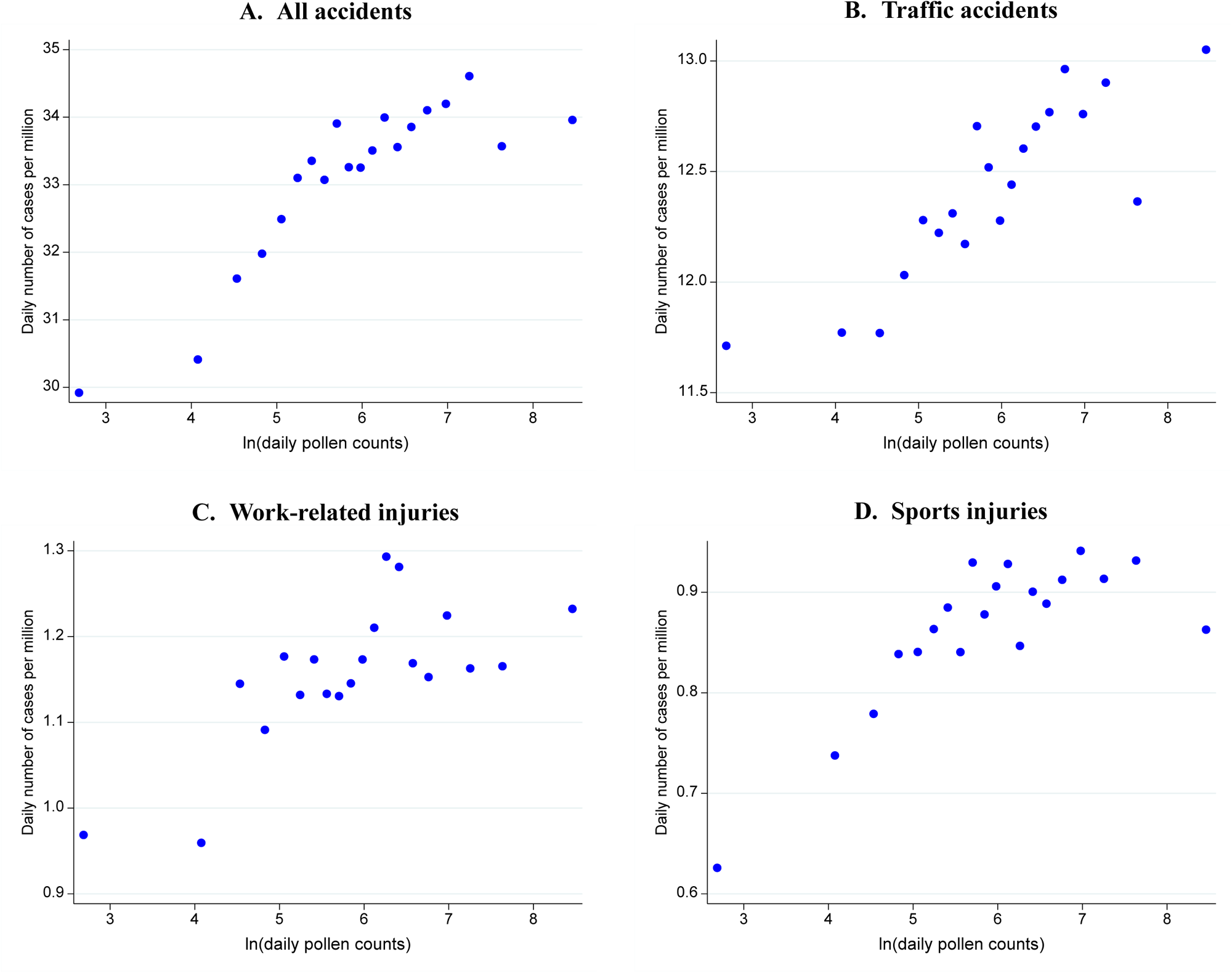
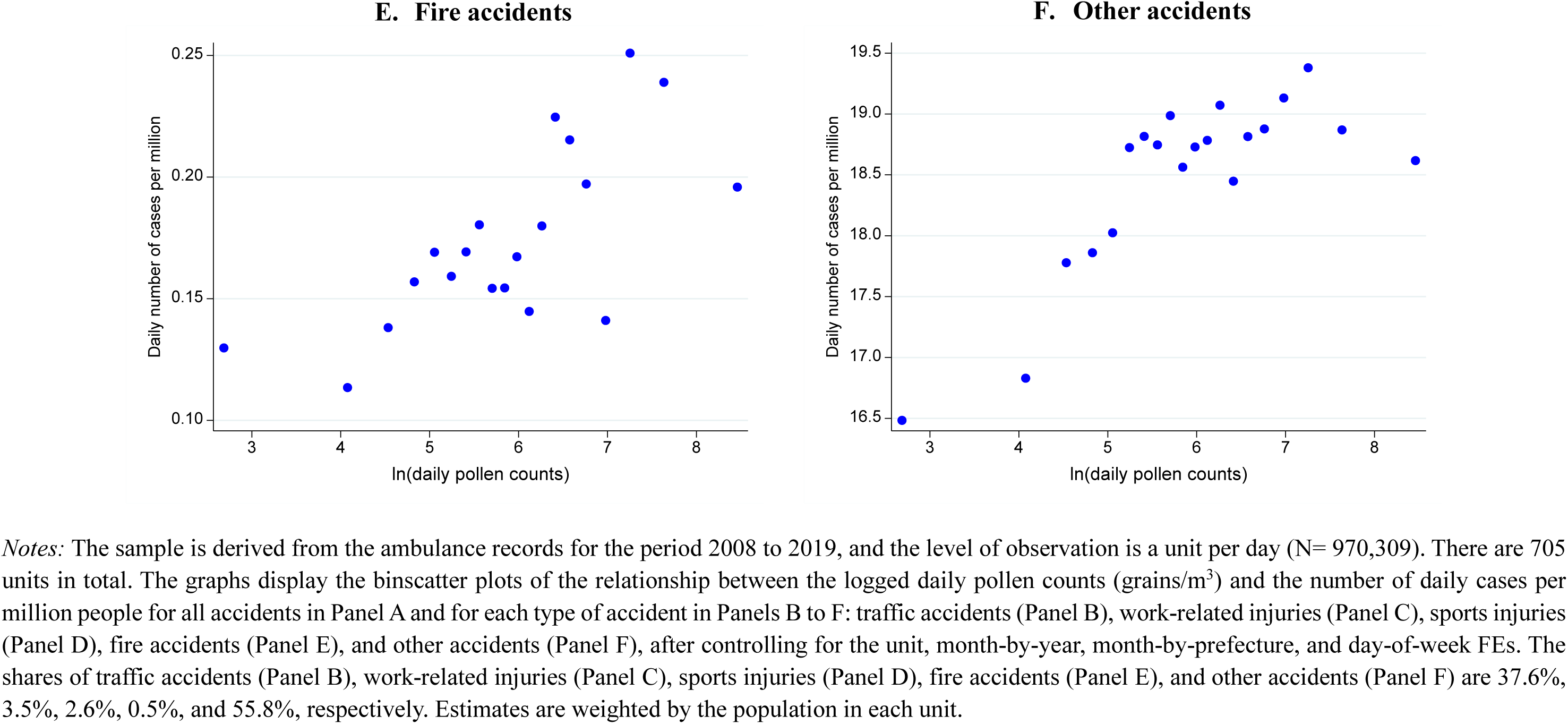
Pollen and the number of accidents.

Table 2 reports the main estimates from Equation [1]. Column (1) indicates that a 10% increase in pollen count leads to an increase in the number of accidents by 0.0231 (0.231×0.1) per million people, which is precise and highly statistically significant (*p*-value<0.001, t-stats= 14.0). Columns (2) to (5) show that, while the magnitude varies by accident type, the elevated pollen concentration increases all types of accidents. In particular, while the share is small (3.5%), the increased incidence of work-related injuries indicates that labor productivity, in general, is likely to be affected by pollen exposure. Owing to space considerations, we focus on the categories “all accidents” and “traffic accidents” from here on. We choose traffic accidents as they make up 37.6% of accidents that occur. We discuss the monetary values of pollen-induced accidents in Section 7.2, where we project the potential damages from climate change.

**Table 2.** Main results: Accidents.

|  | **A. All accidents** | **B. By type** |  |  |  |  |
| --- | --- | --- | --- | --- | --- | --- |
|  |  | Traffic accidents | Work-related injuries | Sports injuries | Fire accidents | Other accidents |
|  | (1) | (2) | (3) | (4) | (5) | (6) |
| ln(pollen counts) | 0.231***<br>(0.020) | 0.079***<br>(0.012) | 0.012***<br>(0.002) | 0.007**<br>(0.003) | 0.006***<br>(0.002) | 0.127***<br>(0.016) |
| R-squared | 0.46 | 0.24 | 0.06 | 0.08 | 0.00 | 0.37 |
| N | 970,309 | 970,309 | 970,309 | 970,309 | 970,309 | 970,309 |
| N of units | 705 | 705 | 705 | 705 | 705 | 705 |
| N of clusters | 120 | 120 | 120 | 120 | 120 | 120 |
| Mean of dep. Var | 33.03 | 12.41 | 1.15 | 0.86 | 0.17 | 18.44 |
| *Share* | *100%* | *37.6%* | *3.5%* | *2.6%* | *0.5%* | *55.8%* |
| Unit FE | X | X | X | X | X | X |
| Day-of-week FE | X | X | X | X | X | X |
| Month-year FE | X | X | X | X | X | X |
| Month-prefecture FE | X | X | X | X | X | X |

Importantly, our estimates provide a lower bound on the effect of pollen on accidents as the ambulance records analyzed do not include accidents on the extremes of the severity distribution (i.e., minor cases and immediate deaths, which do not require ambulance transportation to hospitals). In addition, the severity of injuries is assessed at the time of hospital admission, and this may have led to an underestimation of eventual deaths.^20^

#### Dose-response

Figure C1 plots the estimates from a non-parametric binned regression to examine dose responses more flexibly. Specifically, the logged daily pollen counts in Equation [1] is replaced by dummies for each decile of daily pollen in *levels*. Panel A for all accidents reveals a clear concave relationship, suggesting that even low concentrations of pollen—which are more frequent than higher concentrations—may have a meaningful impact on cognitive performance and hence the incidence of accidents. Notably, this concave dose-response function also seems to indicate the benefits of reducing pollen concentrations even for countries with lower pollen levels than those in our setting. In addition, the shape of the function explains why the level-log specification in Equation [1] fits the data well.

### 5.3. Robustness

Our results on the effect of pollen on accidents are robust to a battery of specification checks, including different types of location and time FEs, different ways of constructing regressors, as well as outcomes, alternative specifications, and placebo exercises.

#### Robustness

Table 3 provides the results of robustness checks and extensions. Our results are robust to different ways of constructing the pollen concentration, including an inverse distance weighted average of three nearby stations (Columns (2) and (3)), adding pollution covariates^21^ and the full interaction of temperature and rain dummies (Columns (4) and (5)), limiting the sample to units within 48 km from pollen monitoring stations to reduce measurement errors (Column (6)), and an unweighted ordinary least square (Column (7)).

**Table 3.** Robustness: Accidents.

|  | (1) | (2) | (3) | (4) | (5) | (6) | (7) | (8) | (9) | (10) |
| --- | --- | --- | --- | --- | --- | --- | --- | --- | --- | --- |
|  |  | Construction of pollen measures |  | Other |  |  |  | Expanding windows of outcome |  | Collapse data at the weekly level |
|  | Baseline | Weighted average of nearby three stations | Drop zero pollen observations | (1)+Add pollution | (1)+Add temperature & rain interactions | Units within 48 km from stations | Un-weighted | Add the following one day | Add the following two days |  |
| A. All accidents |  |  |  |  |  |  |  |  |  |  |
| ln(pollen counts) | 0.231***<br>(0.020) | 0.234***<br>(0.021) | 0.249***<br>(0.021) | 0.220***<br>(0.022) | 0.205***<br>(0.020) | 0.229***<br>(0.020) | 0.402***<br>(0.066) | 0.236***<br>(0.037) | 0.265***<br>(0.052) | 0.213***<br>(0.036) |
| R-squared | 0.46 | 0.46 | 0.46 | 0.47 | 0.46 | 0.48 | 0.33 | 0.62 | 0.71 | 0.85 |
| N | 970,309 | 970,309 | 962,255 | 814,578 | 970,309 | 872,227 | 970,309 | 961,901 | 953,522 | 147,066 |
| B. Traffic accidents |  |  |  |  |  |  |  |  |  |  |
| ln(pollen counts) | 0.079***<br>(0.012) | 0.082***<br>(0.012) | 0.090***<br>(0.013) | 0.073***<br>(0.013) | 0.063***<br>(0.012) | 0.078***<br>(0.012) | 0.137***<br>(0.037) | 0.086***<br>(0.019) | 0.110***<br>(0.027) | 0.124***<br>(0.020) |
| R-squared | 0.24 | 0.24 | 0.24 | 0.24 | 0.24 | 0.26 | 0.12 | 0.38 | 0.48 | 0.70 |
| N | 970,309 | 970,309 | 962,255 | 814,578 | 970,309 | 872,227 | 970,309 | 961,901 | 953,522 | 147,066 |

To address the possibility that the effect of pollen can potentially appear with lags, we extend the outcome window to the following day and then to the following two days (i.e., *t* and *t*+1, or *t*, *t*+1, and *t*+2) in Columns (8) and (9) of Table 3 (Deryugina et al. 2019). These estimates are slightly larger than the contemporaneous effect in Column (1), implying that we may even have underestimated the effect of pollen. The short-lived effect is not surprising; while pollen triggers an allergic reaction within minutes after SAR sufferers are exposed, its effects generally do not last more than four to eight hours (Skoner 2001).^22^ Our results are also robust to aggregating data at the weekly level to net out any temporal displacement and capture dynamic effects that persist over the week (e.g., through deteriorated sleep quality) in Column (10).

Table C1 illustrates that our estimates are robust to more or less stringent FEs, assuring that our results cannot be explained by specific unobserved seasonal or regional patterns. Table C2 also indicates that our results are robust to different clustering choices, including two-way clustering by both monitoring stations and dates, to additionally account for possible spatial correlation or choosing broader geographical areas (46 prefectures) than simply pollen monitoring stations (120 stations). In addition, spatially clustered standard errors, as in Conley (1999), do not inflate our standard errors significantly (last three rows). Table C3 shows that our results are robust to the log-log specification or Poisson pseudo-maximum likelihood model to account for the count nature of the accidents.

#### Placebo

Table C4 falsely assigns the pollen levels of the exact day from the previous year or the next year in the same unit, finding much smaller and statistically insignificant results. Furthermore, Figure C2 displays the binscatter plots of the relationship between the logged daily pollen counts and the number of daily emergency ambulance transportation trips due to cancer.^23^ We find no discernable pattern as expected, which reassures us that our results are not driven by specific unobserved seasonal or regional patterns.

#### Replication

Traffic accidents are also separately recorded in police records, which include traffic accidents that result in personal injury and that are reported to the National Police Agency for the period 2019 to 2020 (see Appendix D for details on data). Table D1 compares the mortality estimates from traffic accidents using ambulance (our primary sample) and police records. While the estimate of the effect of pollen on mortality from traffic accidents using police records (*p*-value<0.01) is slightly larger than that using ambulance records, they are not statistically distinguishable from each other.^24^ We are reassured that the effect of pollen is robust to different samples collected by different government bureaus with potentially different definitions of traffic accidents, which strengthens the internal validity of our estimates.

### 5.4. Heterogeneity

#### Severity

Figure 7 plots the estimates along with a 95% confidence interval for each severity level, separately for all accidents (Panel A) and for traffic accidents (Panel B). Panel A suggests that while the estimates become smaller as the severity level increases, all the estimates, including death/fatal, are positive and statistically significant. Panel B shows similar patterns for traffic accidents. Interestingly, the estimate relative to the mean is larger in death/fatal than any other severity level in the case of traffic accidents, suggesting that pollen exposure may lead to serious consequences.

**Figure 7.**
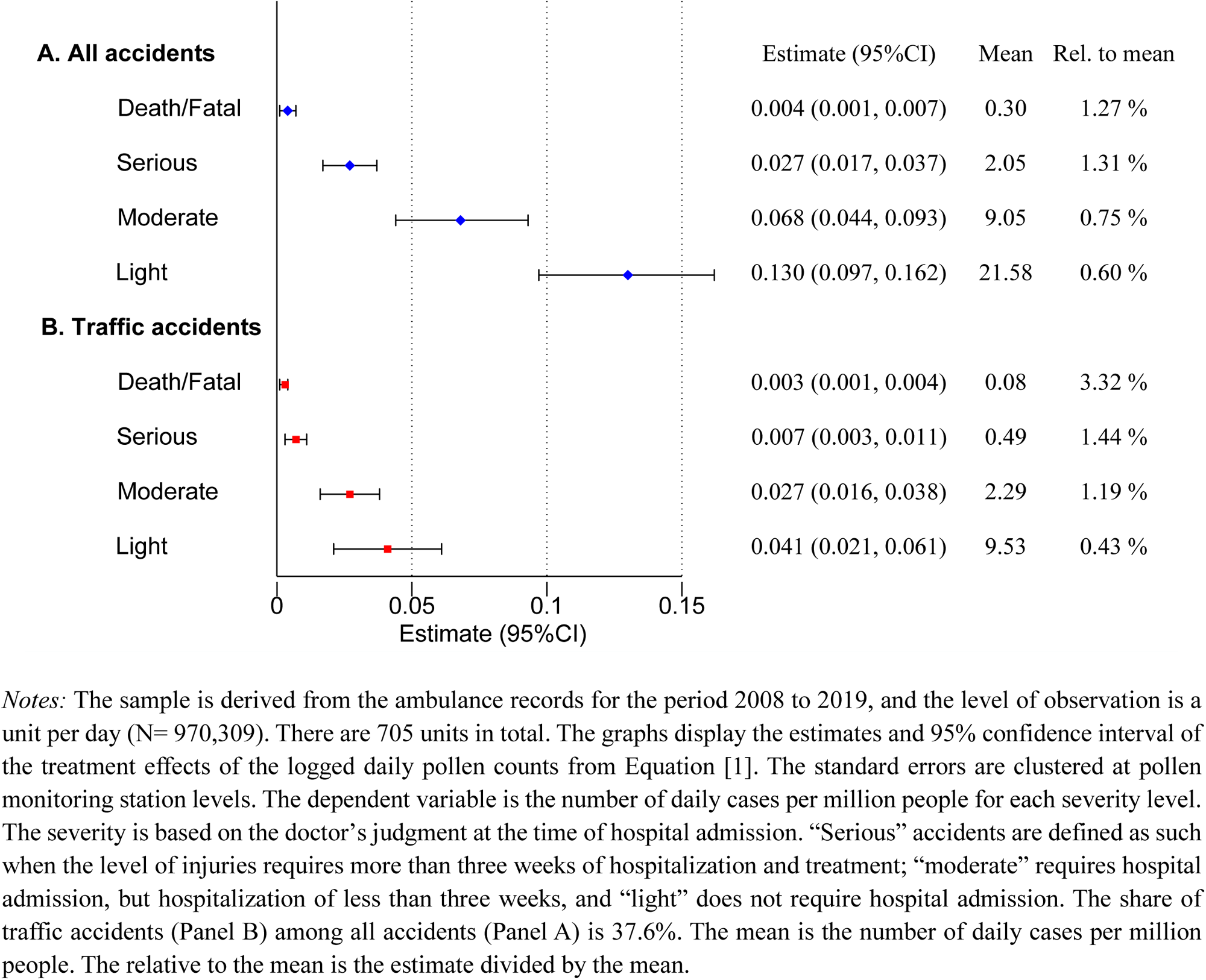
Treatment effects by severity.

#### Other heterogeneity

The ambulance records also include other characteristics of the accidents and people involved. Using all accident samples, Figure 8 explores the heterogeneous treatment effect other than severity.^25^ Panels A and B explore the heterogeneity by demographics, namely, age and gender. Overall, we find statistically significant effects in all age groups and both genders, with relatively similar magnitudes relative to the means, reported on the far right in the figure. The only exception is a slightly larger effect for the elderly (>65), even relative to the high baseline mean. This finding is in line with the elderly’s higher vulnerability to environmental externalities such as heat-mortality (Carleton and Hsiang 2016), cold temperature-mortality (Cohen and Dechezleprêtre 2022), and pollution-mortality (Jia and Ku 2019; Barwick et al. 2020) relationships.

**Figure 8.**
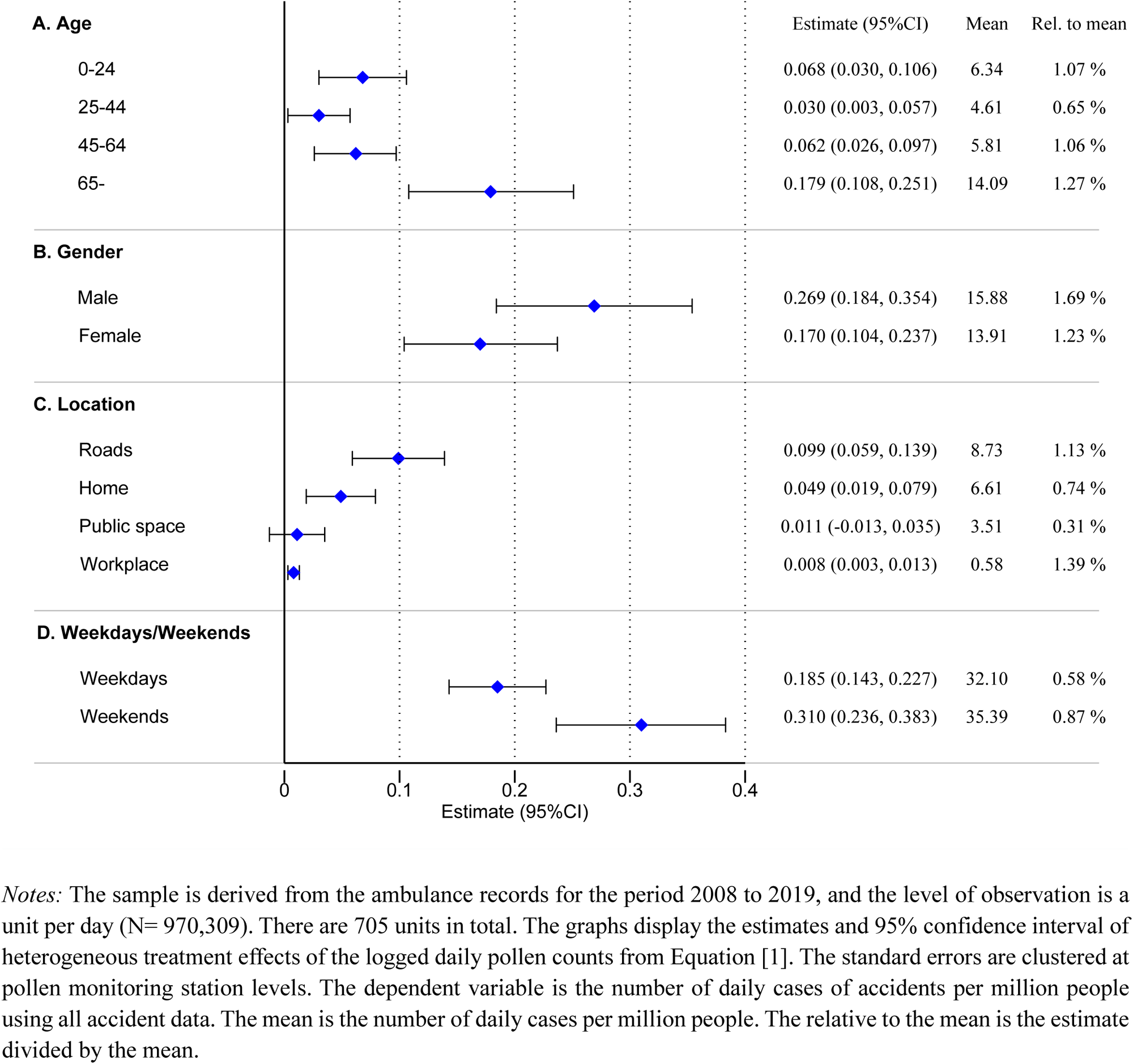
Other heterogeneous treatment effects (All accidents)

Panel C examines the heterogeneity by location of accident occurrence. The increase in the occurrence of accidents even takes place when individuals are at home, indicating how difficult it is for individuals to completely avoid outdoor pollen as the grains can remain on clothing (e.g., woolen coats) and are easily brought indoors. This may also reflect the lingering effect of outdoor pollen exposure. Finally, Panel D shows that the effect of pollen is larger on weekends than on weekdays. On the one hand, most individuals should have more discretion to remain indoors and avoid exposure on weekends, as non-mandatory trips can be canceled or rescheduled. On the other hand, those who go out on weekends might be less experienced drivers who do not usually drive for work during weekdays, or they travel to unfamiliar locations for leisure, making them potentially more subject to the elevated risk of pollen exposure. Our results suggest that the latter seems to dominate the former in this setting.

## 6. Short-term avoidance behaviors

Since both forecasts and real-time pollen information are commonly reported via television, newspapers, and various mobile phone apps during our sample period, people have ample information and time to adopt avoidance behaviors if they take the cognitive risk of pollen exposure seriously. A few low-cost and effective ways to mitigate or avoid the risk of temporal exposure to pollen are often mentioned on television and other media. These include wearing particulate-filtering masks and glasses, washing hands, avoiding wearing clothes that easily attract pollen, taking medications, and avoiding going out (Japan Society of Immunology and Allergology in Otolaryngology 2021).

### 6.1. Conceptual framework

Here, we provide a simple framework to consider the role of avoidance behaviors. Let us assume that *Accidents = f(Sick,Avoid)*, where the number of accidents is the function of the sickness level (*Sick*) and avoidance behaviors (*Avoid*). Since sickness level is influenced by the ambient pollen concentration (*Pollen*) and avoidance behaviors, that is, *Sick = g*(*Pollen, Avoid*),^26^ substituting it yields the following equation.

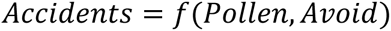

Then, the total derivative can be written as follows (Moretti and Neidell 2011; Neidell 2009):

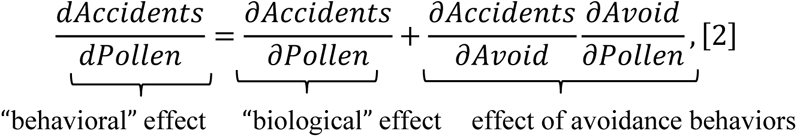

where the “behavioral” effect (what we estimate so far) of pollen on accidents consists of the “biological” effect of pollen (the first component of the right-hand side (RHS) variable) *and* the effect of avoidance behaviors (the second component of the RHS variable). The latter is the product of the marginal return from avoidance behaviors (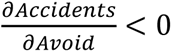), and the magnitude of avoidance behaviors in response to the level of pollen (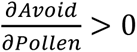). As the second component of the RHS variable is supposed to be negative, the total derivative that already incorporates avoidance behaviors is *smaller* (i.e., underestimated) than the desired partial derivative.^27^

Among the many avoidance behaviors, we specifically examine two types that can be observed through existing data, as complete data on these behaviors are usually unavailable (Deschênes et al. 2017). First, using in-home scanner data, we examine the purchase of products that protect against seasonal allergies, such as medications and masks. Second, using geolocation data, we examine whether people, including allergy sufferers, curtail outdoor activities, which mainly reduces the risk of outdoor accidents. Further, staying indoors limits possible pollen infiltration and, hence, simply averts the appearance of symptoms; as pollen grains are relatively large (≈30 *εm*) compared to much smaller particles, such as PM2.5, they are less likely to be able to enter houses if windows and doors are properly closed. Below, we investigate the extent of these two types of selective avoidance behaviors (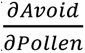) in our context.^28^

### 6.2. Purchase of allergy products

#### Data

We use in-home scanner data (called “Quick Purchase Report” (QPR)) provided by Macromill, Inc, a marketing company that owns one of the largest research panels on consumer purchase behaviors in Japan (Kuroda 2022). QPR collects data from roughly 30,000 monitors to construct a nationally representative panel dataset.^29^ The monitors scan all barcoded items they purchase every day. The data report the name and code, price, and quantity of the products purchased. They also contain some demographic information about the monitors, such as ZIP codes, age, gender, family structure, and income category, which are annually updated.

We extract the purchase records on three items, namely, allergy-related medications, allergy-related eye drops, and masks.^30^ The data include over 28 million individual-day observations, including days with no purchases, from February to May for the period 2012 to 2019. To account for the fact that these products can be easily stockpiled, we collapse data at an individual-week level.^31^ See Table E1 for summary statistics for the sample.

A few limitations of this dataset should be noted. We observe the purchases of over-the-counter (OCT) medications, but not prescription medications. Second, some people may purchase these goods in advance before the pollen season starts or use leftover medications from previous seasons. Therefore, the consumption expenditure measured here provides the lower bound of the actual expenditure on allergy products. Finally, we have no information on actual usage, which is a common issue for any in-home scanner data.

#### Results

Figure E1 displays the binscatter plots of the relationship between the logged average daily pollen counts and the weekly spending per person (in 10^-3^ $) for all allergy products in Panel A and for individual products in Panels B to D: medications (Panel B), eye drops (Panel C), and masks (Panel D). All figures display a linear relationship with the logged pollen counts, suggesting that people are more likely to purchase defensive products as the pollen concentration exacerbates.

Table 4 reports the estimate from the variant of Equation [1], modified to the weekly level data. Column (1) shows that a 10% increase in pollen counts leads to 4.40 (in 10^-3^ $) additional weekly spending on allergy products. On the national level, this translates to $9.6 million (= 4.40×10^-3^×120/7×127.4 million) per season, where 120 days comprise a typical pollen season, and 127.4 million is the total population of Japan. Columns (2)–(4), which describe individual products, show that the largest increase (relative to mean) comes from medication purchases.^32^

**Table 4.** Avoidance behaviors: Purchase of allergy products.

|  | **A. Total** | **B. By category** |  |  |
| --- | --- | --- | --- | --- |
|  |  | Medications | Eye drops | Masks |
|  | (1) | (2) | (3) | (4) |
| ln(pollen counts) | 44.002***<br>(2.545) | 24.521***<br>(1.428) | 13.143***<br>(0.969) | 6.429***<br>(0.824) |
| R-squared | 0.006 | 0.004 | 0.003 | 0.002 |
| N | 4,303,417 | 4,303,417 | 4,303,417 | 4,303,417 |
| N of individuals | 70,795 | 70,795 | 70,795 | 70,795 |
| Mean of dep. var (in 10<sup>-3</sup> \$) | 295.78 | 109.94 | 103.41 | 82.52 |
| *Share* | *100%* | *37%* | *35%* | *28%* |
| Municipality FE | X | X | X | X |
| Year-prefecture FE | X | X | X | X |
| Week FE | X | X | X | X |

*Notes:* The sample is derived from the in-home scanner data for the period from February to May for the years 2012 to 2019, and the level of observation is an individual per week (N= 4,303,417). The dependent variable is the weekly spending (in 10<sup>-3</sup> \$) for each allergy product. The estimates from the variant of Equation [1] are reported along with the standard errors clustered at pollen monitoring station levels in parentheses. In addition to the FEs in the table, we include weather covariates (precipitation, temperature, wind speed) and darkness. Significance levels: \*\*\* $p < 0.01$ , \*\* $p < 0.05$ , \* $p < 0.10$ .

#### Supplementary analysis

We also complement the above analysis by using the Google Trends/Twitter data that include the keywords “mask,” “air purifier,” and leading brand names of allergy medications in Japan, to study whether people seek information on specific protective products. Previous studies document that these searches are closely linked to actual purchases (Goel et al. 2010). Compared to the purchase data, these measures are more likely to capture contemporaneous behavior that should be more directly affected by the daily fluctuation of pollen counts. We are reassured by the fact that Figure F1 shows a strong relationship between pollen counts and both Google Trends data and the number of tweets on these keywords.

### 6.3. Avoidance of going out

#### Data

We use geolocation data (called “Mobile Spatial Statistics” (MSS)) provided by NTT DOCOMO, Inc., Japan’s largest mobile phone carrier. MSS provides the population estimate at a 500×500-meter mesh at an hourly level across Japan, based on the location information of 85 million users of NTT DOCOMO (as of March 2022) from a total population of 127 million for Japan (Terada et al. 2013).^33^ While physical mobility data has received significant attention in the social sciences since the onset of the COVID-19 pandemic (e.g., Google’s COVID-19 Community Mobility Reports), such data are not yet widely used to examine avoidance behaviors or environmental stressors (see Burk et al. 2022).

Our mobility measures capture the estimated population of a mesh with the largest number of establishments in the customer service industry in the municipality to capture bustling areas (e.g., business districts and shopping and dining areas), which are more likely to represent the population engaging in outdoor activities. We use the estimated population at 2 pm, because the daily population in commercial areas tends to peak around this time (Seike et al. 2015). We collapse the estimated population at the unit level by taking the average of all municipalities in the unit for the period from February 2014 to May 2019. We treat this measure as a proxy for engaging in outdoor activities (“outdoor population,” hereafter). See Appendix G for details of the data construction of our mobility measures.^34^

Before examining the relationship between pollen load and mobility measures, we attempt to verify that curtailing outdoor activities is effective in reducing the risk of accidents.^35^ We regress the number of accidents (our main outcome) on our mobility measure with the same sets of FEs and controls as Equation [1] (excluding a logged number of pollen counts). Table E1 shows that outdoor population is highly positively correlated with the number of accidents that occur, reassuring us that simply reducing outdoor activities can be a meaningful means to mitigate the risk of accidents, that is, (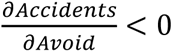).

#### Results

Figure G1 displays the binscatter plots of the relationship between the logged pollen counts and the logged outdoor population. Panel A shows no clear relationship between pollen counts and daytime outdoor population on average. Because most people should have more discretion to stay indoors on weekends than on weekdays, Panel B plots the same relationship for weekends only. Here, we find some negative relationships, suggesting that individuals seem to engage in some avoidance behaviors on weekends. However, as discussed below, the magnitude of avoidance is quite small.

Table 5 reports the estimate from Equation [1], where the outcome is the logged outdoor population. Columns (1) and (2) show that the estimates are negligible and far from statistically significant for all days and weekdays. Column (3) for weekends shows the elasticity of outdoor population with respect to pollen counts is as low as –0.0021 (*p*-value<0.01), implying that a 10% increase in pollen concentration leads to only a 0.021% reduction in the outdoor population. Table 5 also reports the estimates on four indicators for rainfall (base: no rainfall) for comparison. The outdoor population decreases as rain intensity increases, confirming that our mobility measure adequately captures individuals’ decision-making processes in relation to engaging in outdoor activities.^36^ The estimates from Column (3) indicate that rainfall between 1mm and 2mm reduces the outdoor population by 9.9 times more than a 1% increase in the pollen count.

**Table 5.** Avoidance behaviors: Refrain from going out.

Outcome: logged outdoor population
|  | A. All | B. By type of day |  |
| --- | --- | --- | --- |
|  |  | Weekdays | Weekends |
|  | (1) | (2) | (3) |
| ln(pollen counts) | -0.0005<br>(0.0006) | 0.0000<br>(0.0006) | -0.0021***<br>(0.0007) |
| Rainfall (base: no rainfall) |  |  |  |
| <1 mm | -0.0061***<br>(0.0013) | -0.0059***<br>(0.0011) | -0.0047**<br>(0.0019) |
| 1 mm ≤ & < 2 mm | -0.0167***<br>(0.0035) | -0.0144***<br>(0.0024) | -0.0208***<br>(0.0080) |
| ≥ 2 mm | -0.0167***<br>(0.0045) | -0.0150***<br>(0.0041) | -0.0249***<br>(0.0082) |
| R-squared | 0.98 | 0.99 | 0.99 |
| N | 478,853 | 343,454 | 135,399 |
| Unit FE | X | X | X |
| Day-of-week FE | X | X | X |
| Month-year FE | X | X | X |
| Month-prefecture FE | X | X | X |

### 6.4. Policy implications

Our analysis reveals that people are engaging in some avoidance behaviors. Hence, one might argue that the present status of protective behavior reflects people’s revealed preferences that already incorporate the potential risk (Leard and Roth 2019) and, thus, there is no room for policy intervention. However, it is more natural to think that the current status that relies on individuals’ self-protection to mitigate pollen exposure and prevent accidents would be insufficient for the following reasons. First, to our knowledge, this is the first study to document the causal link between pollen exposure and the risk of accidents in a field setting. Hence, it is hard to believe that most people do fully understand such risk and consider it when making decisions. Second, the negative externality of accidents is unlikely to be incorporated into individual behaviors.

Since ample information on pollen is already provided in various forms in Japan, it is crucial to draw more attention to such information.^37^ For example, public information campaigns such as the “pollen alert”—which encourage decreasing outdoor activities, or recommend using public transportation on high pollen days due to the heightened risk—can be a low-cost and effective tool to reduce pollen-induced accidents. In fact, a few studies demonstrate that alerts for smog and ozone successfully induce people to take precautionary actions to reduce exposure (e.g., Anderson et al. 2022; Cutter and Neidell 2009). Considering the negative externality of accidents, the government can mandate firms to allow sick leave for seasonal allergies, or at least allow remote work for patients with severe seasonal allergies. Providing a subsidy for protective products or technology (e.g., allergen immunotherapy) is another option.

Finally, unlike “ex-post” government interventions, such as pollen alerts, “ex-ante” interventions, such as trimming pollen-emitting plants and replanting newly developed trees that emit less pollen (e.g., the Japanese cedar) to remove the source of pollen “production,” could be more cost-effective in the long-run. However, as implementing this process may take time, ex-post interventions, like the pollen alert, and aids for engaging in avoidance behavior, can be an effective temporary remedy.

## 7. Additional analysis

### 7.1. Medium-term adaptation

While our focus so far is limited to short-term outcomes, longer-term adaptation (either technological, behavioral, or biological) may take time. Recent medications for seasonal allergies cause less drowsiness and reduce the risk of unsafe driving. Table A1 lists medications for seasonal allergies released during the period 1994 to 2017 in Japan and shows that it is now less common for manufacturers to mention the medication’s effect on driving ability, through cautionary text, such as “Driving not allowed.” In fact, using in-home scanner data used above, Figure A4 shows that the share of spending on allergy medications that do not prohibit driving among total medication spending increased from 25.2% in 2012 to 45.6% in 2019.^38^ Individuals may also engage in defensive spending (e.g., buying air purifiers or receiving allergen immunotherapy) over a longer time span. Furthermore, unlike short-run physiological acclimatization, *behavioral* acclimatization may require more time to take effect (Graff Zivin and Neidell 2014).

Thus, we utilize data that spans a 12-year period from 2008 to 2019 to examine how the impact of pollen on accidents has changed over time. Figure 9 presents estimates of the effects of pollen separately for each year. While pollen-accident sensitivity becomes marginally smaller in the later periods compared to that in the earlier periods, the magnitude of the decline is slight, and none of the estimates in the later periods are statistically distinguishable from those in the earlier periods.

**Figure 9.**
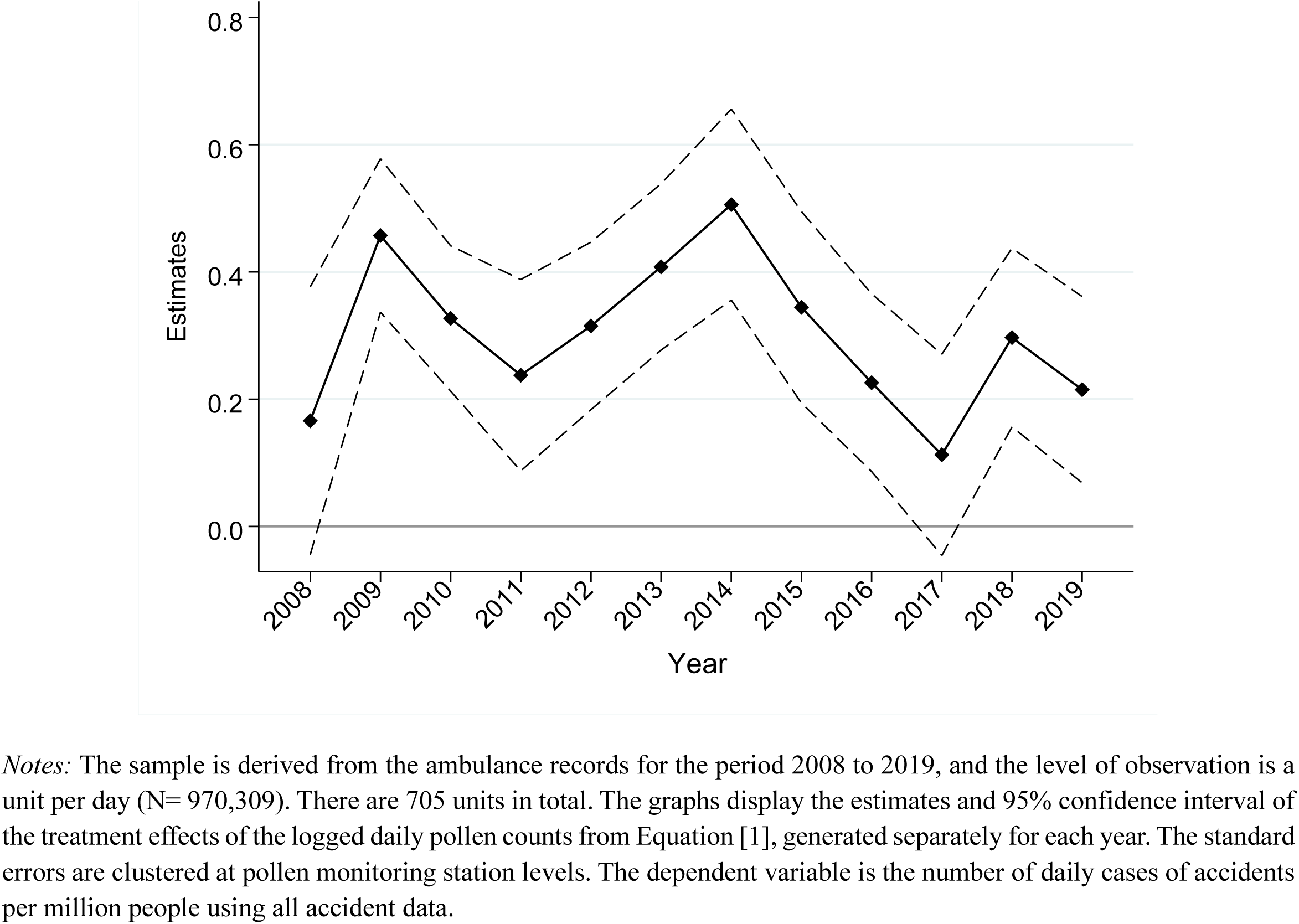
Effects of pollen over time.

We also divide 705 units into quantiles by the mean of pollen counts during the period 2008 to 2019 to investigate whether people who live in regions with a high pollen concentration, on average, experience smaller impacts of pollen concentrations than those who live in regions with a low pollen concentration through undertaking a series of longer-term adaptations.^39^ Figure C4 indicates that the estimates are reasonably similar across quantiles, again demonstrating limited longer-term adaptive potential. Overall, we fail to find compelling evidence of medium-term adaptation to the destructive impact of pollen exposure.

### 7.2. Projecting damages due to climate change

This section combines the estimates of the effect of pollen on accidents documented thus far with projections of future climate, as well as the relationship between the temperature and pollen counts obtained in Figure 1, to measure the magnitude of expected changes in the number of pollen-induced accidents.^40^

Table 6 summarizes the projected damages based on the “business as usual” scenario (RCP 8.5), which predicts that the summer temperature in Japan will increase by 4.1℃ in the years 2076 to 2095 (MEXT and JMA 2020). We calculate the number of additional pollen-induced accidents associated with this temperature change as follows: Based on the relationship between the summer temperature and pollen counts (Panel A of Figure 1), this increase in temperature could lead to additional daily pollen counts of 686 (= 167.4×4.1) grains/m^3^, which would correspond to a 0.529 increase in the logged pollen counts from the mean of 984 grains/m^3^. We then multiply the estimates from Panel A of Figure 6 for each severity level by 0.529, and then by 120 (days per pollen season), and then further by 127.4 (the total population) to calculate the additional annual accidents as is reported in Row (1). Row (1) indicates that a 4.1℃ increase in the average summer season temperature is expected to increase the pollen-induced death/fatal, serious, moderate, and light accidents by 30, 216, 541, and 1036, respectively, bringing the total number of additional annual accidents to 1,823.

**Table 6.**
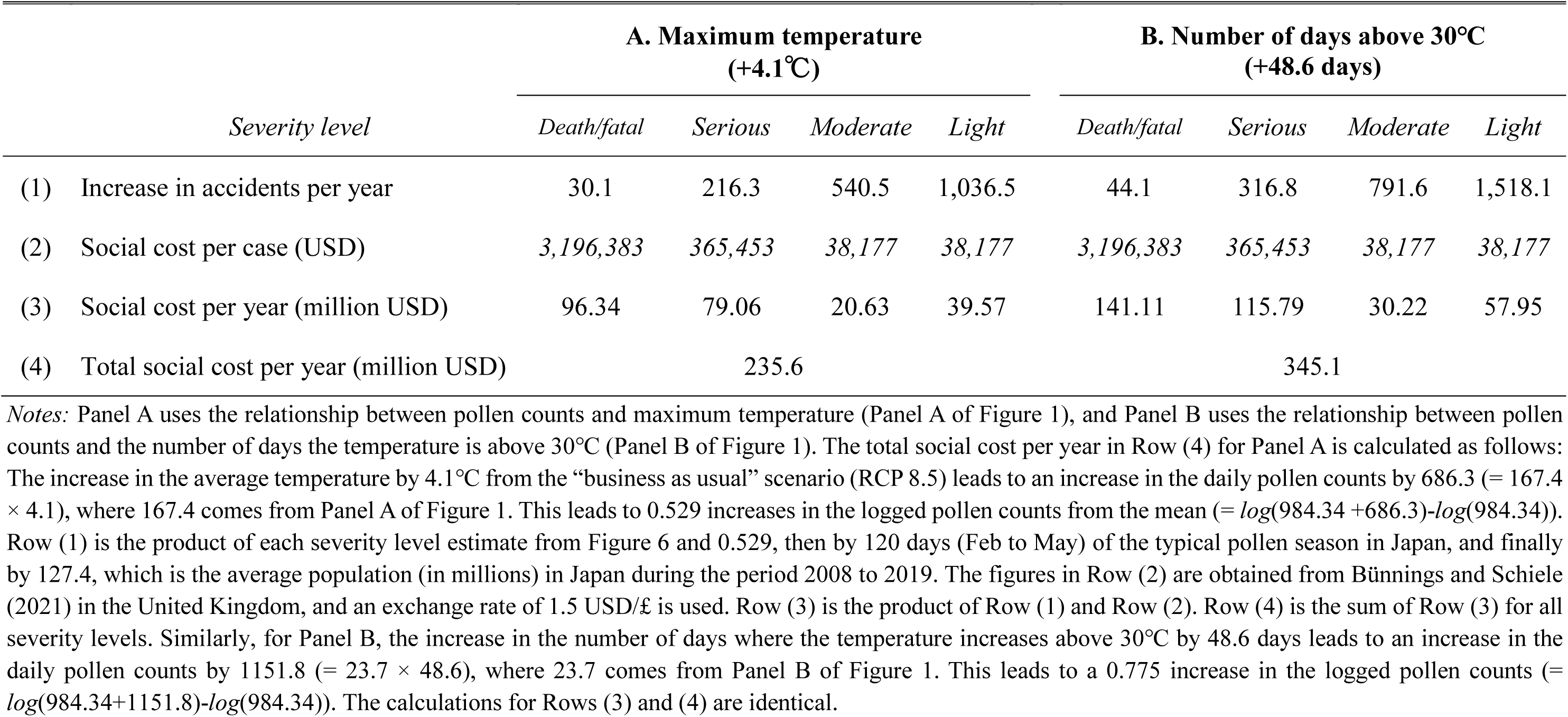
The impact of climate change from the “business as usual” scenario.

| *Severity level* | **A. Maximum temperature**<br>**(+4.1°C)** |  |  |  | **B. Number of days above 30°C**<br>**(+48.6 days)** |  |  |  |
| --- | --- | --- | --- | --- | --- | --- | --- | --- |
|  | *Death/fatal* | *Serious* | *Moderate* | *Light* | *Death/fatal* | *Serious* | *Moderate* | *Light* |
| (1) Increase in accidents per year | 30.1 | 216.3 | 540.5 | 1,036.5 | 44.1 | 316.8 | 791.6 | 1,518.1 |
| (2) Social cost per case (USD) | 3,196,383 | 365,453 | 38,177 | 38,177 | 3,196,383 | 365,453 | 38,177 | 38,177 |
| (3) Social cost per year (million USD) | 96.34 | 79.06 | 20.63 | 39.57 | 141.11 | 115.79 | 30.22 | 57.95 |
| (4) Total social cost per year (million USD) | 235.6 |  |  |  | 345.1 |  |  |  |

We then convert these additional accidents into monetary terms by multiplying the resulting accident counts in Row (1) by the average accident costs from Bünnings and Schiele (2021) in the United Kingdom, as is reported in Row (2).^41^ Row (3) demonstrates that pollen-induced accidents resulting in death/fatal, serious, moderate, and light injuries correspond to actual monetary costs of $96.3 million, $79.1 million, $20.6 million, and $39.6 million, respectively. This equates to a total annual social cost of $236 million, which can be seen in Row (4). Interestingly, this figure far exceeds the budget for the Japanese Forestry Agency’s pollen reduction program, which is currently $1.1 million (Forest Agency 2021).^42^

We note that our simple damage projection calculation probably yields a conservative estimate because (i) we do not taken into account any extension of the pollen season due to the increasingly hotter summer seasons, (ii) we only consider approximately four months of high pollen season, (iii) we are unable to take into account the accidents that occur at both sides of the severity distribution spectrum (i.e., minor cases that do not require ambulance transportation to hospitals and immediate deaths), and (iv) we make use of the relatively conservative value of a statistical life (Bünnings and Schiele 2021) rather than a more commonly utilized value.^43^

Finally, to the extent that spending on allergy products can be considered defensive spending rather than just another health expense (i.e., transfer from individuals to firms), such spending should be included in the social cost (Deschênes et al. 2017). Based on the computation in Section 6.2, a 0.529 (instead of 0.1) increase in the logged pollen counts due to elevated temperature leads to $50.8 million (= $9.6 million×0.529/0.1) additional spending on allergy products. This figure is not trivial compared to the $236 million associated with pollen-induced accidents calculated above, suggesting the empirical importance of defensive spending. Note again that the cost of allergy products includes that of OCT drugs only.

## 8. Conclusion

This study is the first assessment of the impact of pollen exposure on the likelihood of accidents using Japanese archived ambulance records for the period 2008 to 2019. We find that exposure to elevated pollen levels increases the occurrence of all types of accidents. Our results are consistent with those of established, clinically based studies that have documented the adverse cognitive effects of pollen exposure, the long-term implications of which have not previously been assessed in the real-world, except for children’s academic performance. In addition, as we find the effects of pollen exposure to be broadly uniform across a range of observed demographics, locations, and calendar months, and find limited evidence of changes in the treatment effects over time, the underlying mechanism linking pollen exposure to accidents may be generalizable across different contexts.

Our analysis of internet search activities and social media posts for pollen-related topics suggests that individuals seem to have a good awareness of daily pollen levels. However, while some people obviously take measures to alleviate their allergy symptoms by purchasing available technologies, the very limited avoidance behaviors, determined from geolocation data, imply that they lack awareness of or underestimate the cognitive risk of pollen exposure. Thus, the current policy that relies on individuals’ self-protection to mitigate pollen exposure may only have limited benefits. Governmental efforts to raise public awareness of the risks of pollen exposure and promote widespread behavioral change might be necessary.

Finally, it is imperative to reiterate that the estimated social cost we establish might only reflect the *tip of the iceberg* of the potentially extensive social cost associated with increasing pollen counts. While we remain agnostic about the underlying mechanisms, to the extent that exposure to pollen impairs cognitive function, any daily human activity that requires normal cognitive performance and decision-making abilities can be affected. Thus, it is vital to quantify the potential damage in order to acquire a better understanding of the full costs of pollen concentration.

## Data Availability

The main data (accident records) used in the paper is administrative data in Japan. Because of confidentiality agreements with the government that provided the data, we are not allowed to make this data available to the public. However, any researcher can easily have access to all the datasets by simply applying to the following addresses.
Ambulance records
Data description: ambulance records archive
Source: Fire and Disaster Management Agency (FDMA) of the Ministry of Internal Affairs and Communications
https://www.fdma.go.jp/en/post1.html
Police records
Data description: traffic accident records of accidents that involve injuries or deaths
Source: National Policy Agency (NPA)
https://www.npa.go.jp/publications/statistics/koutsuu/opendata/index_opendata.html
Pollen
Data description: Hourly pollen counts from 120 stations, as well as hourly rainfall, temperature, wind speed, and wind direction from nearby weather stations during the pollen season (February to May except for Hokkaido, where the pollen season is March to June).
Source: Ministry of the Environment (MOE), Pollen Monitoring System (Hanako-san)
https://tenki.jp/pollen/
Note: MOE terminated data collection of pollen counts in 2021.
Temperature
Data description: Hourly temperature (outside of the pollen season)
Source: Japan Automated Meteorological Data Acquisition System (AMeDAS)
operated by the Japan Meteorological Agency (JMA)
https://www.data.jma.go.jp/obd/stats/etrn/
Pollution
Data description: Hourly SO2, NO, NO2, CO, OX, PM10
Source: National Institute for Environmental Studies
https://www.nies.go.jp/igreen/index.html
Geolocation data
Data description: called Mobile Spatial Statistics data, which are estimates based on the location information of 85 million NTT DOCOMO cellphone users (as of March 2022)
Source: NTT DOCOMO, Inc
https://mobaku.jp/ (in Japanese)
In-home scanner data
Data description: called Quick Purchase Report, which is the daily panel of purchase records from roughly 30,000 monitors
Source: Macromill, Inc
https://www.macromill.com/service/digital-data/consumer-purchase-history-data/ (in Japanese)

## Appendix A: Additional Figures and Tables

**Figure A1.**
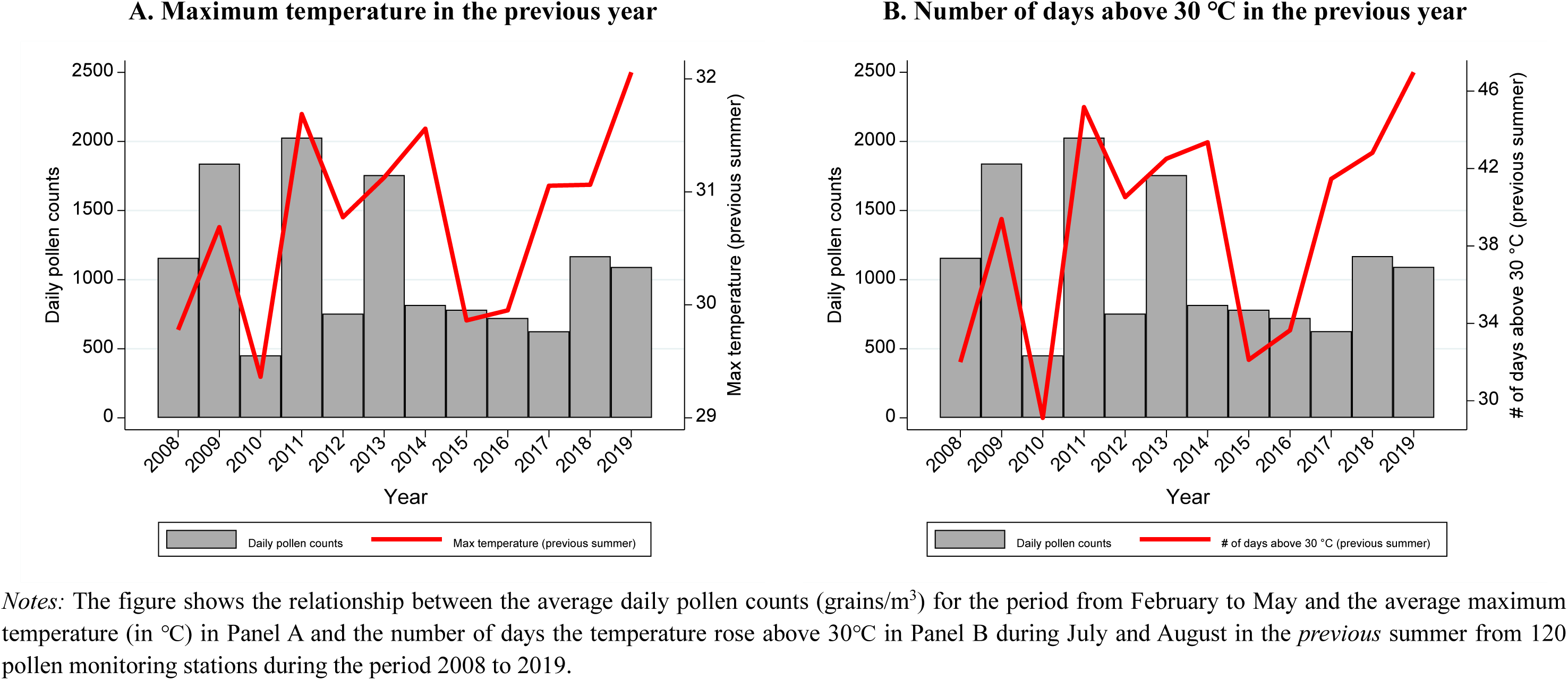
Times series of pollen counts and maximum temperature.

**Figure A2.**
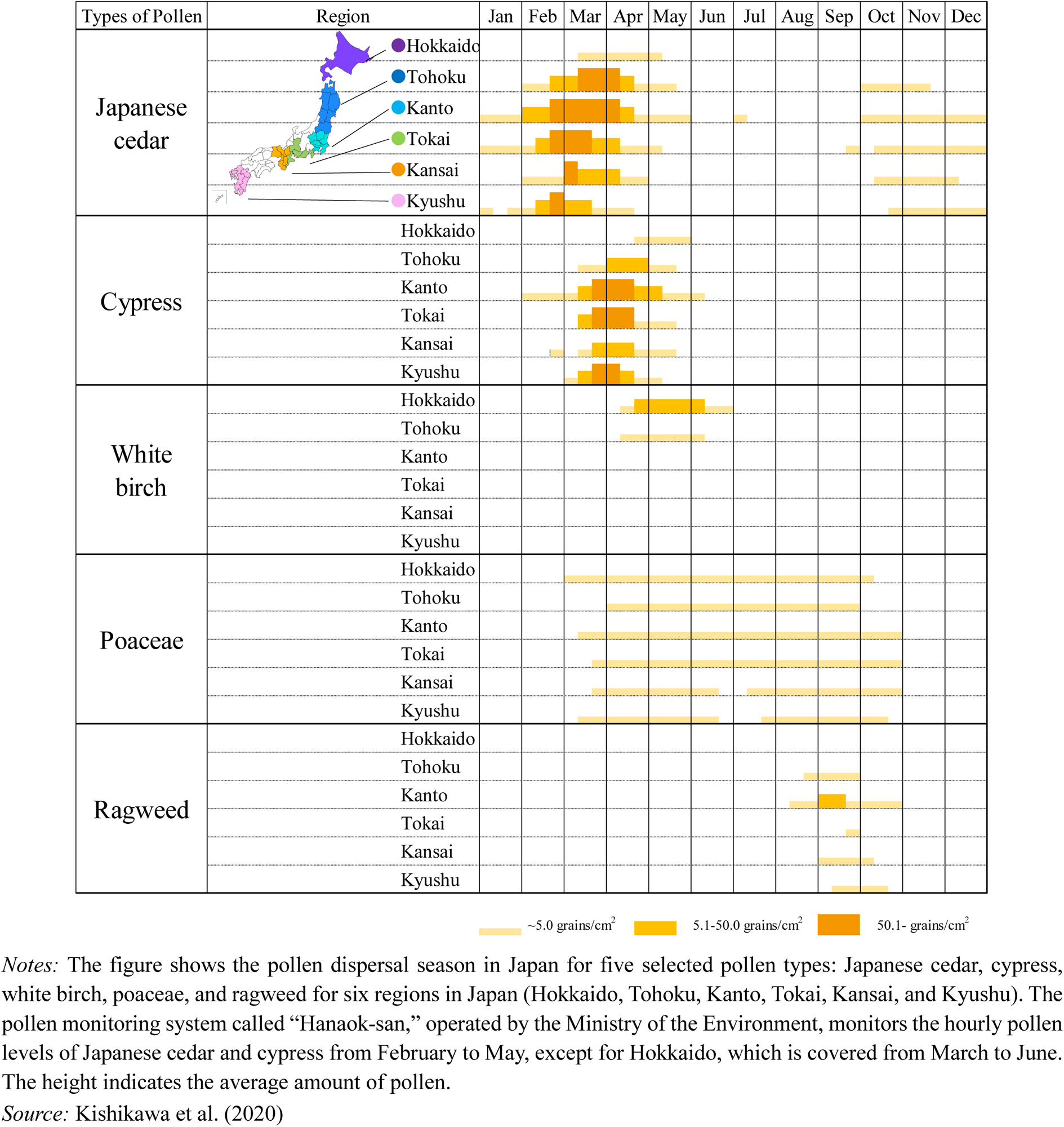
Calendar of pollen season in Japan.

**Figure A3.**
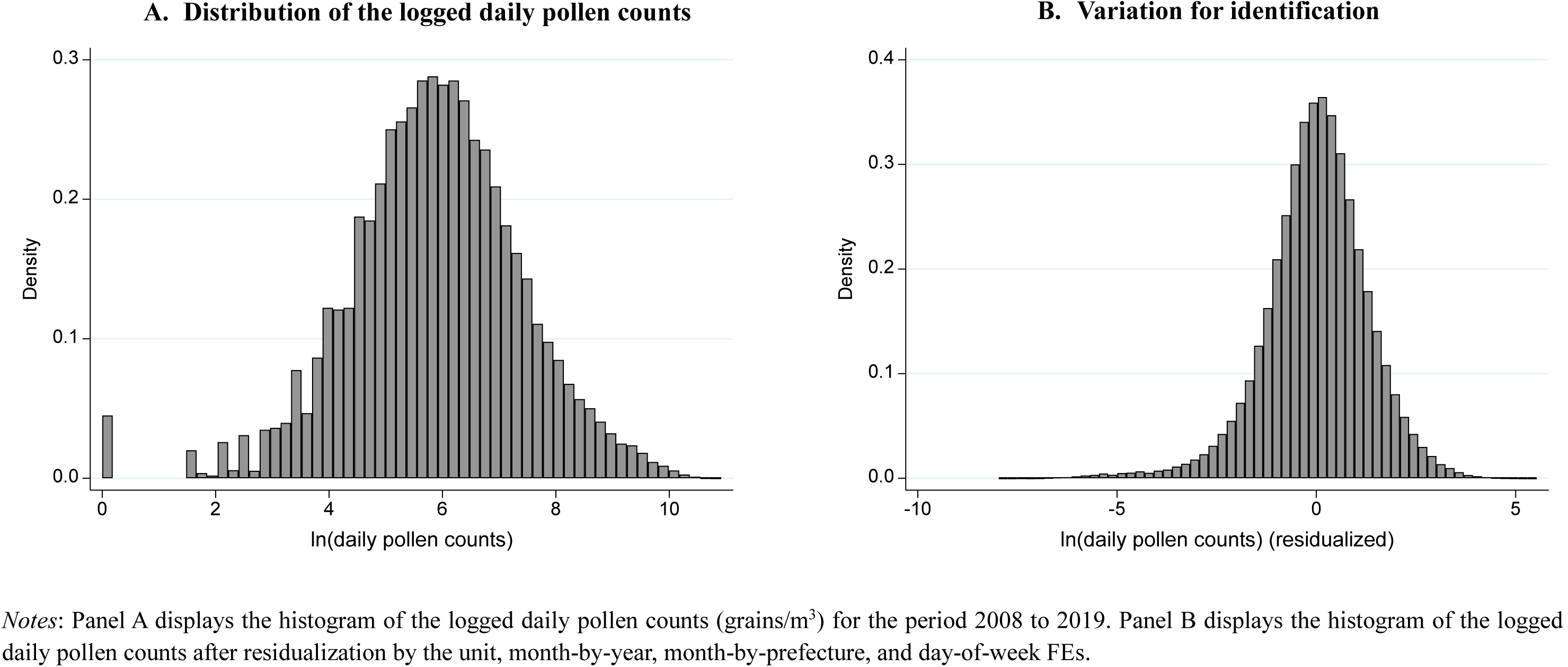
Variation in pollen counts.

**Figure A4.**
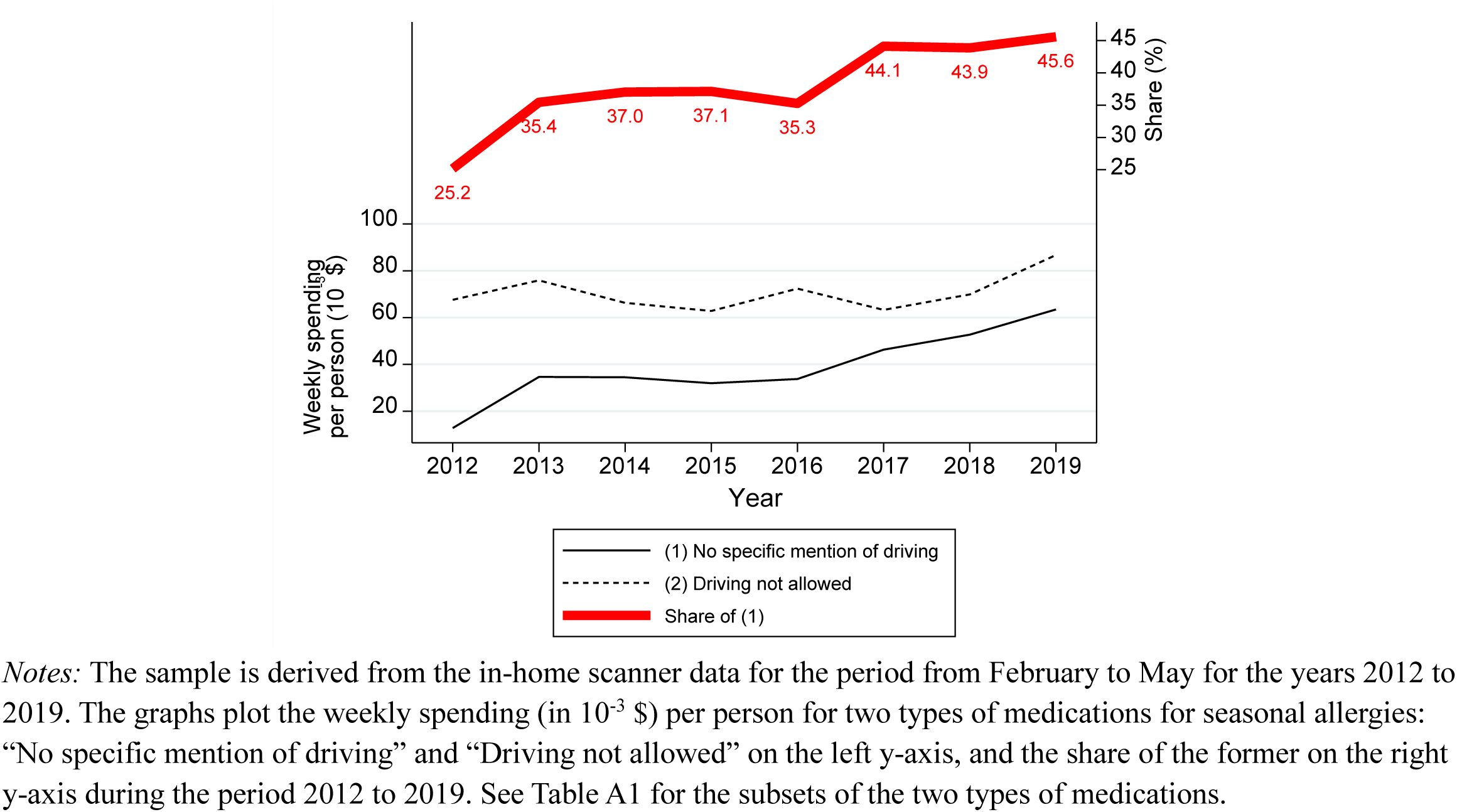
Time series of spending on medications for seasonal allergies (by type)

**Figure A5.**
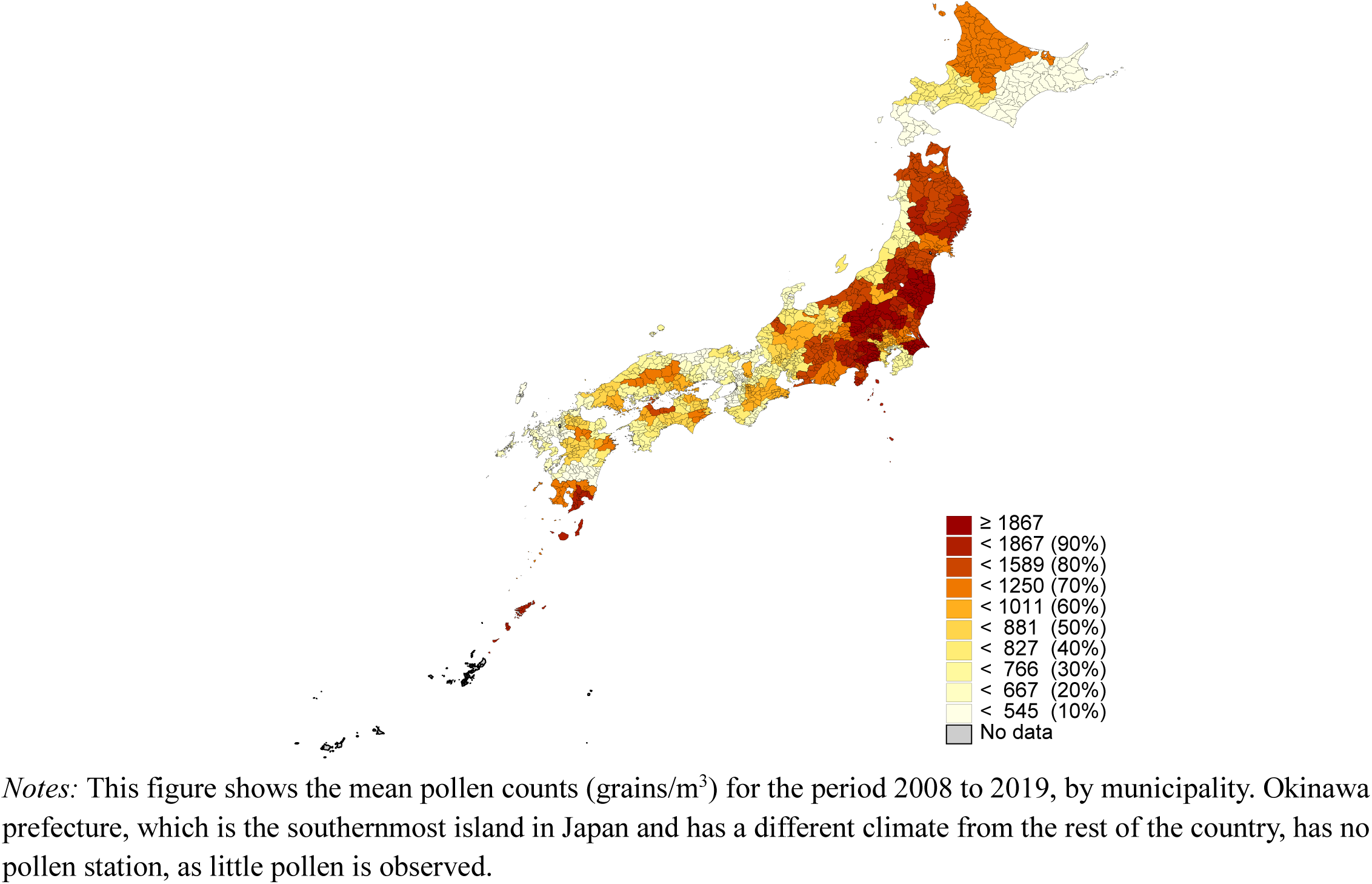
Spatial variation in pollen counts.

**Table A1.** List of medications for seasonal allergies.

| **Brand name**<br>**in English** | **Year of**<br>**release** | **Mention of driving** |
| --- | --- | --- |
| Alesion | 1994 | Driving not allowed |
| Evastel | 1996 | Driving not allowed |
| Zyrtec | 1998 | Driving not allowed |
| Talion | 2000 | Driving not allowed |
| Allegra | 2001 | No specific mention of driving |
| Allelock | 2001 | Driving not allowed |
| Claritin | 2002 | No specific mention of driving |
| Xyzal | 2010 | Driving not allowed |
| Dellegra | 2013 | No specific mention of driving |
| Bilanoa | 2016 | No specific mention of driving |
| Desalex | 2016 | No specific mention of driving |
| Rupafin | 2017 | Driving not allowed |

*Notes:* The table lists the brand names of allergy medications used to treat seasonal allergies together with the year of release of the medication as well as any particular mention of whether you are allowed to drive after taking the medication. The medications highlighted in gray do not include any specific mention of driving after taking same. Note that our primary data on accidents span 2008 to 2019.

## Appendix B: “First stage”—Symptoms of Seasonal Allergies

**Figure B1.**
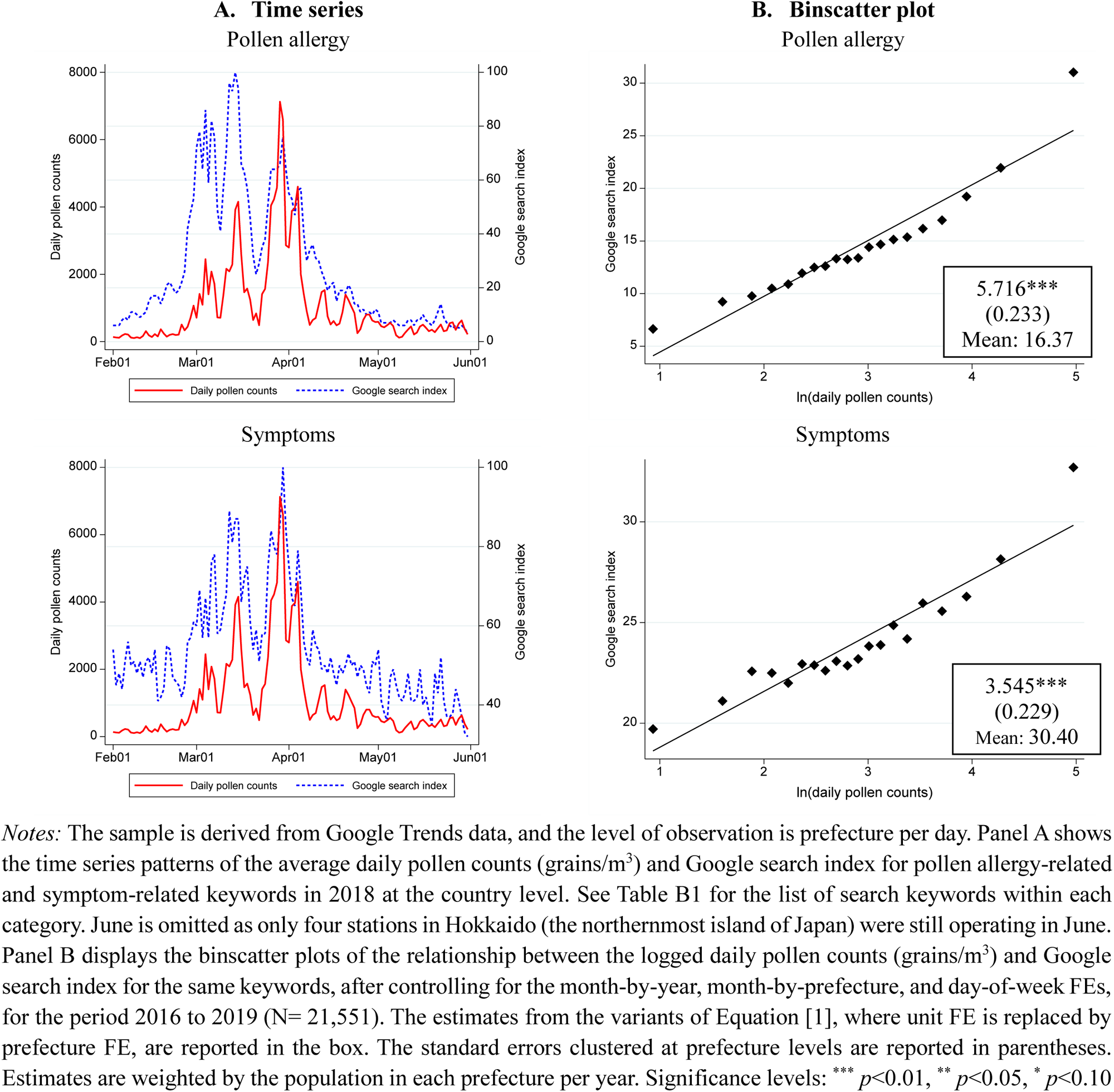
Daily pollen counts and Google Trend.

**Figure B2.**
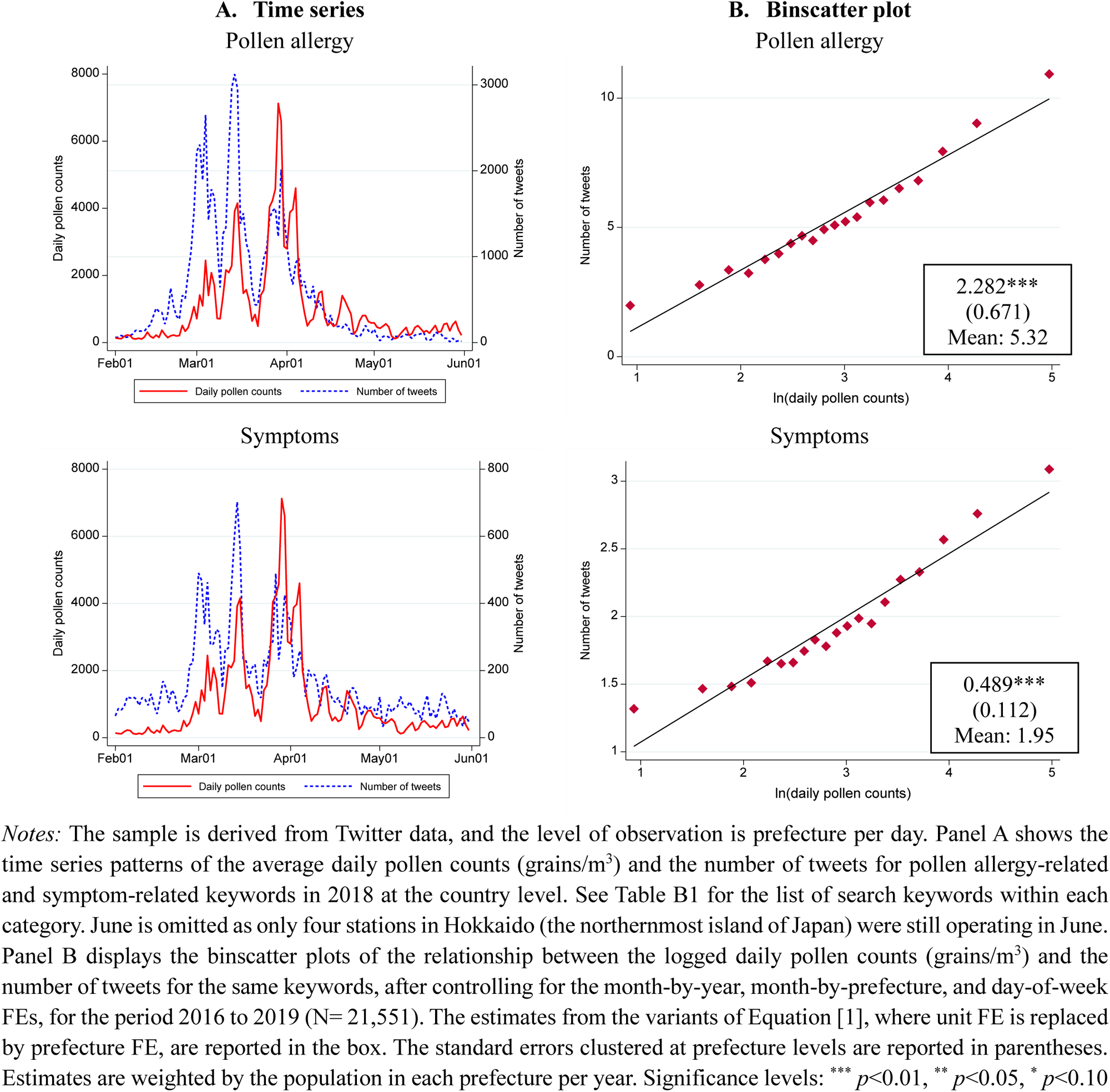
Daily pollen counts and tweets.

**Figure B3.**
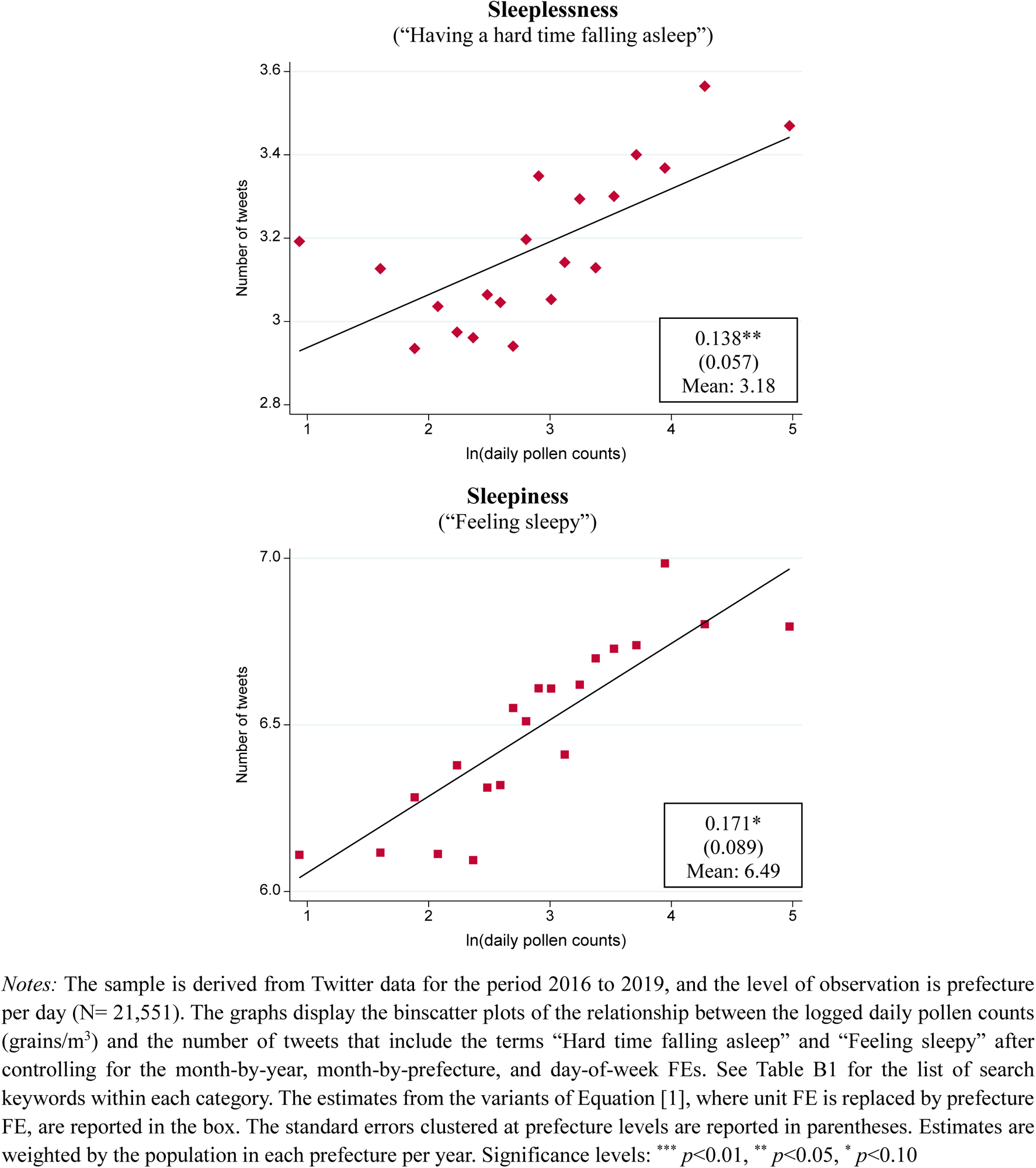
Binscatter plot of pollen counts and tweets: Sleep-related.

**Table B1.**
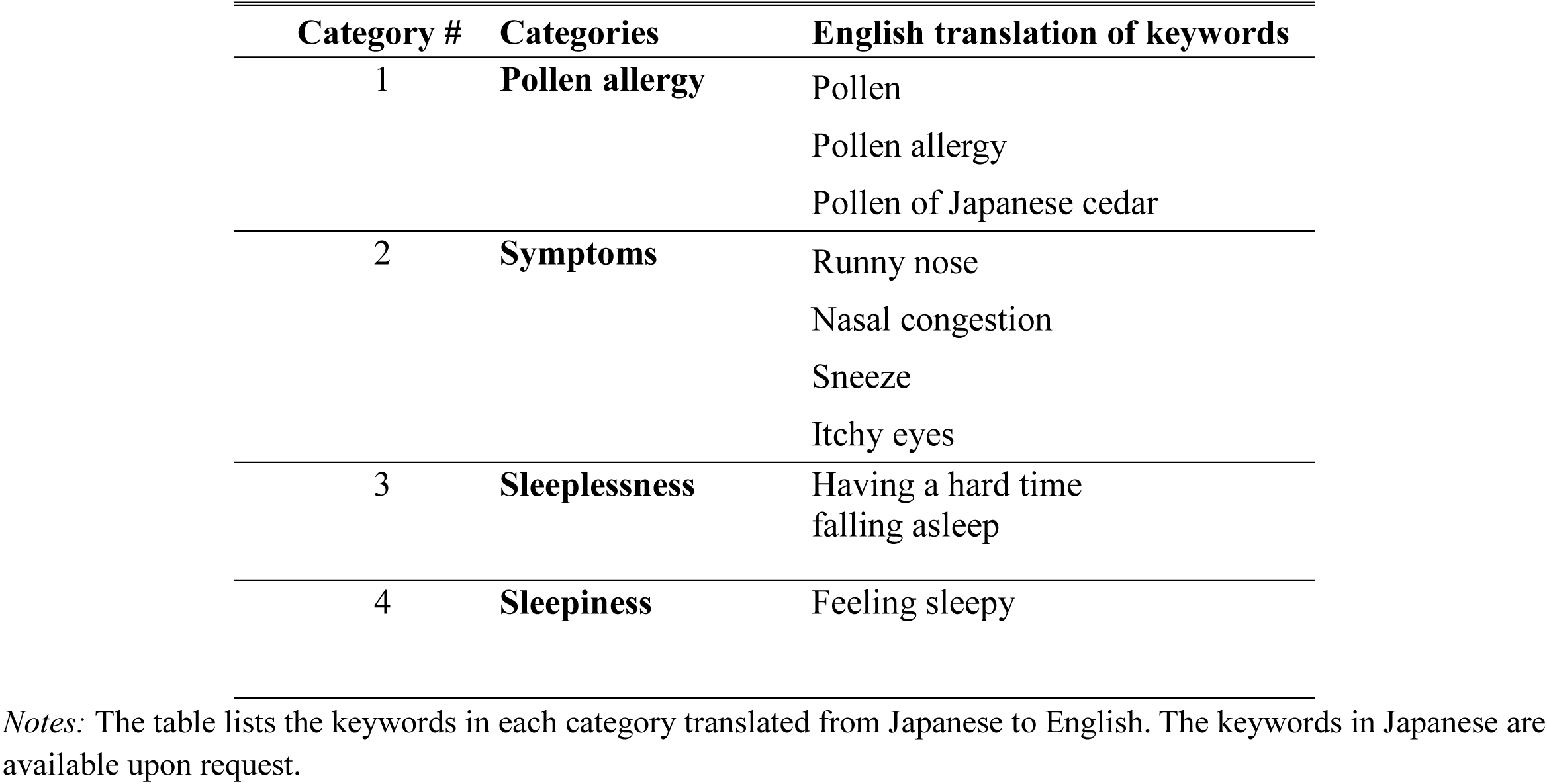
List of search keywords for symptoms.

## Appendix C: Ambulance Records

**Figure C1.**
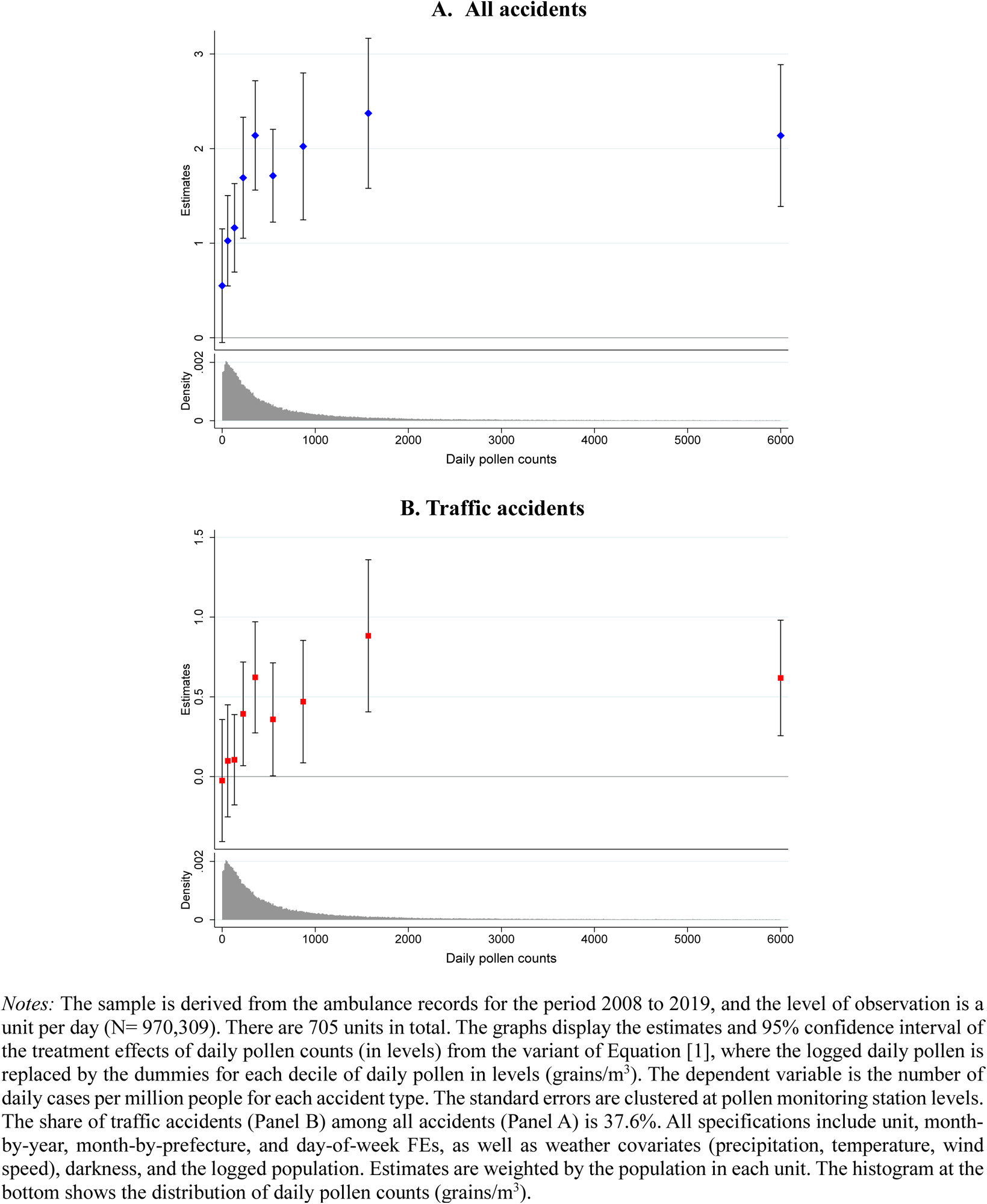
Dose responses.

**Figure C2.**
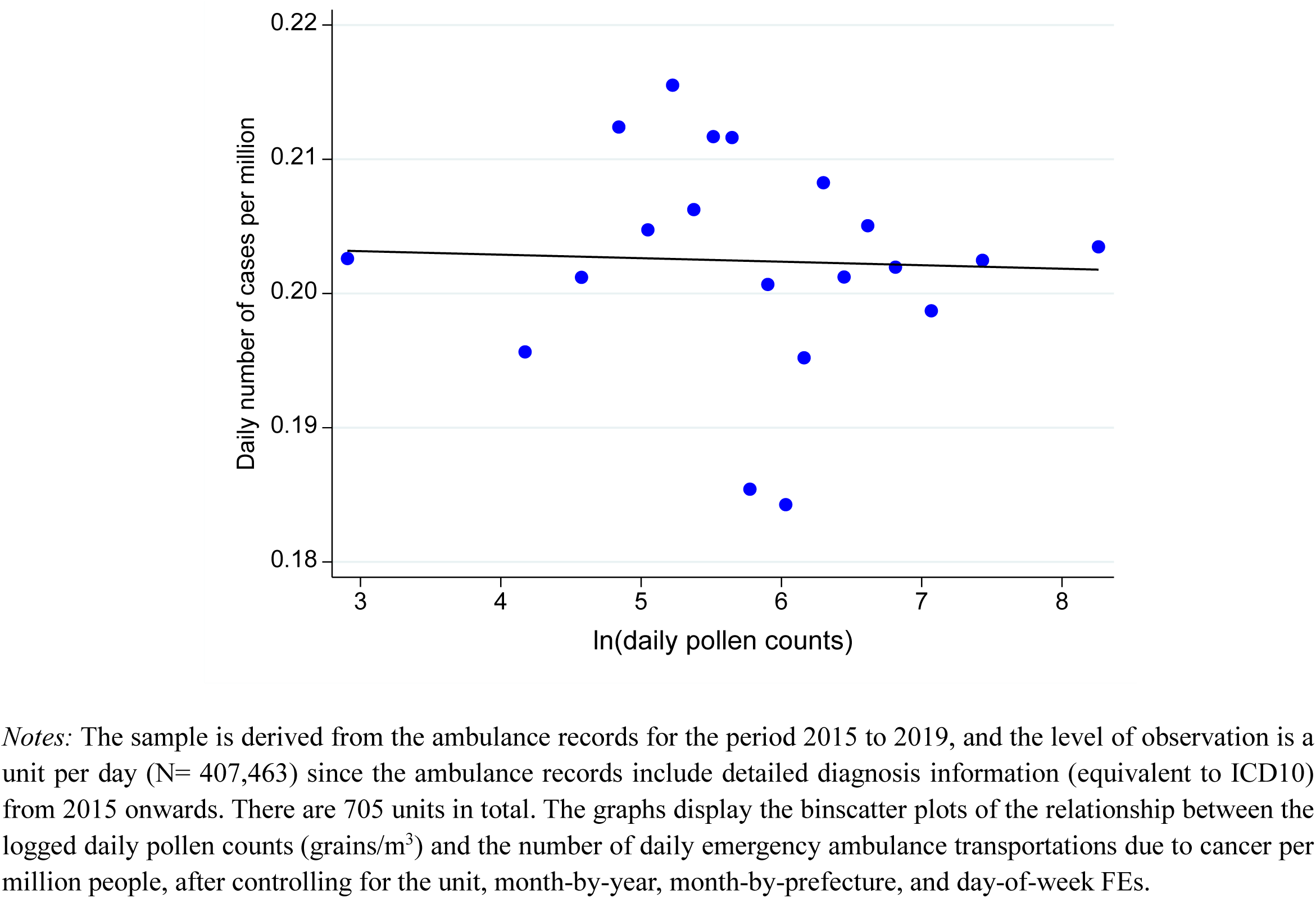
Pollen and emergency ambulance transportation due to cancer.

**Figure C3.**
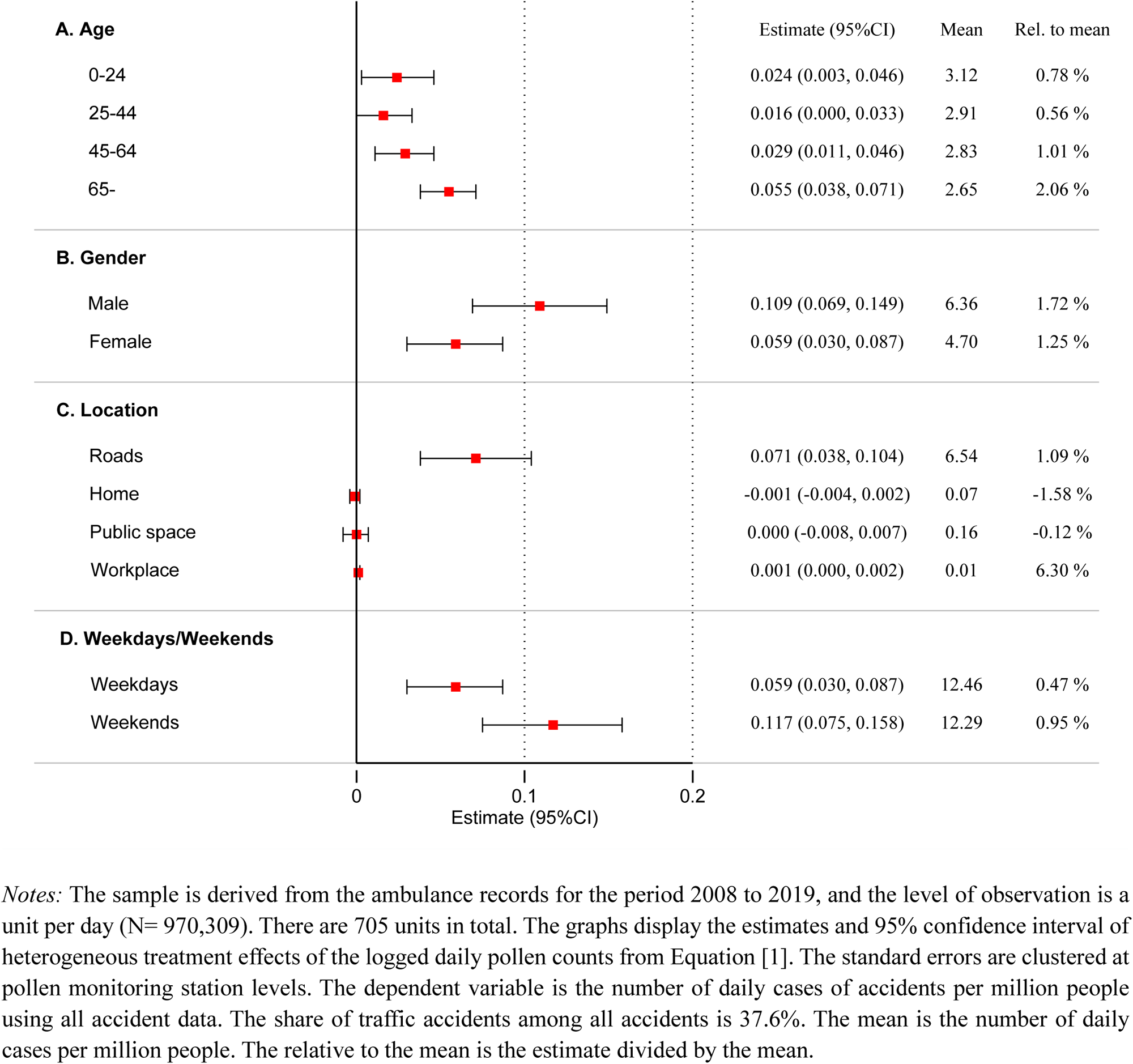
Other heterogeneous treatment effects (Traffic accidents)

**Figure C4.**
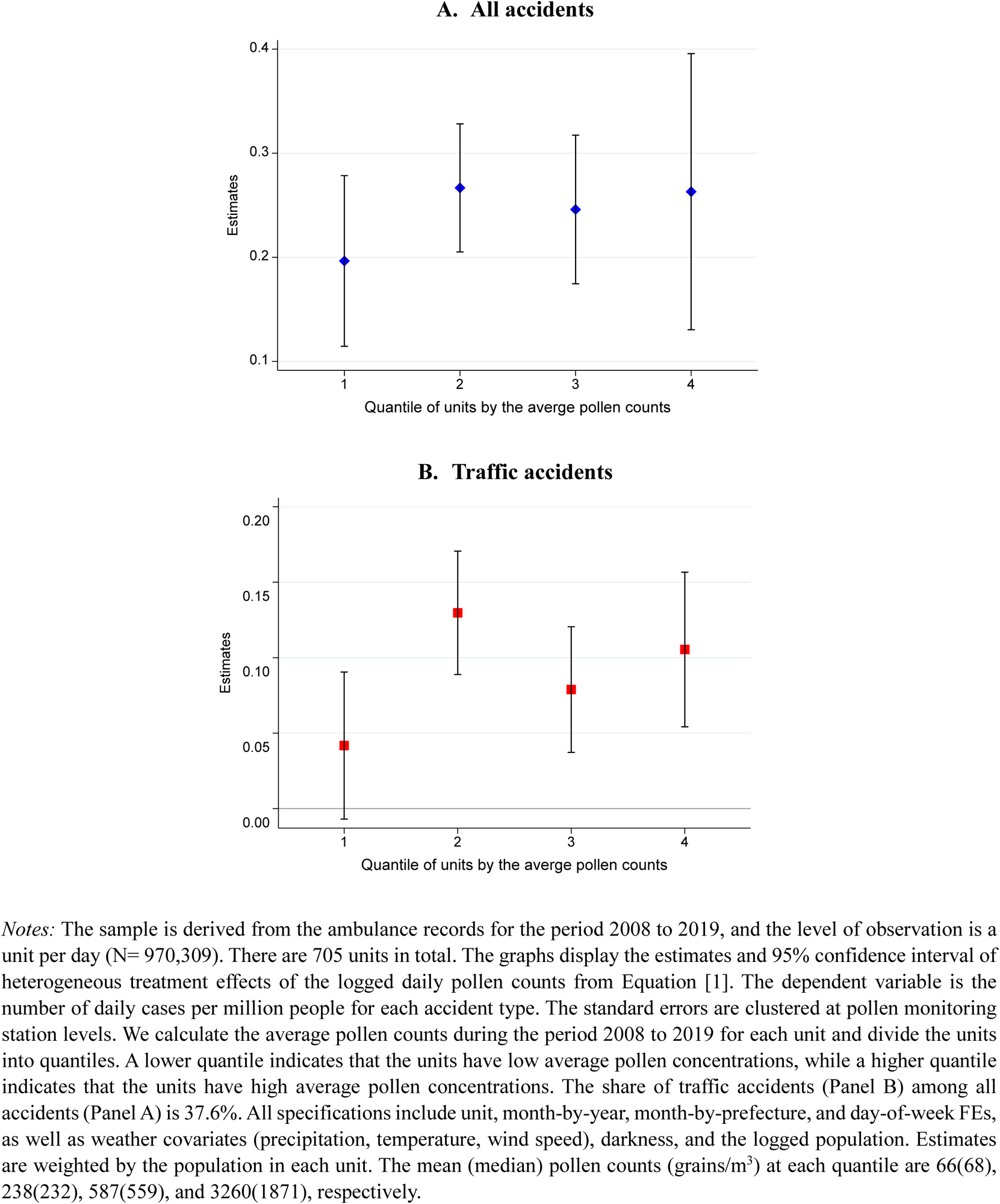
Treatment effects by regions with a high pollen concentration vs. regions with a low pollen concentration.

**Table C1.** Different time FE.

|  | (1) | (2) | (3) | (4) | (5) | (6) |
| --- | --- | --- | --- | --- | --- | --- |
| **A. All accidents** |  |  |  |  |  |  |
| ln(pollen counts) | 0.231***<br>(0.020) | 0.217***<br>(0.019) | 0.235***<br>(0.020) | 0.241***<br>(0.020) | 0.280***<br>(0.022) | 0.331***<br>(0.024) |
| R-squared | 0.46 | 0.46 | 0.46 | 0.47 | 0.46 | 0.49 |
| N | 970,309 | 970,309 | 970,309 | 970,309 | 970,309 | 969,623 |
| Mean of dep. Var | 33.03 | 33.03 | 33.03 | 33.03 | 33.03 | 33.03 |
| **B. Traffic accidents** |  |  |  |  |  |  |
| ln(pollen counts) | 0.079***<br>(0.012) | 0.079***<br>(0.012) | 0.091***<br>(0.013) | 0.084***<br>(0.013) | 0.118***<br>(0.014) | 0.140***<br>(0.015) |
| R-squared | 0.24 | 0.24 | 0.24 | 0.24 | 0.24 | 0.27 |
| N | 970,309 | 970,309 | 970,309 | 970,309 | 970,309 | 970,306 |
| Mean of dep. Var | 12.41 | 12.41 | 12.41 | 12.41 | 12.41 | 12.41 |
| Unit FE | X | X | X |  | X |  |
| Day-of-week FE | X | X | X | X | X | X |
| Month-year FE | X |  | X | X |  |  |
| Month-prefecture FE | X | X |  |  |  |  |
| Month & Year FE |  | X |  |  |  |  |
| Month-unit FE |  |  |  | X |  |  |
| Month-year-prefecture FE |  |  |  |  | X |  |
| Month-year-unit FE |  |  |  |  |  | X |

*Notes:* The sample is derived from the ambulance records for the period 2008 to 2019, and the level of observation is a unit per day (N= 970,309). There are 705 units in total. The dependent variable is the number of daily cases per million people for each accident type. The estimates from Equation [1] are reported along with the standard errors clustered at pollen monitoring station levels in parentheses. In addition to the FEs in the table, we include weather covariates (precipitation, temperature, wind speed), darkness, and the logged population. Column (1) replicates the results from Table 2 (baseline) for ease of comparison. The share of traffic accidents (Panel B) among all accidents (Panel A) is 37.6%. Estimates are weighted by the population in each unit. Significance levels: \*\*\* $p < 0.01$ , \*\* $p < 0.05$ , \* $p < 0.10$

**Table C2.** Different levels of clustering.

| Clustering variables | N of clusters | A. All accidents |  | B. Traffic accidents |  |
| --- | --- | --- | --- | --- | --- |
|  |  | 0.231 |  | 0.079 |  |
| Monitoring stations (baseline) | 120 | (0.020) | *** | (0.012) | *** |
| Monitoring stations and date | 120 + 1796 | (0.028) | *** | (0.016) | *** |
| Monitoring stations and month-year | 120 + 60 | (0.030) | *** | (0.019) | *** |
| Unit | 705 | (0.019) | *** | (0.012) | *** |
| Unit and date | 705 + 1796 | (0.028) | *** | (0.016) | *** |
| Unit and month-year | 705 + 60 | (0.031) | *** | (0.019) | *** |
| Prefecture | 47 | (0.016) | *** | (0.012) | *** |
| Prefecture and date | 47 + 1796 | (0.025) | *** | (0.015) | *** |
| Prefecture and month-year | 47 + 60 | (0.028) | *** | (0.018) | *** |
| Conley (50 km) | - | (0.018) | *** | (0.012) | *** |
| Conley (100 km) | - | (0.019) | *** | (0.012) | *** |
| Conley (150 km) | - | (0.020) | *** | (0.013) | *** |

*Notes:* The sample is derived from the ambulance records for the period 2008 to 2019, and the level of observation is a unit per day (N= 970,309). The dependent variable is the number of daily cases per million people for each accident type. The estimates from Equation [1] are reported. All specifications include unit, month-by-year, month-by-prefecture, and day-of-week FEs, as well as weather covariates (precipitation, temperature, wind speed), darkness, and the logged population. The number of pollen monitoring stations, units, and prefectures are 120, 705, and 46, respectively. The last three rows report the spatially clustered standard errors as in Conley (1999) with 50, 100, and 150 km distance cutoffs, respectively. The share of traffic accidents (Panel B) among all accidents (Panel A) is 37.6%. Estimates are weighted by the population in each unit. Significance levels: \*\*\* $p < 0.01$ , \*\* $p < 0.05$ , \* $p < 0.10$

**Table C3.** Alternative specifications.

|  | (1) | (2) | (3) |
| --- | --- | --- | --- |
|  | level-log<br>OLS<br>(Baseline) | log-log<br>OLS | Poisson<br>pseudo-maximum<br>likelihood<br>(PPML) |
| **A. All accidents** |  |  |  |
| ln(pollen counts) | 0.231***<br>(0.020) | 0.0054***<br>(0.0006) | 0.030***<br>(0.003) |
| R-squared | 0.46 | 0.90 | - |
| N | 970,309 | 970,309 | 970,309 |
| **B. Traffic accidents** |  |  |  |
| ln(pollen counts) | 0.079***<br>(0.012) | 0.0056***<br>(0.0010) | 0.013***<br>(0.002) |
| R-squared | 0.24 | 0.80 | - |
| N | 970,309 | 970,309 | 970,309 |
| Unit FE | X | X | X |
| Day-of-week FE | X | X | X |
| Month-year FE | X | X | X |
| Month-prefecture FE | X | X | X |

**Table C4.** Placebos.

|  | (1) | (2) | (3) |
| --- | --- | --- | --- |
|  | Baseline | Assigning last<br>year's pollen<br>counts | Assigning next<br>year's pollen<br>counts |
| **A. All accidents** |  |  |  |
| ln(pollen counts) | 0.231***<br>(0.020) | 0.008<br>(0.022) | 0.021<br>(0.022) |
| R-squared | 0.46 | 0.47 | 0.46 |
| N | 970,309 | 879,777 | 881,226 |
| Mean of dep. var | 33.03 | 33.21 | 32.89 |
| **B. Traffic accidents** |  |  |  |
| ln(pollen counts) | 0.079***<br>(0.012) | -0.018<br>(0.012) | -0.002<br>(0.011) |
| R-squared | 0.24 | 0.24 | 0.24 |
| N | 970,309 | 879,777 | 881,226 |
| Mean of dep. var | 12.41 | 12.27 | 12.59 |
| Unit FE | X | X | X |
| Day-of-week FE | X | X | X |
| Month-year FE | X | X | X |
| Month-prefecture FE | X | X | X |

## Appendix D: Police Records

Police records contain all 690,415 (or 106,533 in pollen season) traffic accidents for the period 2019 to 2020. The data are at the individual level of accidents and include information on the location, date, and time the accident occurred. Unlike the ambulance service, where the unit of service is a unit (N= 705), the police service is administered at the municipality level (N= 1,700). Thus, we aggregate the number of casualties to the municipality-day level by adding the hourly observations within the municipalities.

Figure D1 displays the binscatter plots of the relationship between the logged average daily pollen counts (grains/m^3^) and the number of deaths from traffic accidents per million people after controlling for municipality, month-by-year, month-by-prefecture, and day-of-the-week FEs. The figure shows the positive relationship between the two variables.

Table D1 reports the estimates from Equation [1], where unit FE is replaced by municipality FE. For ease of comparison, Column (1) replicates the death/fatal estimates for traffic accidents from the 2008 to 2019 ambulance records (from the first row of Panel B in Figure 4). Column (2) reports the mortality estimates from the 2019 to 2020 police records. The estimate of 0.0040 (*p*-value<0.01) in Column (2) is larger than the estimate of 0.0026 from Column (1), indicating that we may have slightly underestimated the effect of pollen exposure on the number of deaths as a result of traffic accidents. However, the two estimates are not statistically distinguishable at the conventional level.

**Figure D1.**
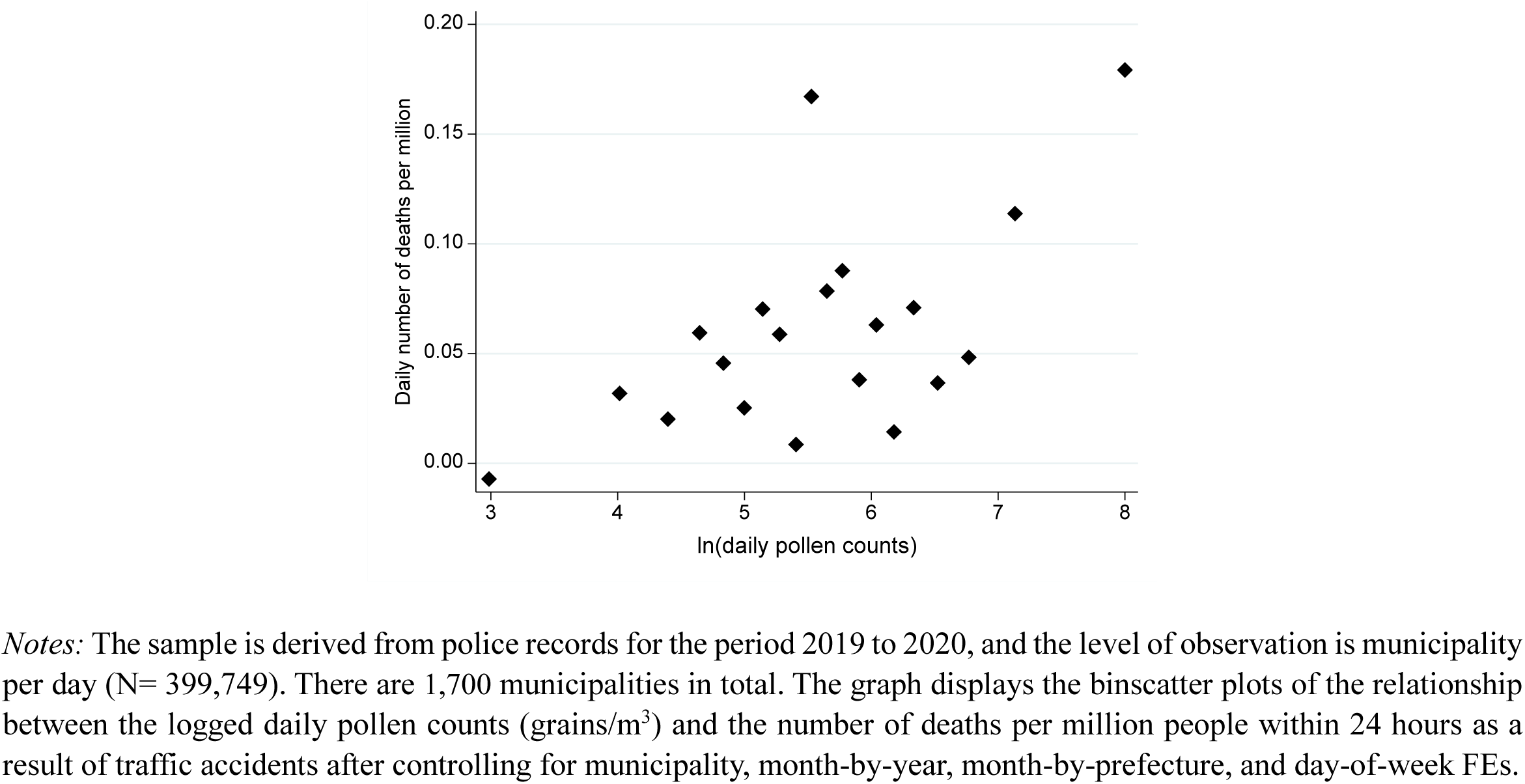
Pollen and mortality from traffic accidents (police records)

**Table D1.** Mortality as a result of traffic accidents.

(ambulance records vs. police records)
|  | Ambulance<br>Record | Police<br>Record |
| --- | --- | --- |
|  | (1) | (2) |
| ln(pollen counts) | 0.0026***<br>(0.0007) | 0.0040***<br>(0.0015) |
| R-squared | 0.01 | 0.01 |
| N | 970,309 | 399,749 |
| N of unit/municipality | 705 | 1,700 |
| N of clusters | 120 | 120 |
| Mean of dep. var | 0.16 | 0.12 |
| Unit/municipality FE | X | X |
| Day-of-week FE | X | X |
| Month-year FE | X | X |
| Month-prefecture FE | X | X |

*Notes:* The sample for Column (1) is derived from the ambulance records for the period 2008 to 2019, while the sample for Column (2) is derived from police records for the period 2019 to 2020. The level of observation is unit per day for Column (1) and municipality per day for Column (2). There are 705 units and 1,700 municipalities in total. The dependent variable is the number of deaths as a result of traffic accidents per million people. The estimates from Equation [1] are reported along with the standard errors clustered at pollen monitoring station levels in parentheses. In addition to the FEs in the table, we include weather covariates (precipitation, temperature, wind speed), darkness, and the logged population. Column (1) is identical to the first row of Panel B in Figure 4. Estimates are weighted by the population in each unit in Column (1) and by the population in each municipality in Column (2). Significance levels: \*\*\* $p < 0.01$ , \*\* $p < 0.05$ , \* $p < 0.10$

## Appendix E: Avoidance behaviors from in-home scanner data

**Figure E1.**
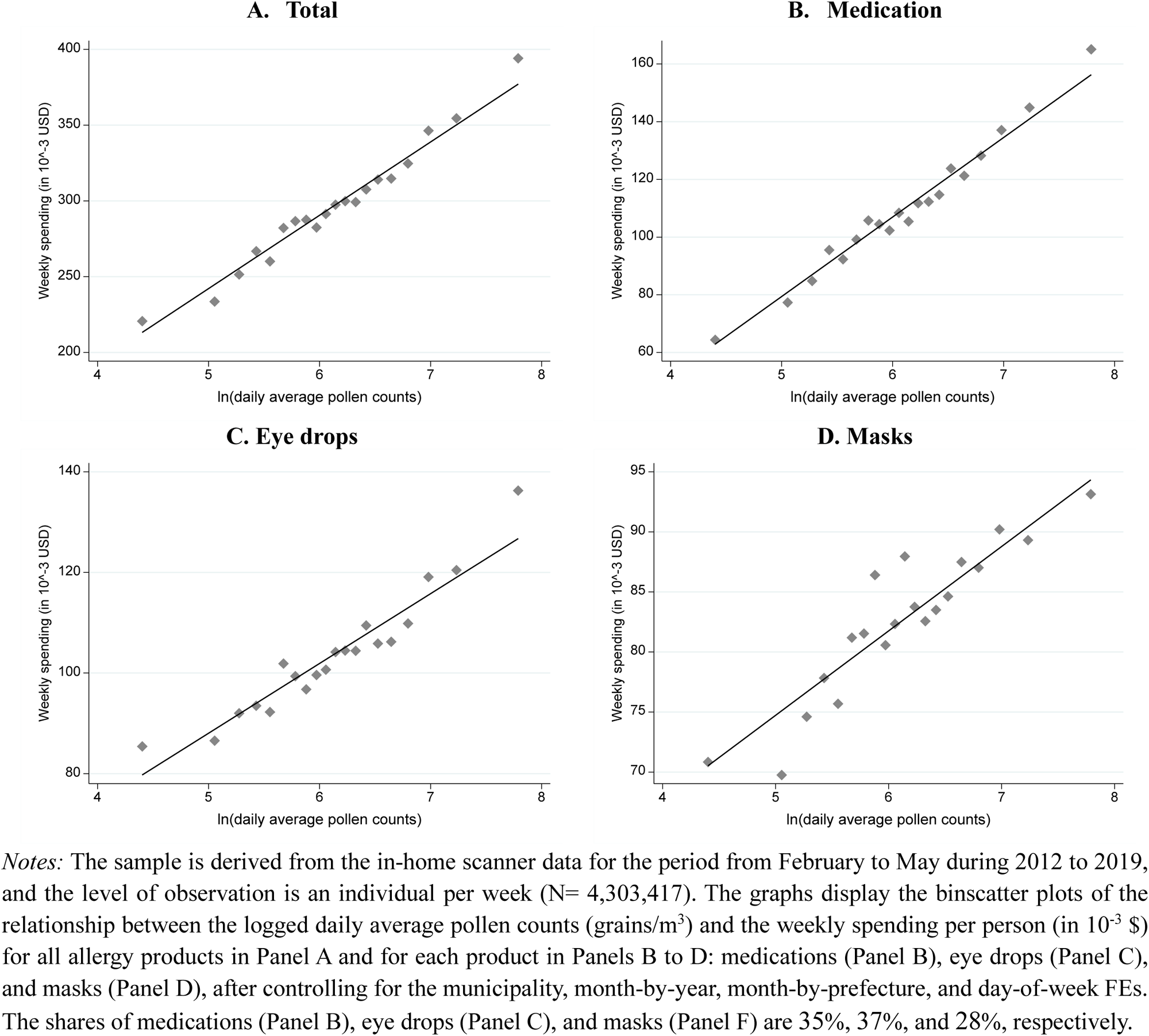
Pollen counts and weekly spending on allergy products.

**Table E1.** Summary statistics of in-home scanner data.

| **Variables** | **Obs** | **Mean** | **Std. dev.** | **Min** | **Max** |
| --- | --- | --- | --- | --- | --- |
| **<u>A. Outcomes (weekly per person in 10<sup>-3</sup> \$)</u>** | | | | | |
| Spending: Total | 4,303,417 | 295.78 | 2343.95 | 0 | 2,141,620 |
| Spending: Medications | 4,303,417 | 109.94 | 1528.34 | 0 | 227,200 |
| Spending: Eye drops | 4,303,417 | 103.41 | 1018.20 | 0 | 104,000 |
| Spending: Masks | 4,303,417 | 82.52 | 1324.56 | 0 | 2,141,620 |
| **<u>B. Individual characteristics</u>** |  |  |  |  |  |
| Ages: 0–24 | 4,303,417 | 0.13 | 0.33 | 0 | 1 |
| Ages: 25–44 | 4,303,417 | 0.38 | 0.48 | 0 | 1 |
| Ages: 45–64 | 4,303,417 | 0.38 | 0.49 | 0 | 1 |
| Ages: 65 years and older | 4,303,417 | 0.11 | 0.32 | 0 | 1 |
| Female | 4,303,417 | 0.50 | 0.50 | 0 | 1 |
| Student | 4,303,417 | 0.11 | 0.31 | 0 | 1 |
| Household head | 4,303,417 | 0.46 | 0.50 | 0 | 1 |
| Have kids | 4,303,417 | 0.55 | 0.50 | 0 | 1 |
| Own house | 4,303,417 | 0.58 | 0.49 | 0 | 1 |
| Annual salary category | 4,303,417 | 2.61 | 2.22 | 1 | 13 |
| **<u>C. Regressors (daily average in a week)</u>** |  |  |  |  |  |
| Pollen counts (grains/m<sup>3</sup>) | 4,303,417 | 843.2 | 1337.4 | 0 | 22,812 |
| Logged (Pollen counts) | 4,303,417 | 0.1 | 0.2 | 0 | 3.9 |
| Precipitation (mm) | 4,303,417 | 12.0 | 6.0 | -8 | 26 |
| Average temperature (°C) | 4,303,417 | 2.8 | 0.9 | 0 | 11.6 |
| Average wind speed (m/s) | 4,303,417 | 10.5 | 1.3 | 7 | 13 |

*Notes:* The sample is derived from the in-home scanner data for the period from February to May during 2012 to 2019, and the level of observation is an individual per week. The spending is the weekly aggregate per person (in 10<sup>-3</sup> \$).

## Appendix F: Avoidance Behaviors from Google Trends/tweets

**Figure F1.**
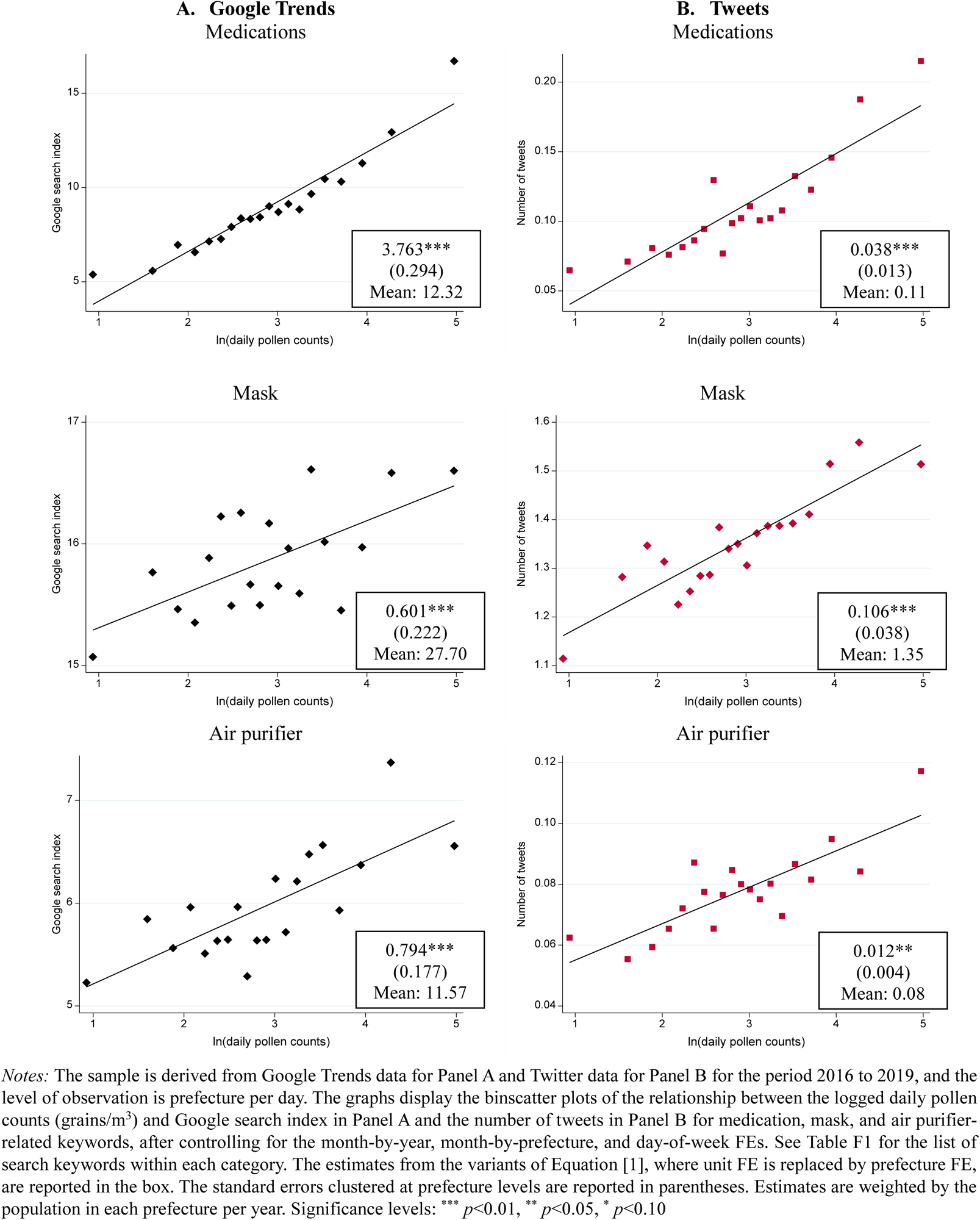
Pollen and Google Trends/tweets.

**Table F1.** List of search keywords.

| **Category #** | **Categories** | **English translation of keywords** |
| --- | --- | --- |
| 1 | **Medications**<br>(product name of<br>popular allergy<br>medications) | Alesion<br>Allegra<br>Claritin |
| 2 | **Mask** | Mask |
| 3 | **Air purifier** | Air purifier |

*Notes:* This table lists keywords in each category translated from Japanese to English. The keywords in Japanese are available upon request.

## Appendix G: Avoidance Behaviors from Geolocation Data

Our geolocation data are called “Mobile Spatial Statistics” (MSS), provided by NTT DOCOMO, Inc., Japan’s largest mobile phone carrier. Based on the location information of 85 million users of NTT DOCOMO (as of March 2022), MSS provides the population estimate at a 500×500-meter mesh at an hourly level across Japan. See Terada et al. (2013) for detailed procedures for constructing population estimates.

Our dataset on mobility measures is constructed as follows. First, for each municipality, we choose a 500×500-meter mesh with the largest number of establishments in the customer service industry (e.g., accommodation, restaurants, and entertainment) based on information obtained from the 2016 Economic Census (MIC 2019).^1^ We choose the service industry to capture bustling areas (e.g., business districts and shopping and dining areas), which are more likely to represent the population engaging in outdoor activities. Second, we provide the list of the meshes to NTT DOCOMO, Inc, which returns the estimated population at each mesh for the period February 2014 to May 2019. Third, we collapse the estimated population at the unit level by taking the average of all municipalities in the unit. We use the estimated population at 2 pm, as the daily population in commercial areas tends to peak around that time (Seike et al. 2015).^2^ We treat this measure as a proxy for engaging in outdoor activities (“outdoor population,” hereafter) to examine avoidance behaviors.

Finally, we briefly discuss the advantages and disadvantages of this dataset. There are two types of geolocation data in Japan: the first come from the leading smartphone mapping application in Japan (“Docomo Chizu NAVI”), which collects GPS coordinates of each smartphone device whenever the device is turned on. The most attractive feature of this application is that it effectively follows each individual over time (but only in very recent years); researchers can identify “home” locations as the most frequent locations of geographically contiguous stays (Miyauchi et al. 2021), and measure whether the individuals leave their home. The drawback is that the sample is limited to individuals who gave permission to share location information, causing selection issues to the users of the particular application as well as those who gave permission, and also resulting in a small sample size (545,000 users as of 2019). Meanwhile, the second type of data, like ours, based on information transmission from each mobile terminal (of 85 million users in our case) to base stations as long as mobile devices are turned on, are more nationally representative with wide spatial coverage of the entire country, while they only provide the hourly estimated population in a given area.

As the main objective of this study is to provide nationally representative estimates of the effects of pollen exposure on accidents and corresponding avoidance behaviors over an extended period of time, we chose the latter dataset for analysis in this study. Owing to its representativeness and long time span of the sample, this dataset has been widely used, especially in measuring people’s mobility

**Figure G1.**
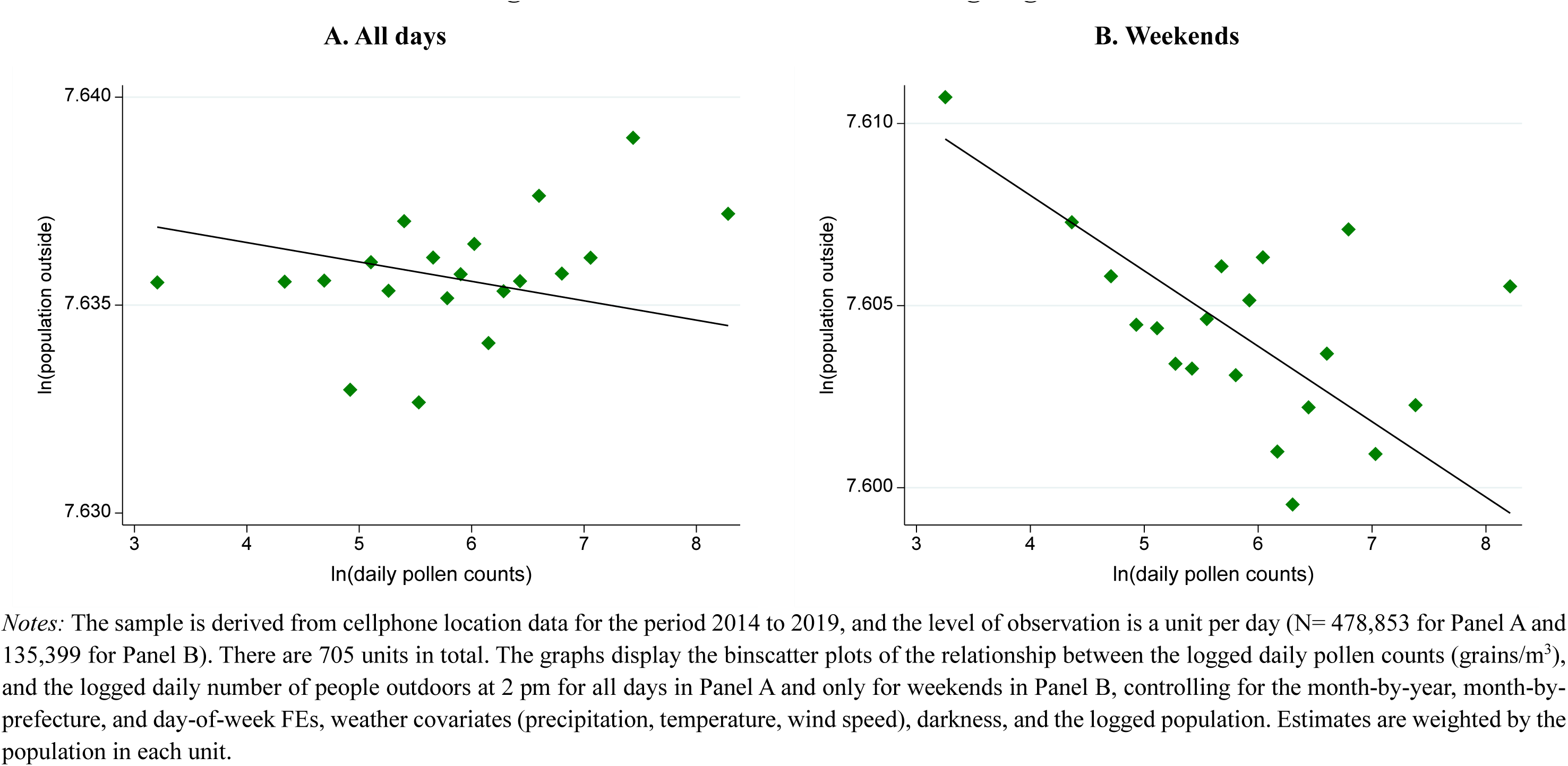
Pollen and avoidance of going out.

**Table G1.** Avoidance of going out and accidents.

|  | **A. All accidents** |  |  | **B. Traffic accidents** |  |  |
| --- | --- | --- | --- | --- | --- | --- |
|  | All | Weekdays | Weekends | All | Weekdays | Weekends |
|  | (1) | (2) | (3) | (4) | (5) | (6) |
| ln(outdoor population) | 0.960**<br>(0.418) | 0.465<br>(0.441) | 2.007**<br>(0.775) | 0.484**<br>(0.224) | 0.553**<br>(0.254) | 1.024***<br>(0.341) |
| R-squared | 0.49 | 0.48 | 0.51 | 0.24 | 0.24 | 0.23 |
| N | 478,853 | 343,454 | 135,399 | 478,853 | 343,454 | 135,399 |
| Unit FE | X | X | X | X | X | X |
| Day-of-week FE | X | X | X | X | X | X |
| Month-year FE | X | X | X | X | X | X |
| Month-prefecture FE | X | X | X | X | X | X |

**Table G2.** Avoidance of going out with full covariates.

Outcome: logged outdoor population
|  | A. All | B. By type of day |  |
| --- | --- | --- | --- |
|  |  | Weekdays | Weekends |
|  | (1) | (2) | (3) |
| ln(pollen counts) | -0.0005<br>(0.0006) | 0.0000<br>(0.0006) | -0.0021***<br>(0.0007) |
| Rainfall (base: no rainfall) |  |  |  |
| <1 mm | -0.0061***<br>(0.0013) | -0.0059***<br>(0.0011) | -0.0047**<br>(0.0019) |
| 1 mm ≤ & <2 mm | -0.0167***<br>(0.0035) | -0.0144***<br>(0.0024) | -0.0208***<br>(0.0080) |
| ≥2 mm | -0.0167***<br>(0.0045) | -0.0150***<br>(0.0041) | -0.0249***<br>(0.0082) |
| Mean temperature (base: <0 °C) |  |  |  |
| [0, 5) °C | 0.0018<br>(0.0033) | 0.0068**<br>(0.0034) | 0.0096<br>(0.0064) |
| [5, 10) °C | 0.0111***<br>(0.0036) | 0.0139***<br>(0.0037) | 0.0189***<br>(0.0067) |
| [10, 15) °C | 0.0089**<br>(0.0043) | 0.0120***<br>(0.0043) | 0.0160**<br>(0.0070) |
| [15, 20) °C | 0.0061<br>(0.0045) | 0.0094*<br>(0.0049) | 0.0172**<br>(0.0068) |
| [20, 25) °C | 0.0120***<br>(0.0043) | 0.0165***<br>(0.0046) | 0.0200***<br>(0.0069) |
| ≥ 25 °C | 0.0273***<br>(0.0057) | 0.0264***<br>(0.0066) | 0.0319***<br>(0.0079) |
| Mean wind speed | -0.0022***<br>(0.0005) | -0.0021***<br>(0.0005) | -0.0023***<br>(0.0007) |
| Darkness | -0.0084***<br>(0.0014) | -0.0090***<br>(0.0012) | -0.0073***<br>(0.0023) |
| ln(population) | 1.3108***<br>(0.1975) | 1.2630***<br>(0.1997) | 1.4497***<br>(0.2074) |
| R-squared | 0.98 | 0.99 | 0.99 |
| N | 478,853 | 343,454 | 135,399 |
| Pref FE | X | X | X |
| Day-of-week FE | X | X | X |
| Month-year FE | X | X | X |
| Month-prefecture FE | X | X | X |

*Notes:* The sample is derived from cellphone location data for the period from February to May for the years 2014 to 2019, and the level of observation is a unit per day. There are 705 units in total. The estimates from variants of Equation [1] are reported along with the standard errors clustered at pollen monitoring station levels in parentheses. The outcome is the logged daily outdoor population at 2 pm. Estimates are weighted by the population in each unit. Significance levels:
\*\*\* $p < 0.01$ , \*\* $p < 0.05$ , \* $p < 0.10$

## Appendix H: Data Appendix

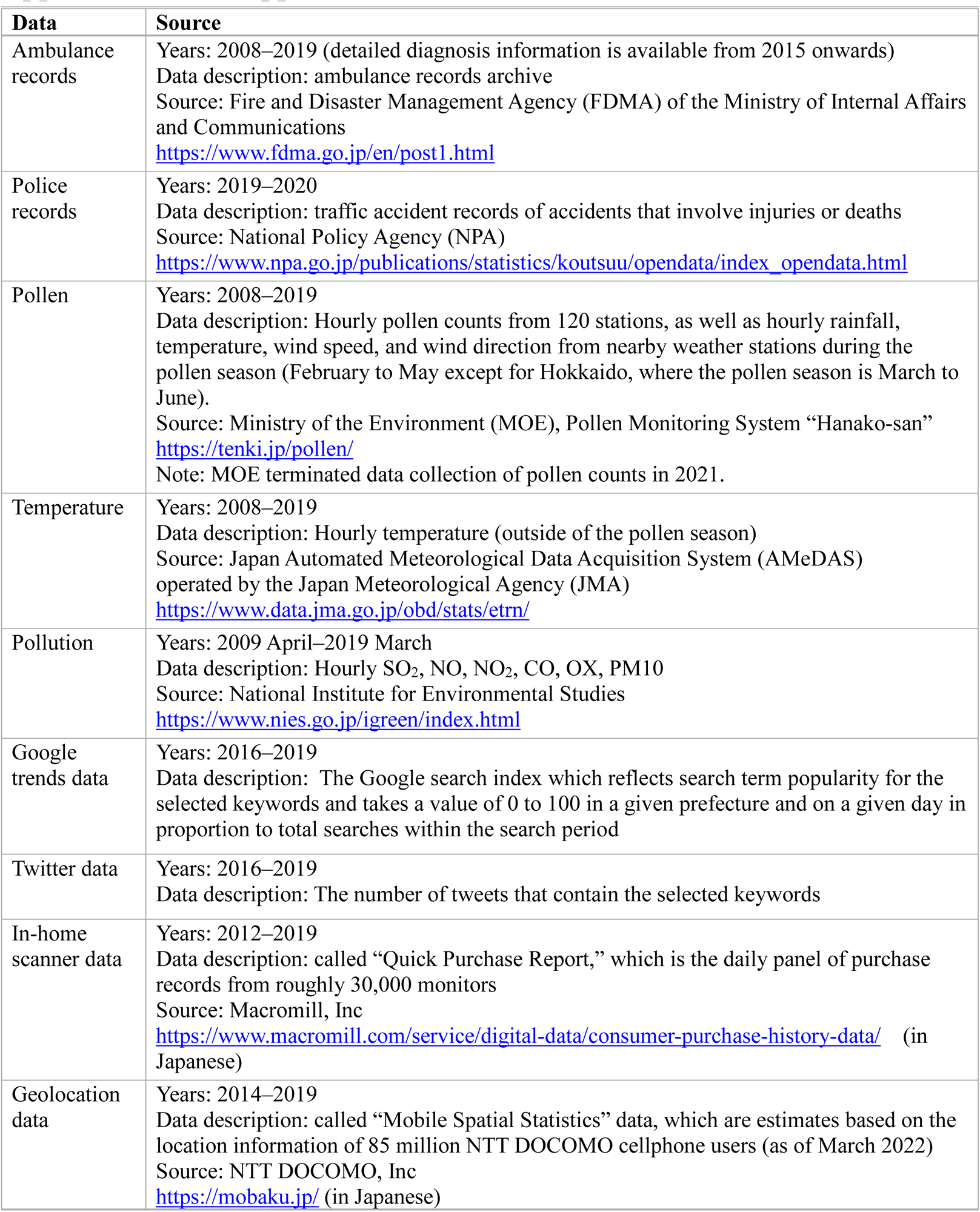

## Footnotes

1 Economists gather knowledge on the impact of exposure to air pollution on a variety of outcomes ranging from health, human capital, labor productivity, and crime. See Graff Zivin and Neidell (2013) and Aguilar-Gomez et al. (2022) for summaries of relevant literature.

2 Traffic accidents are the second leading cause of accidental deaths (after asphyxia), with a recorded average of over 4,000 and 700,000 annual fatalities and injuries, respectively, for the period 2008 to 2019 (MHLW 2009; NPA 2022), in a total population of 127 million in Japan.

3 Broten et al. (2019) find that workers who are injured on the job face a subsequent earnings penalty of 8% on average, and this increases to 30% for those who are permanently disabled.

4 The current evidence presents the significant adaptation potential in the context of mortality to heat exposure (Barreca et al. 2016; Heutel et al. 2021), infant mortality to dust (Adhvaryu et al. 2022), with mixed evidence for workplace injuries to heat (Dillender 2021; Park et al. 2021).

5 The prevalence rates of SAR in 2019 are 30.1% (ages 5–9), 49.5% (10–19), 47.5% (20–29), 46.8% (30–39), 47.5% (40–49), 45.7% (50–59), 36.9% (60–69), and 20.5% (70–) (Matsubara et al. 2020). In addition, the prevalence rate is generally high in urban areas, and in Tokyo, it increased from 19.4% in 1996 to approximately 48.8% in 2016 (Tokyo Metropolitan Institute of Public Health 2017).

6 Vuurman et al. (2014) argue that the magnitude of the impairing effects of allergic rhinitis on driving is comparable to the effects of blood alcohol content (BAC) of 0.05%, which is the legal limit in many countries. Smith (2016) also demonstrates that a one-hour sleep loss increases the probability of being in a drowsiness-related fatal motor vehicle accident by 46%.

7 See for example: pollution (e.g., Chang et al. 2016, 2019; Ebenstein et al. 2016; Graff Zivin and Neidell 2012; He et al. 2019; La Nauze and Severnini 2021; Sager 2019; Zhang et al. 2018) and temperature (e.g., Adhvaryu et al. 2020; Lavy et al. 2022; Park et al. 2021; Somanathan et al. 2021).

8 The number of pollen stations has remained at 120 since 2008; thus, our estimates are not affected by the increase or decrease in the number of monitoring stations. The movement of monitoring stations is limited to a handful of stations, and the distance of movement is very small.

9 Okinawa prefecture, which is the southernmost remote island in Japan, with a different climate from the rest of Japan, has no pollen station, as little pollen is observed. We exclude Okinawa from the entire analysis.

10 Chalfin et al. (2019), who examine the effect of pollen on crime in cities in the United States, use criminal records from stations within 30 miles (48 km) of the city center, while the National Allergy Bureau suggests that pollen measurements are valid within a 20-mile (32 km) radius of each station.

11 The exception is Hokkaido prefecture, which is located in the far north (with four stations), and thus the monitoring period is delayed by one month (March to June).

12 Tokyo is excluded from the sample because (i) metropolitan Tokyo (23 wards of central Tokyo and most cities in Tokyo) falls under one ambulance operating system (Tokyo Fire Department), which provides no regional variation in pollen counts, and (ii) data from metropolitan Tokyo are publicly available only from 2016 onwards.

13 The ambulance records also include emergency cases due to illness (72.3% of all records). Since our focus is accidents, we extract data on five types of accidents from the ambulance archives: traffic accidents, work-related injuries, sports injuries, fire accidents, and other accidents, which add up to 25.9% of all records. The remaining records relate to self-injurious activities, assault, drowning, natural disasters, and other categories (1.8%).

14 The timestamp of each accident reflects the time at which the accident is reported to emergency response units rather than the actual time that the accident occurred. This might result in some degree of measurement error in terms of the hour (more likely) than the date. As a result, we aggregate accidents to the daily level, following the existing literature (e.g., Park et al. 2021).

15 For example, these accidents range from minor to major ones and include: (i) slipping on a step on the road and falling down, (ii) slipping on a snowy road and falling down, (iii) spilling a pot and getting burned, and (iv) slamming a finger in a screen door.

16 We have truncated pollen counts at the 99.9 percentile (55,104 grains/m^3^) to account for outliers.

17 We add one to account for zero pollen counts (0.83%) before taking the log. Later, we show that our results are robust to dropping these observations and taking logs without adding 1 in Table 3. Bensnes (2016) and Marcotte (2017) also take the log form of pollen counts.

18 The cutoff of 2016 is motivated by data completeness as well as the fact that Google made a change to its data collection system on January 1, 2016. We follow Brodeur et al. (2021) to construct daily-level Google Trends data across multiple years using overlapping periods of daily and weekly data.

19 We assign prefecture based on the location at the time of the tweet.

20 For example, the number of work-related injuries leading to death that are recorded in our ambulance records for 2019 is 392, while the corresponding number reported to the Ministry of Health, Labour and Welfare (MHLW) is 845 (MHLW 2020). Similarly, the number of work-related injuries, including injuries of *all* severity levels in our ambulance records in 2019, is 50,578, while the total number of work-related injuries that result in either death or at least four days of work absenteeism reported to the MHLW is 125,611 (MHLW 2020).

21 The correlation between pollen counts and other pollutants is as low as 0.02 (CO)-0.12 (PM10), allowing pollen to have an independent impact on the number of accidents that occur.

22 The short-lived effect of pollen exposure is consistent with previous literature documenting the short-lived impact of pollution exposure on health (e.g., Schlenker and Walker 2016), and productivity (e.g., Graff Zivin and Neidell 2012; Chang et al. 2016, 2019).

23 The ambulance records include detailed diagnostic information (equivalent to ICD10) from 2015 onward.

24 One potential reason for this observation is that the police records include all the deaths *within 24 hours* due to traffic accidents, unlike ambulance records, which only include deaths that occur at the time of hospital admission.

25 For completeness, the heterogeneity analysis for the traffic accidents is displayed in Figure C3.

26 More precisely, the sickness level is the function of the *dose* of pollen one is exposed to, *Sick = g*(*Dose*), and the dose is determined by ambient pollen concentration (*Pollen*) and avoidance behaviors (*Avoid*), that is, *Dose* = *h*(*Pollen,Avoid*). Substituting it in the first equation yields *Stick* = *g(Pollen,Avoid)*, as shown in the main text.

27 For example, Neidell (2009) finds that the “reduced-form” effect of ozone is 40% and 160% smaller for the elderly and children, respectively, than the pure “biological” effect.

28 Checking against the conceptual framework, each type of avoidance behavior is considered to correspond to different pathways to reduce the risk of accidents. The purchase of allergy products corresponds to the *Avoid* term in *Sick* = *g*(*Pollen, Avoid*), which lowers the sickness level and, hence, *indirectly* reduces the accidental risk. By contrast, curtailing outdoor activities mainly corresponds to the *Avoid* term in *Accidents* = *f*(*Sick, Avoid*), which *directly* reduces the likelihood of outdoor accidents. Of course, this also contains the first type of avoidance effects, as staying indoors limits pollen exposure, thus lowering the sickness level.

29 The QPR monitors are selected in each region to represent the gender, age group, and family structure (marital status and existence of a housemate) of the country. To maintain data quality, the monitor is replaced by another one with similar characteristics when unusual scans are detected, or scans are not observed for several weeks. Thus, the number of active QPR monitors at any given time is maintained at approximately 30,000, and the total number of unique QPR monitors during our sample period is 70,795.

30 While medications are intended to mitigate the symptoms (e.g., stop a runny nose), they may not always reduce the risk of accidents because they induce drowsiness for some individuals (i.e., the sign of 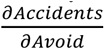 is unclear for them).

31 Indeed, the weekly analysis provides larger estimates than would be implied by a linear scale-up of the daily estimate (not shown).

32 These results are consistent with those of previous studies, which document a similar relationship between pollen counts and sales of OCT allergy medication in New York City in the United States (Ito et al. 2015), and Japan (Kuroda 2022).

33 Owing to its representativeness and the long time span of the sample, this dataset has been widely used, especially for measuring people’s mobility during the COVID-19 pandemic (e.g., Kondo 2021; Kuroda et al. 2022).

34 As in the other studies on physical movement, we cannot distinguish two possibilities for staying indoors: people may be extremely sick and have to stay home or they may display some form of avoidance.

35 We do not similarly verify whether the purchase of allergy products effectively reduces accidents because the regional sampling units in in-home scanner data are only ten divisions (albeit nationally representative), and they do not correspond to the detailed regional units of the ambulance records (N= 705).

36 See Table G2 for the estimates of other weather covariates with expected signs: While the outdoor population increases as the temperature rises, it decreases as the average wind speed and duration of darkness increases.

37 This distinction is important for policy considerations since if there is a problem with access to information, we would suggest that the government should attempt to increase the dissemination of pollen information more widely (Barwick et al. 2020; Jha and Nauze 2022). However, if access to information is of a sufficient level, as in this case, but attention is lacking, we would have to propose more effective ways to raise awareness of the risks and encourage further behavioral change.

38 We classify both nasal and oral medications for seasonal allergies into two types based on any particular mention of whether driving is not allowed after taking the medication, including the ones listed in Table A1.

39 See Figure A5, which plots the spatial variation in pollen counts across the country.

40 We acknowledge that we make two strong assumptions here: (i) the level of protective technologies stay the same; (ii) the marginal treatment effect of an unanticipated “weather” shock documented so far is identical to the marginal effect of an anticipated “climate” shift. Past studies have adopted a similar approach when projecting the impact of these gradual changes on income (Deryugina and Hsiang 2014), mortality (Deschênes and Greenstone 2011), amenity values (Baylis 2020), and other outcomes.

41 We determine the prospective additional cost to be $3.2 million for fatal accidents (those resulting in death), $365,000 for accidents resulting in serious injuries, and $38,000 for those resulting in both moderate and light injuries (Bünnings and Schiele 2021). For simplicity, an exchange rate of 1.5 $/£ is used. Unfortunately, to our knowledge, there are no appropriate estimates of accident costs at each severity level in Japan.

42 Panel B of Table 5, using the number of days where the temperature increased above 30℃ in the previous year (Panel B of Figure 1), gives roughly 40% larger estimates of social cost than those in Panel A of Table 6, which uses the maximum temperature.

43 For example, the central estimate for the value of statistical life used by the Environmental Protection Agency in the United States is $9.8 million in 2021, and Smith (2016) similarly uses $4 million to $10 million per fatality, all of which are larger than the figure of $3.2 million that we utilize.

1 Owing to budgetary reasons, our data consist of one mesh per municipality. However, we confirm that our mobility measure captures overall daytime outdoor activities adequately, using the 2019 data for which we have an outdoor population for all meshes. Indeed, our mobility measure from the representative mesh correlates as high as 0.889 with the summary measure that uses the average of all the meshes with at least one establishment in the service industry.

2 We also examine alternative methods of constructing outdoor mobility measures, specifically, the difference between daytime (2pm) and nighttime (4am) populations and the ratio of daytime to nighttime populations. The results are qualitatively similar (not shown) mainly due to the fact that the nighttime population is relatively stable over time, and thus it does not provide much additional information after controlling unit FE. during the COVID-19 pandemic (e.g., Kondo 2021; Kuroda et al. 2022).

## References

Abadie, A., S. Athey, G. W. Imbens, J. Wooldridge. 2017. “When Should You Adjust Standard Errors for Clustering?” NBER Working Paper No. 24003.

Abouk, R., S. Adams. 2013. “Texting Bans and Fatal Accidents on Roadways: Do They Work? Or Do Drivers Just React to Announcements of Bans?” American Economic Journal: Applied Economics, 5(2): 179–199.

Adhvaryu, A., N. Kala, A. Nyshadham. 2020. “The Light and the Heat: Productivity Co-Benefits of Energy-Saving Technology.” The Review of Economics and Statistics, 102(4): 779–792.

Adhvaryu, A., P. Bharadwaj, J. Fenske, A. Nyshadham, R. Stanley. 2022. “Dust and Death: Evidence from the West African Harmattan.” The Economic Journal, forthcoming.

Aguilar-Gomez, S., H. Dwyer, J.S. Graff Zivin, M.J. Neidell. 2022. “This Is Air: The ‘Nonhealth’ Effects of Air Pollution.” Annual Review of Resource Economics, 14: 403–425.

Anderegg, W.R.L., J.T. Abatzoglou, L.D.L. Anderegg, L. Bielory, P.L. Kinney, L. Ziska. 2021. “Anthropogenic Climate Change is Worsening North American Pollen Seasons.” Proceedings of the National Academy of Sciences of the United States of America, 118(7): e2013284118.

Anderson, M., M. Hyun, J. Lee. 2022. “Bounds, Benefits, and Bad Air: Welfare Impacts of Pollution Alerts.” NBER Working Paper No. 29637.

Anstey, K., J. Wood, S. Lord, J. Walker. 2005. “Cognitive, Sensory and Physical Factors Enabling Driving Safety in Older Adults.” Clinical Psychology Review, 25(1): 45–65.

Anstey, K., M. Horswill, J. Wood, C. Hatherly. 2012. “The Role of Cognitive and Visual Abilities as Predictors in the Multifactorial Model of Driving Safety.” Accident Analysis & Prevention, 45: 766–774.

Barreca, A., K. Clay, O. Deschênes, M. Greenstone, J.S. Shapiro. 2016. “Adapting to Climate Change: The Remarkable Decline in the US Temperature-Mortality Relationship over the Twentieth Century.” Journal of Political Economy, 124(1): 105–159.

Barwick, P.J., S. Li, L. Lin, E. Zou. 2020. “From Fog to Smog: The Value of Pollution Information.” NBER Working Paper No. 26541.

Baylis, P. 2020. “Temperature and Temperament: Evidence from Twitter.” Journal of Public Economics, 184: 104161.

Bensnes, S.S. 2016. “You Sneeze, You Lose: The Impact of Pollen Exposure on Cognitive Performance during High-stakes High School Exams.” Journal of Health Economics, 49: 1– 13.

Brodeur, A., A.E. Clark, S. Fleche, N. Powdthavee. 2021. “COVID-19, Lockdowns and Well-being: Evidence from Google Trends.” Journal of Public Economics, 193: 104346.

Broten, N., M. Dworsky, D. Powell. 2019. “How do Alternative Work Arrangements Affect Income Risk after Workplace Injury?” NBER Working Paper No. 25989.

Bünnings, C., V. Schiele. 2021. “Spring Forward, Don’t Fall Back: The Effect of Daylight Saving Time on Road Safety.” The Review of Economics and Statistics, 103(1): 165–176.

Burke, M., S. Heft-Neal, J. Li, A. Driscoll, P. Baylis, M. Stigler, et al. 2022. “Exposures and Behavioural Responses to Wildfire Smoke.” Nature Human Behaviour, 6: 1351–1361.

Carleton, T., S. Hsiang. 2016. “Social and Economic Impacts of Climate.” Science, 353(6304): aad9837.

Chalfin, A., S. Danagoulian, M. Deza. 2019. “More Sneezing, Less Crime? Health Shocks and the Market for Offenses.” Journal of Health Economics, 68: 102230.

Chang, T.Y., J. Graff Zivin, T. Gross, M. Neidell. 2019. “The Effect of Pollution on Worker Productivity: Evidence from Call Center Workers in China.” American Economic Journal: Applied Economics, 11(1): 151–172.

Chang, T.Y., J. Graff Zivin, T. Gross, M. Neidell. 2016. “Particulate Pollution and the Productivity of Pear Packers.” American Economic Journal: Economic Policy, 8(3): 141– 169.

Cohen, F., A. Dechezleprêtre. 2022. “Mortality, Temperature, and Public Health Provision: Evidence from Mexico.” American Economic Journal: Economic Policy, 14(2): 161–192.

Conley, T.G. 1999. “GMM Estimation with Cross Sectional Dependence.” Journal of Econometrics, 92(1): 1–45.

Craig, T.J., J.L. McCann, F. Gurevich, M.J. Davies. 2004. “The Correlation between Allergic Rhinitis and Sleep Disturbance.” Journal of Allergy and Clinical Immunology, 114(5): S139– S145.

Cutter, W.B., Neidell, M. 2009. “Voluntary Information Programs and Environmental Regulation: Evidence from ‘Spare the Air’.” Journal of Environmental Economics and Management, 58(3): 253–265.

D’Amato, G., L. Cecchi, S. Bonini, C. Nunes, I. Annesi-Maesano, H. Behrendt, G. Liccardi, T. Popov, P. Van Cauwenberge. 2007. “Allergenic Pollen and Pollen Allergy in Europe.” Allergy, 62(9): 976–990.

Dell, M., B.F. Jones, B.A. Olken. 2014. “What Do We Learn from the Weather? The New Climate-Economy Literature.” Journal of Economic Literature, 52(3): 740–798.

Deryugina, T., S. Hsiang. 2014. “Does the Environment Still Matter? Daily Temperature and Income in the United States.” NBER Working Paper No. 20750.

Deryugina, T., G. Heutel, N.H. Miller, D. Molitor, J. Reif. 2019. “The Mortality and Medical Costs of Air Pollution: Evidence from Changes in Wind Direction.” American Economic Review, 109(12): 4178–4219.

Deschênes, O., M. Greenstone. 2011. “Climate Change, Mortality, and Adaptation: Evidence from Annual Fluctuations in Weather in the US.” American Economic Journal: Applied Economics, 3(4): 152–185.

Deschênes, O., M. Greenstone, J.S. Shapiro. 2017. “Defensive Investments and the Demand for Air Quality: Evidence from the NOx Budget Program.” American Economic Review, 107(10): 2958–2989.

Dillender, M. 2021. “Climate Change and Occupational Health: Are There Limits to Our Ability to Adapt?” Journal of Human Resources, 56(1): 184–224.

Ebenstein, A., V. Lavy, S. Roth. 2016. “The Long-Run Economic Consequences of High-Stakes Examinations: Evidence from Transitory Variation in Pollution.” American Economic Journal: Applied Economics, 8(4): 36–65.

Erbas, B., J.H. Chang, S. Dharmage, E.K. Ong, R. Hyndman, E. Newbigin, M. Abramson. 2007. “Do Levels of Airborne Grass Pollen Influence Asthma Hospital Admissions?” Clinical & Experimental Allergy, 37: 1641–1647.

European Commission. 2009. “Causes and Circumstances of Accidents at Work in the EU.” Luxembourg: Office for Official Publications of the European Communities. https://ec.europa.eu/eurostat/documents/53621/53703/Full-Publication%5BEN%5D-WO.pdf/6e90be02-c41e-43d6-87d4-68a4a7899ad1 (accessed May 23, 2022).

Forest Agency. 2021. “Kafun hassei-gen taisaku suishin jigyō” [Projects on Countermeasures Against Pollen Sources]. https://www.rinya.maff.go.jp/j/sin_riyou/kafun/attach/pdf/hojyo-7.pdf (accessed May 23, 2022).

Goel, S., J.M. Hofman, S. Lahaie, D.M. Pennock, D.J. Watts. 2010. “Predicting Consumer Behavior with Web search.” Proceedings of the National Academy of Sciences of the United States of America, 107(41): 17486–17490.

Graff Zivin, J., M. Neidell. 2012. “The Impact of Pollution on Worker Productivity.” American Economic Review, 102(7): 3652–3673.

Graff Zivin, J., M. Neidell. 2013. “Environment, Health, and Human Capital.” Journal of Economic Literature, 51(3): 689–730.

Graff Zivin, J., M. Neidell. 2014. “Temperature and the Allocation of Time: Implications for Climate Change.” Journal of Labor Economics, 32(1): 1–26.

Greiner, A.N., P.W. Hellings, G. Rotiroti, G.K. Scadding. 2011. “Allergic Rhinitis.” Lancet, 378: 2112–2122.

Hamaoui-Laguel, L., R. Vautard, L. Liu, F. Solmon, N. Viovy, D. Khvorostyanov, et al. 2015. “Effects of Climate Change and Seed Dispersal on Airborne Ragweed Pollen Loads in Europe.” Nature Climate Change, 5(8): 766–771.

He, J., H. Liu, A. Salvo. 2019. “Severe Air Pollution and Labor Productivity: Evidence from Industrial Towns in China.” American Economic Journal: Applied Economics, 11(1): 173– 201.

Hellgren, J., A. Cervin, S. Nordling, A. Bergman, L. Cardell. 2010. “Allergic Rhinitis and the Common Cold – High Cost to Society.” Allergy, 65(6): 776–783.

Heutel, G., N.H. Miller, D. Molitor. 2021. “Adaptation and the Mortality Effects of Temperature across U.S. Climate Regions.” The Review of Economics and Statistics, 103(4): 740–753.

Ito, K., K.R. Weinberger, G.S. Robinson, P.E. Sheffield, R. Lall, R. Mathes, Z. Ross, P.L. Kinney, T.D. Matte. 2015. “The Associations between Daily Spring Pollen Counts, Over-the-counter Allergy Medication Sales, and Asthma Syndrome Emergency Department Visits in New York City, 2002-2012.” Environmental Health, 14: 1–12.

Japan Society of Immunology and Allergology in Otolaryngology. 2021. “The Guidebook for Allergic Rhinitis.” (in Japanese) http://www.jiaio.umin.jp/common/pdf/guide_allergy2021.pdf (accessed March 23, 2022).

Jáuregui, I., J. Mullol, I. Dávila, M. Ferrer, J. Bartra, A. Del Cuvillo, J. Montoro, J. Sastre, A. Valero. 2009. “Allergic Rhinitis and School Performance.” Journal of Investigational Allergology and Clinical Immunology, 19: 32–39.

Jha, A., A. Nauze. 2022. “US Embassy Air-quality Tweets led to Global Health Benefits.” Proceedings of the National Academy of Sciences of the United States of America, 119(44): e2201092119.

Jia, R., H. Ku. 2019. “Is China’s Pollution the Culprit for the Choking of South Korea? Evidence from the Asian Dust.” The Economic Journal, 129(624): 3154–3188.

Kay, G.G. 2000. “The Effects of Antihistamines on Cognition and Performance.” Journal of Allergy and Clinical Immunology, 105(6): S622–S627.

Kondo, K. 2021. “Simulating the Impacts of Interregional Mobility Restriction on the Spatial Spread of COVID-19 in Japan.” Scientific Reports, 11: 18951.

Kuroda, Y. 2022. “The Effect of Pollen Exposure on Consumption Behaviors: Evidence from Home Scanner Data.” Resource and Energy Economics, 67: 101282.

Kuroda, Y., T. Sato, Y. Matsuda. 2022. “How Long do Voluntary Lockdowns Keep People at Home? The Role of Social Capital during the COVID-19 Pandemic.” Data Science and Service Research Discussion Paper No. 125.

Lamb, C.E., P.H. Ratner, C.E. Johnson, A.J. Ambegaonkar, A.V. Joshi, D. Day, N. Sampson, B. Eng. 2006. “Economic Impact of Workplace Productivity Losses due to Allergic Rhinitis Compared with Select Medical Conditions in the United States from an Employer Perspective.” Current Medical Research and Opinion, 22: 1203–1210.

La Nauze, A., E. R. Severnini. 2021 “Air Pollution and Adult Cognition: Evidence from Brain Training.” NBER Working Paper No. 28785.

Lavy, V., G. Rachkovski, O. Yoresh. 2022. “Heads Up: Does Air Pollution Cause Workplace Accidents?” NBER Working Paper No. 30715.

Leard, B., K. Roth. 2019. “Voluntary Exposure Benefits and the Costs of Climate Change.” Journal of the Association of Environmental and Resource Economists, 6(1): 151–185.

Marcotte, D.E. 2015. “Allergy Test: Seasonal Allergens and Performance in School.” Journal of Health Economics, 40: 132–140.

Marcotte, D.E. 2017. “Something in the Air? Air Quality and Children’s Educational Outcomes.” Economics of Education Review, 56: 141–151.

Matsubara, A., M. Sakashita, M. Goto, K. Kawashima, T. Matsuoka, S. Kondo, et al. 2020. “Epidemiological Survey of Allergic Rhinitis in Japan 2019.” (in Japanese) Nippon Jibiinkoka Gakkai Kaiho, 123: 485–490.

McAfoose, J., B.T. Baune. 2009 “Evidence for a Cytokine Model of Cognitive Function.” Neuroscience & Biobehavioral Reviews, 33(3): 355–366.

Meisel, Z.F., J.M. Pines, D. Polsky, J.P. Metlay, M.D. Neuman, C.C. Branas. 2011. “Variations in Ambulance Use in the United States: The Role of Health Insurance.” Academic Emergency Medicine, 18(10): 1036–1044.

MEXT (Ministry of Education, Culture, Sports, Science, and Technology) and JMA (Japan Meteorological Agency). 2020. “Climate Change in Japan 2020.” (in Japanese) https://www.data.jma.go.jp/cpdinfo/ccj/2020/pdf/cc2020_shousai.pdf (accessed March 23, 2022).

MHLW (Ministry of Health, Labour and Welfare). 2009. “Overview of Unexpected Accidental Deaths in 2009.” (in Japanese) https://www.mhlw.go.jp/toukei/saikin/hw/jinkou/tokusyu/furyo10/dl/gaikyo.pdf (accessed March 23, 2022).

MHLW (Ministry of Health, Labour and Welfare). 2020. “Occurrence of Work-related Injuries in 2019.” (in Japanese) https://www.mhlw.go.jp/bunya/roudoukijun/anzeneisei11/rousai-hassei/dl/b19-16.pdf (accessed March 23, 2022).

Moretti, E., M. Neidell. 2011. “Pollution, Health, and Avoidance Behavior: Evidence from the Ports of Los Angeles.” Journal of Human Resources, 46(1): 154–175.

Neidell, M. 2009. “Information, Avoidance Behavior, and Health: The Effect of Ozone on Asthma Hospitalizations.” Journal of Human Resources, 44(2): 450–478.

NPA (National Policy Agency). 2022. “Number of Deaths from Traffic Accidents.” (in Japanese) https://www.npa.go.jp/publications/statistics/koutsuu/toukeihyo.html (accessed March 23, 2022).

Park, J., N. Pankratz, A. Behrer. 2021. “Temperature, Workplace Safety, and Labor Market Inequality.” IZA DP No. 14560.

Sager, L. 2019. “Estimating the Effect of Air Pollution on Road Safety using Atmospheric Temperature Inversions.” Journal of Environmental Economics and Management, 98: 102250.

Santos, C.B., E.L. Pratt, C. Hanks, J. McCann, T.J. Craig. 2006. “Allergic Rhinitis and its Effect on Sleep, Fatigue, and Daytime Somnolence.” Annals of Allergy, Asthma & Immunology, 97(5): 579–587.

Schlenker, W., W.R. Walker. 2016. “Airports, Air Pollution, and Contemporaneous Health.” Review of Economic Studies, 83(2): 768–809.

Schmidt, C.W. 2016. “Pollen Overload: Seasonal Allergies in a Changing Climate.” Environmental Health Perspectives, 24(4): A70–A75.

Seike, T., H. Mimaki, S. Morita. 2015. “Study on the Population Characteristics in a City Center District Utilizing Mobile Spatial Statistics” (in Japanese). Journal of Architecture and Planning, 80(713): 1625–1633.

Skoner, D.P. 2001. “Allergic Rhinitis: Definition, Epidemiology, Pathophysiology, Detection, and Diagnosis.” Journal of Allergy and Clinical Immunology, 108(1): S2–S8.

Smith, A.C. 2016. “Spring Forward at Your Own Risk: Daylight Saving Time and Fatal Vehicle Crashes.” American Economic Journal: Applied Economics, 8(2): 65–91.

Somanathan, E., R. Somanathan, A. Sudarshan, M. Tewari. 2021. “The Impact of Temperature on Productivity and Labor Supply: Evidence from Indian Manufacturing.” Journal of Political Economy, 129(6): 1797–1827.

Terada, M., T. Nagata, M. Kobayashi. 2013. “Mobile Spatial Statistics’ Supporting Development of Society and Industry—Population Estimation Technology for Mobile Spatial Statistics.” NTT DOCOMO Technical Journal, 14: 16–23.

Tokyo Metropolitan Institute of Public Health. 2017. “Hay Fever Patients’ Survey Report.” (in Japanese) http://www.tokyo-eiken.go.jp/files/kjkankyo/kafun/jittai/houkokusho.pdf (accessed March 23, 2022).

Vuurman, E.F., L.L. Vuurman, I. Lutgens, B. Kremer. 2014. “Allergic Rhinitis is a Risk Factor for Traffic Safety.” Allergy, 69(7): 906–912.

Wakamiya, S., S. Matsune, K. Okubo, E. Aramaki. 2019. “Causal Relationships Among Pollen Counts, Tweet Numbers, and Patient Numbers for Seasonal Allergic Rhinitis Surveillance: Retrospective Analysis.” Journal of Medical Internet Research, 21(2): e10450.

Wilken, J.A., R. Berkowitz, R. Kane. 2002. “Decrements in Vigilance and Cognitive Functioning Associated with Ragweed-induced Allergic Rhinitis.” Annals of Allergy, Asthma & Immunology, 89(4): 372–380.

Xing, J., Z. Hu, F. Xia, J. Xu, E. Zou, 2023. “Urban Forests: Environmental Health Values and Risks.” NBER Working Paper No. 31554.

Yamada, T., H. Saito, S. Fujieda. 2014. “Present State of Japanese Cedar Pollinosis: The National Affliction.” Journal of Allergy and Clinical Immunology, 133(3): 632–639.

Zhang, X., X. Chen, X. Zhang. 2018. “The Impact of Exposure to Air Pollution on Cognitive Performance.” Proceedings of the National Academy of Sciences of the United States of America, 115(37): 9193–9197.

Zhang, Y., A.L. Steiner. 2022. “Projected Climate-driven Changes in Pollen Emission Season Length and Magnitude over the Continental United States.” Nature Communications, 13: 1234.

Ziello, C., T.H. Sparks, N. Estrella, J. Belmonte, K.C. Bergmann, E. Bucher, et al. 2012. “Changes to Airborne Pollen Counts across Europe.” PLoS One, 7(4): e34076.

Ziska, L.H., L. Makra, S.K. Harry, N. Bruffaerts, M. Hendrickx, F. Coates, et al. 2019. “Temperature-related Changes in Airborne Allergenic Pollen Abundance and Seasonality across the Northern Hemisphere: A Retrospective Data Analysis.” The Lancet Planetary Health, 3(3): e124–e131.

## References

Kishikawa, R. et al. 2020. “Pollen Calendar of Important Allergenic Airborne Pollen in Japan.” (in Japanese) Japanese Journal of Palynology, 65(2): 55–66.

## References

MIC (Ministry of Internal Affairs and Communications). 2019. “Economic Census.” (in Japanese) https://www.e-stat.go.jp/gis/statmap-search?page=1&type=1&toukeiCode=00200553&toukeiYear=2016&aggregateUnit=H&serveyId=H002005112016&statsId=T000918 (accessed March 23, 2022).

Miyauchi, Y., K. Nakajima, S.J. Redding. 2021. “The Economics of Spatial Mobility: Theory and Evidence Using Smartphone Data.” NBER Working Paper No. 28497.

